# Multi-ancestry proteome-phenome-wide Mendelian randomization offers a comprehensive protein-disease atlas and potential therapeutic targets

**DOI:** 10.1101/2024.10.17.24315553

**Authors:** Chen-Yang Su, Adriaan van der Graaf, Wenmin Zhang, Dong-Keun Jang, Susannah Selber-Hnatiw, Ta-Yu Yang, Guillaume Butler-Laporte, Kevin Y. H. Liang, Fumihiko Matsuda, Maria C. Costanzo, Noel P. Burtt, Jason Flannick, Sirui Zhou, Vincent Mooser, Tianyuan Lu, Satoshi Yoshiji

**Affiliations:** McGill Genome Centre, McGill University, Montréal, Québec, Canada; Quantitative Life Sciences, McGill University, Montreal, Québec, Canada; Department of Computational Biology, University of Lausanne, Lausanne, Switzerland; Montreal Heart Institute, Université de Montréal, Montreal, Québec, Canada; Programs in Metabolism and Medical & Population Genetics, The Broad Institute of MIT and Harvard, Cambridge, MA, USA; Department of Human Genetics, McGill University, Montreal, Québec, Canada; Center for Genomic Medicine, Graduate School of Medicine, Kyoto University, Kyoto, Japan; Lady Davis Institute, Jewish General Hospital, McGill University, Montréal, Québec, Canada; Harvard Medical School, Boston, MA, USA; Department of Population Health Sciences, University of Wisconsin-Madison, Madison, WI, USA; Department of Biostatistics and Medical Informatics, University of Wisconsin-Madison, Madison, WI, USA; Center for Genomic Science Innovation, University of Wisconsin-Madison, Madison, WI, USA; Center for Demography of Health and Aging, University of Wisconsin-Madison, Madison, WI, USA

## Abstract

Circulating proteins influence disease risk and are valuable drug targets. To enhance the discovery of protein-phenotype associations and identify potential therapeutic targets across diverse populations, we conducted proteome-phenome-wide Mendelian randomization in three ancestries, followed by comprehensive sensitivity analyses. We tested the potential causal effects of up to 2,265 unique proteins on a curated list of 355 distinct phenotypes, assessing 726,035 protein-phenotype pairs in European, 33,078 in African, and 115,352 in East Asian ancestries. Notably, 119 proteins were instrumentable only in African ancestry and 17 proteins only in East Asian ancestry due to allele frequency differences that are common in these ancestries but rare in European ancestry. We identified 3,949, 56, and 325 unique protein-phenotype pairs in European, African, and East Asian ancestries, respectively, and assessed their druggability using multiple databases. We highlighted the causal role of IL1RL1 in inflammatory bowel diseases, supported by multiple orthogonal lines of evidence. Taken together, this study underscores the importance of multi-ancestry inclusion and offers a comprehensive atlas of protein-phenotype associations across three ancestries, enhancing our understanding of proteins involved in disease etiology and potential therapeutic targets. Results are available at the Common Metabolic Diseases Knowledge Portal (https://broad.io/protein_mr_atlas).

## Introduction

Circulating proteins play a major role in a multitude of biological pathways^1–3^, are important biomarkers for disease diagnosis, prognosis, and prevention^4–6^, and serve as valuable drug targets^7–10^. Current high-throughput proteomics platforms measure thousands of circulating plasma proteins. With measurements in large cohorts, recent studies have conducted genome-wide association studies (GWAS) to evaluate genetic variants associated with the abundances of thousands of proteins^11–14^. We can leverage these proteomic GWAS to find causal proteins for diseases and prioritize drug targets through Mendelian randomization (MR)^15,16^. In this case, MR uses genetic variants associated with variation in plasma protein levels (known as protein quantitative trait loci, pQTLs) as instruments to measure the causal effect of an exposure (protein abundance) on an outcome (a complex trait). This is shown to reduce confounding and reverse causation biases affecting many epidemiological studies, provided that three key assumptions are met: (1) the genetic variant is associated with the exposure, (2) there is no confounding of the instrument-outcome association, and (3) the genetic variant influences the outcome solely through the exposure.

Despite applications of proteogenomics in elucidating disease mechanisms and identifying potential therapeutic targets^17–23^, previous pQTL studies^18,19^ have been based on smaller sample sizes and measured fewer proteins, assessed limited outcomes, and most importantly, have predominantly focused on individuals of European ancestry. African ancestry proteomics cohorts have emerged in recent years^13,14,24^, yet existing studies have assessed a limited number of outcomes and comparisons with other ancestries have also been limited^19^. Similarly, in East Asian ancestries, few large-scale proteome-phenome wide MR studies exist^25,26^, largely due to the lack of publicly available pQTLs, despite the presence of existing East Asian ancestry biobanks providing hundreds of publicly available GWAS outcomes^27,28^.

The inclusion of multiple ancestries in a proteome-phenome wide atlas of associations can potentially offer significant benefits. Diverse ancestry inclusion in MR leverages the natural variations in genetic architecture present across populations, which allows analyses on otherwise unseen genetic variation, increases in statistical power due to allele frequency increases, and differentiation of causal effect magnitude across populations^29,30^. Combined, this approach may be able to identify a greater number of instrumentable proteins (i.e., proteins that can be tested in MR), which may lead to an increase in discoveries. By leveraging insights obtained through instrumentable proteins in one ancestry, the identified protein-phenotype associations could contribute to our understanding of disease mechanisms, which may be generalizable across different ancestries and potentially benefit all populations. For instance, PCSK9 loss-of-function variants Y142X and C679X predispose individuals to naturally lower LDL cholesterol levels. These mutations were found to be common in African Americans but rare in European Americans^31^ and inspired the development of PCSK9 inhibitors mimicking these variants to effectively reduce LDL cholesterol levels and risk of cardiovascular events^32,33^. In addition to providing insights into novel therapeutic targets, enhancing diversity in genomic studies is crucial to ensure equitable health outcomes and address imbalances in health disparities across populations^34^.

Here, we combined four of the largest European ancestry proteomics cohorts (*n* = up to 35,559), two of the largest African proteomics cohorts (*n* = up to 1,871), and a new East Asian proteomics cohort (*n* = 1,823). We then performed MR and colocalization analyses on a curated list of the most recent and largest ancestry-specific outcome GWAS to date for 179 European and 26 African ancestry outcomes, as well as 206 East Asian ancestry outcomes from Biobank Japan to construct an atlas of protein-phenotype associations. We integrated our findings with multiple drug databases to assess the druggability of the associations and highlighted novel targets. Overall, our study supports the prioritization of thousands of protein-phenotype associations and provides a comprehensive, updated resource for the community, significantly expanding our understanding of these associations. Results are publicly available at the Common Metabolic Diseases Knowledge Portal (https://broad.io/protein_mr_atlas).

## Results

The overall study design is shown in **Fig. 1**.

**Figure 1.**
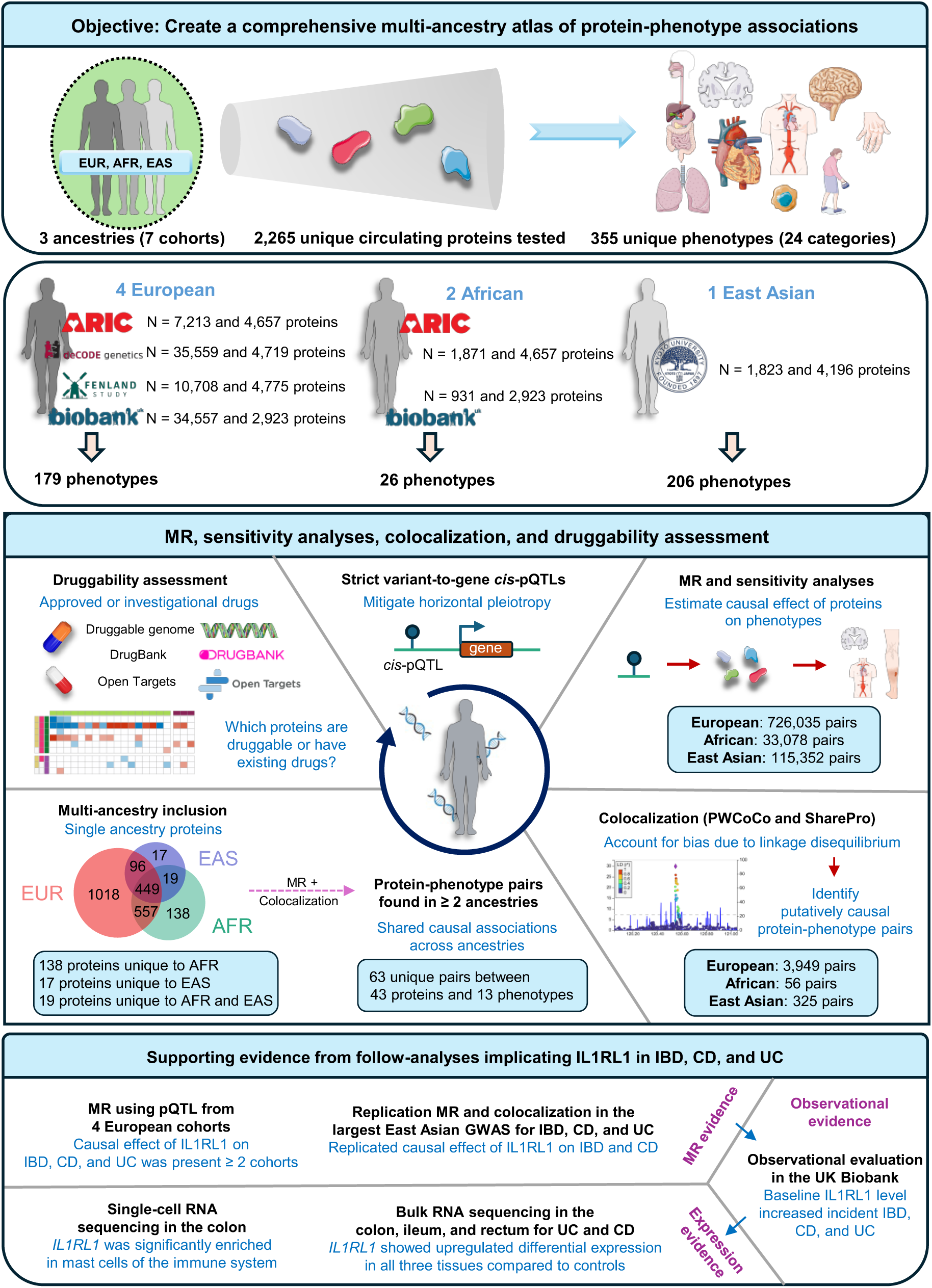
Study design. We assessed the causal role of 2,265 circulating plasma proteins across three ancestries (four European, two African, and a new East Asian ancestry cohort) on up to 355 phenotypes/outcomes with extensively curated GWAS in each ancestry. For European and African ancestry outcomes, we collected the largest and most recent GWAS available as of February 2024 for 179 and 26 outcomes, respectively, while 206 GWAS from BioBank Japan were used for East Asian ancestry outcomes. We implemented a unique approach for defining *cis*-pQTLs using a multi-step process combining a strict *cis*-pQTL definition with Open Targets Genetics variant-to-gene score^35^ filtering to minimize risk of horizontal pleiotropy, which we term “strict variant-to-gene (V2G) *cis*-pQTLs”. Then, we performed multi-ancestry proteome-wide MR and colocalization analyses using PWCoCo^18,36^ and SharePro^37^, a new colocalization method we developed, on the curated phenotypes to identify causal protein-phenotype associations. Next, we overlapped prioritized protein-phenotype pairs with drug databases such as the druggable genome^38^, DrugBank^39^, and the Open Targets platform^40^. Finally, as an illustrative example of the value of our atlas, we pinpoint IL1RL1 and explore its role in inflammatory bowel disease using multiple lines of evidence. EUR: European, AFR: African, EAS: East Asian; MR: Mendelian randomization; IBD: Inflammatory bowel disease; CD: Crohn’s disease; UC: Ulcerative colitis.

### 1. Genetic instrument selection for proteins

We summarize the selected genetic instruments in **Supplementary Table 1**. Briefly, we used proteomic GWAS from four European ancestry cohorts: ARIC (4,657 proteins measured in up to 7,213 individuals)^13^, deCODE (4,719 proteins measured in up to 35,559 individuals)^12^, Fenland (4,775 proteins measured in up to 10,708 individuals)^11^, and UKB-PPP (2,923 proteins measured in up to 34,557 individuals)^14^. For African ancestry, we analyzed proteomic GWAS from two cohorts: ARIC (4657 proteins measured in up to 1,871 individuals)^13^ and UKB-PPP (2923 proteins measured in up to 931 individuals)^14^. Additionally, we included a new East Asian ancestry cohort, the Kyoto University Nagahama cohort which measured 4,196 proteins in up to 1,823 individuals.

#### 1.1. Unifying cis-pQTL across cohorts

We re-defined *cis*-pQTLs across proteomics cohorts as pQTLs within 500 kb of the transcription start site (TSS) of the protein-coding gene, with independence and significance defined with linkage disequilibrium (LD) *r*^2^ < 0.001 and *P* < 5 × 10^-8^, respectively. Independent pQTLs outside the *cis*-region were labeled as *trans*-pQTLs. We analyzed *cis*-pQTLs because they are more likely to have direct biological effects on the proteins of interest^41,42^. We verified that newly defined *cis*-pQTLs had high concordance within ancestries (**Supplementary Note 1A**).

#### 1.2. Identifying strict V2G cis-pQTLs

We mitigated the risk of horizontal pleiotropy by using a unique approach to select genetic instruments which we term strict V2G *cis*-pQTLs (**Supplementary Note 1B**). Strict V2G *cis-* pQTLs are associated with a single protein-coding gene (“strict”) and have the strongest link to the corresponding protein-coding gene based on the Open Targets Genetics V2G score, which uses multiple sources of evidence to map variants to genes (“V2G”)^35^ (**Extended Data Fig. 1** and **Supplementary Tables 2–8**). We also assessed if these strict V2G *cis*-pQTLs were protein altering variants (PAVs), as PAVs may affect protein structure and lead to bias in effect size estimation^1^. We found that only 70 of 7,399 (0.95%), 30 of 1,684 (1.8%), and 9 of 663 (1.4%) strict V2G *cis*-pQTLs in European, African, and East Asian ancestries, respectively, were PAVs or in high LD with PAVs of high impact. Notably, strict V2G *cis*-pQTLs were more enriched for *cis*-eQTLs, indicating their role in local gene regulation impacting protein levels (**Extended Data Fig. 2a**). They had significantly larger effects on protein levels (**Extended Data Fig. 2b**) and were significantly closer to the TSS of the corresponding protein-coding gene (**Extended Data Fig. 3**). Hence, strict V2G selection of pQTLs limits the risks of violating MR assumptions (specifically no horizontal pleiotropy) while still retaining adequate statistical power. Specifically, all proteins in each cohort had F-statistics above 10, suggesting that the risk of weak instrument bias is limited^43^ (**Supplementary Table 9**).

### 2. Multi-ancestry inclusion is important in instrumenting proteins and reveals population-specific variants

Across the three ancestries, we were able to instrument 2,265 unique proteins with 449 of these being shared across all three ancestries (**Fig. 2a**). Specifically, we instrumented 2,110 proteins in European, 1,144 in African, and 581 in East Asian ancestries (see **Supplementary Note 1C** for cohort level description and **Extended Data Fig. 4**). We identified 1,018 proteins that were unique to individuals of European ancestry. Including African ancestries allowed for an additional 138 proteins, with 119 unique to African ancestry and 19 shared with East Asian ancestries. Including an East Asian ancestry cohort allowed an additional 17 unique proteins (**Fig. 2a**). Altogether, these findings underscore the value of including multiple ancestries in proteomic analyses.

**Figure 2.**
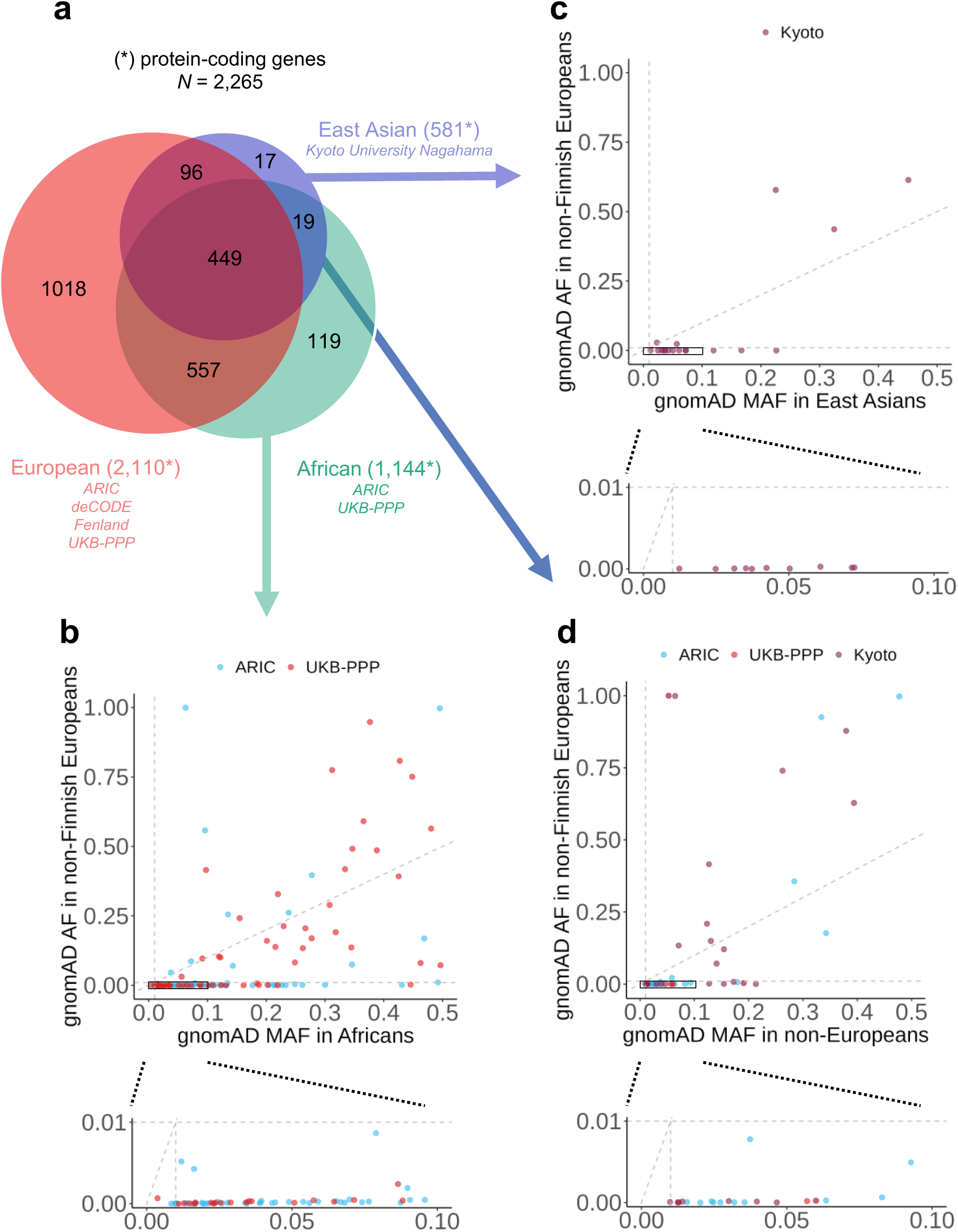
Cross ancestry comparison of overlapping instrumentable protein-coding genes. (a) Overlapping instrumentable protein-coding genes across three ancestries. Protein-coding genes were quantified based on Ensembl gene IDs. Red: Unique protein-coding genes across four European ancestry proteomics cohorts; green: unique protein-coding genes across two African ancestry proteomics cohorts; blue: East Asian ancestry proteomics cohort protein-coding genes. (b) gnomAD minor allele frequencies (MAF) of *cis*-pQTLs instrumenting the 119 proteins uniquely instrumentable in African ancestry when plotted against corresponding gnomAD non-Finnish European allele frequency (AF). The black box in the top plot is zoomed in and shown in the bottom plot. Blue: ARIC African *cis*-pQTLs. Red: UKB-PPP African ancestry cohort *cis*-pQTLs. (c) gnomAD minor allele frequencies (MAF) of *cis*-pQTLs instrumenting the 17 proteins uniquely instrumentable in East Asian ancestry when plotted against corresponding gnomAD non-Finnish European AF. The black box in the top plot is zoomed in and shown in the bottom plot. Purple: Kyoto University Nagahama East Asian ancestry cohort *cis*-pQTLs. (d) gnomAD MAF minor allele frequencies (MAF) of *cis*-pQTLs instrumenting the 19 proteins common to African and East Asian but not European ancestries when plotted against corresponding gnomAD non-Finnish AF. The black box in the top plot is zoomed in and shown in the bottom plot. Blue: ARIC African *cis*-pQTLs. Red: UKB-PPP African *cis*-pQTLs. Purple: Kyoto University Nagahama East Asian ancestry cohort *cis*-pQTLs.

Next, to better understand the unique proteins in non-European ancestries, we compared the allele frequencies of their genetic instruments using gnomAD^44^. Of the 130 genetic instruments for the 119 proteins unique to African ancestries, 89 (68.5%) had a minor allele frequency (MAF) below 0.01 in European ancestries (**Fig. 2b**). Similarly, for the 18 genetic instruments for the 17 proteins unique to East Asian ancestry, 13 (72.2%) had a MAF below 0.01 in European ancestry (**Fig. 2c**). The majority (29, 63.0%) of the 46 unique genetic instruments for the 19 proteins instrumentable by both African and East Asian but not by European ancestries, were rare (MAF < 0.01) in European ancestry (**Fig. 2d**). These results suggest that populational allele frequency differences allow for more proteins to be included in MR analyses. We refer to them as uniquely instrumentable proteins.

### 3. Two-sample Mendelian randomization

We performed two-sample MR using strict V2G *cis*-pQTLs as instrumental variables to determine causal proteins implicated in human complex traits and diseases. Across three ancestries, we considered 355 unique outcomes (**Fig. 3a**) pertaining to 24 phenotype categories (**Fig. 3b**). These phenotypes/outcomes (as of February 2024) included 179 of the most up to date and largest phenotypic GWAS available for European ancestries (**Supplementary Table 10**), 26 of the largest available GWAS for African ancestries (**Supplementary Table 11**), and 206 from BioBank Japan for East Asian ancestries (**Supplementary Table 12**).

**Figure 3.**
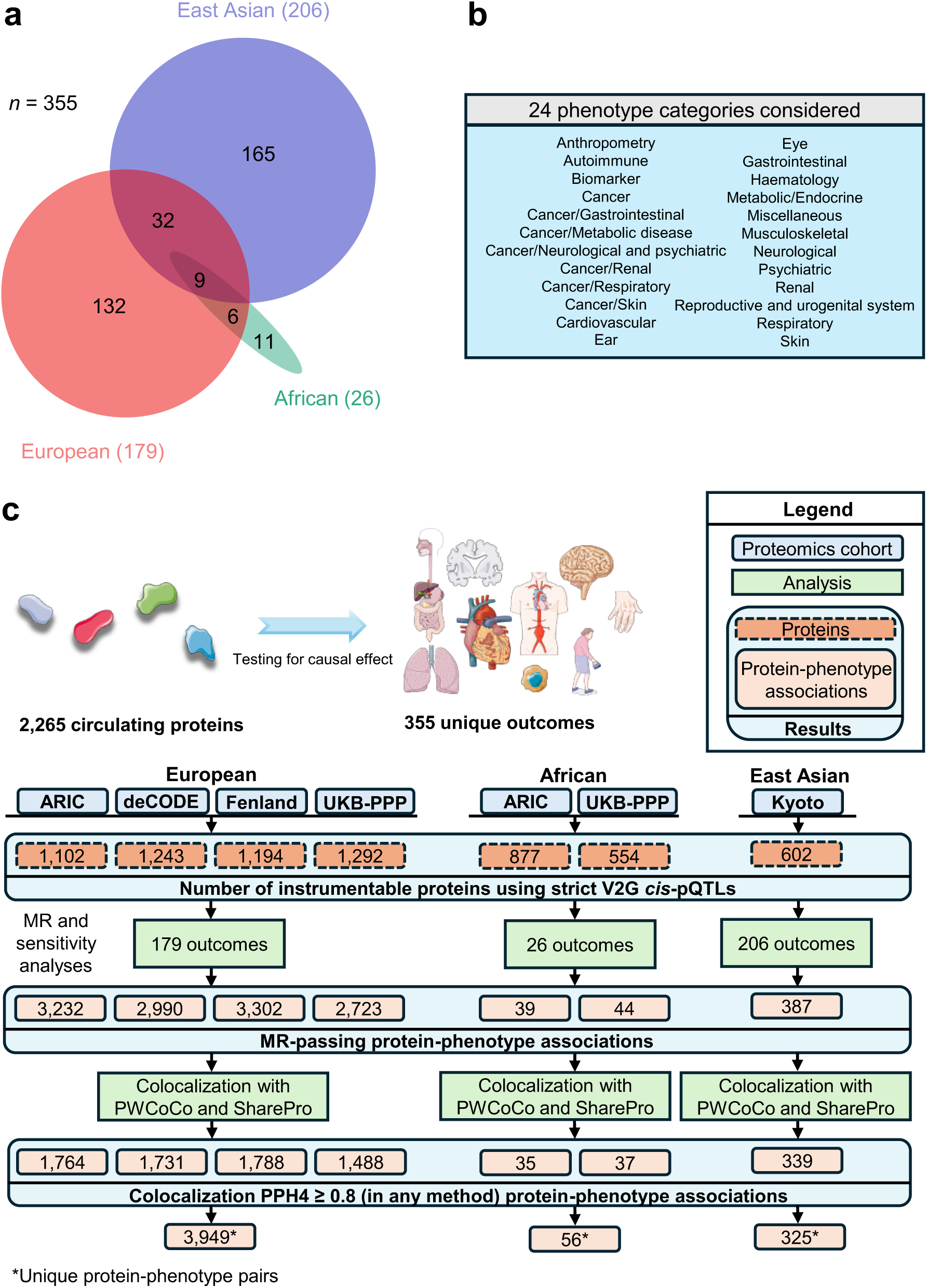
MR analyses to determine the effects of proteins on phenotypes. (a) Curated GWAS phenotypes common between different ancestries. European (red), African (green), and East Asian (blue) ancestries. (b) Phenotype categories for outcomes used in all three ancestries. (c) Flowchart summary of MR and colocalization analyses to identify protein-phenotype associations for each cohort.

We tested a total of 726,035 protein-phenotype pairs in European ancestries (173,748 for ARIC, 182,433 for deCODE, 180,018 for Fenland, and 189,836 for UKB-PPP), 33,078 in African ancestries (20,442 for ARIC and 12,636 for UKB-PPP), and 115,352 in East Asian ancestries (Kyoto University Nagahama cohort) (see **Data availability**). To control for false positives, we applied a Benjamini-Hochberg-corrected *P* value (false discovery rate, FDR)^45^ threshold of 0.05 (5%) per cohort in each ancestry^19^. We note that Bonferroni correction is overly stringent given that (i) proteins are correlated with one another, and (ii) we tested the same protein-phenotype associations across cohorts, making these tests not independent. Nevertheless, we also provide the most stringent Bonferroni corrected associations in section 5. Results were further filtered based on multiple sensitivity analyses robust to MR assumption violations and retained associations are now termed “MR-passing”. Assessment of sample overlap between European ancestry GWAS outcomes that were based on the UK Biobank and proteomics from the European ancestry UKB-PPP cohort was performed (**Supplementary Note 2**). Of the tested associations, a total of 12,247 associations were considered MR-passing in European, 83 in African, and 387 in East Asian ancestries (**Supplementary Table 13**).

### 4. Colocalization analyses

MR results may be confounded by independent causal variants in LD^46,47^. To guard against such bias, for each MR-passing protein-phenotype pair, we performed colocalization analysis using two methods, PWCoCo^18,36^ and SharePro^37^, to verify that the protein abundance and the tested outcome share the same genetic signals (**Fig. 3c**). An MR-passing protein-phenotype pair was considered putatively causal if it was supported by at least one method with a posterior colocalization probability (PP_max_) ≥ 0.8. Across all cohorts and all three ancestries, 56.5% (7,182 out of 12,717) of MR-passing associations were supported by colocalization evidence (**Supplementary Table 13**).

### 5. Putatively causal protein-phenotype associations

Upon MR, sensitivity analyses, and colocalization, we identified 3,949 unique putatively causal protein-phenotype pairs in European (**Extended Data Fig. 5a** and **Supplementary Table 14**), 56 in African (**Extended Data Fig. 5b** and **Supplementary Table 15**), and 325 in East Asian ancestries (**Extended Data Fig. 5c** and **Supplementary Table 16**). Here, we use protein to refer to protein-coding genes to harmonize across SomaScan and Olink platforms. Results are also hosted at https://broad.io/protein_mr_atlas. We described cohort level associations and proteins implicated in a multitude of phenotypes in **Supplementary Notes 3 and 4**. Particularly, 1,617, 30, and 135 unique associations in European, African, and East Asian ancestries further withstood the Bonferroni correction accounting for the total number of tests across all cohorts and all ancestries (*P* < 0.05 / 874,465 = 5.7 × 10^-8^), however, this threshold is likely overly conservative due to many proteins being correlated with one another as well as non-independent tests from the same protein-phenotypes associations being tested across cohorts (**Supplementary Tables 14 – 16**).

In European ancestries, the 3,949 significant unique protein-phenotype pairs identified in European ancestries involved putatively causal effects between 995 proteins and 146 phenotypes, of which only 56 (1.4%) showed discordant MR estimates in one of the tested cohorts (**Supplementary Table 14**). These discrepancies could be attributed to population differences or variations in proteomic assays, such as differential effects from SomaScan aptamers targeting different domains compared to Olink assays. Of the 3,893 remaining putatively causal European ancestry pairs between 991 proteins and 146 outcomes, 1,692 (43.5%) of the identified putatively causal associations were from 452 proteins uniquely instrumentable by European ancestries. Further, 3,853 (99.0%) associations have not been previously reported by earlier proteome-phenome wide MR studies from Zheng et al.^18^ and Zhao et al.^19^.

#### 5.1. Cardiovascular and autoimmune diseases

We demonstrate the highly interconnected nature between proteins and outcomes by highlighting cardiovascular (**Fig. 4a** and **Supplementary Note 5A**) and autoimmune phenotypes (**Fig. 4b**) in European ancestries given their significant impact on health. Cardiovascular outcomes were influenced by up to 103 proteins (median: 7) while some proteins influenced up to 8 cardiovascular outcomes (median: 1) (**Fig. 4a**). For instance, we found that an s.d. increase in genetically predicted ULK3 levels increases systolic and diastolic blood pressure, pulse pressure, and hypertension. ULK3 is a nuclear kinase which may contribute to vascular disease by mediating autophagy dysregulation^48^. In concordance, a recent study showed that functional splicing effects of ULK3 can contribute to coronary artery disease (CAD)^49^ suggesting effective modulation of ULK3 may be beneficial for reducing cardiovascular risk.

**Figure 4.**
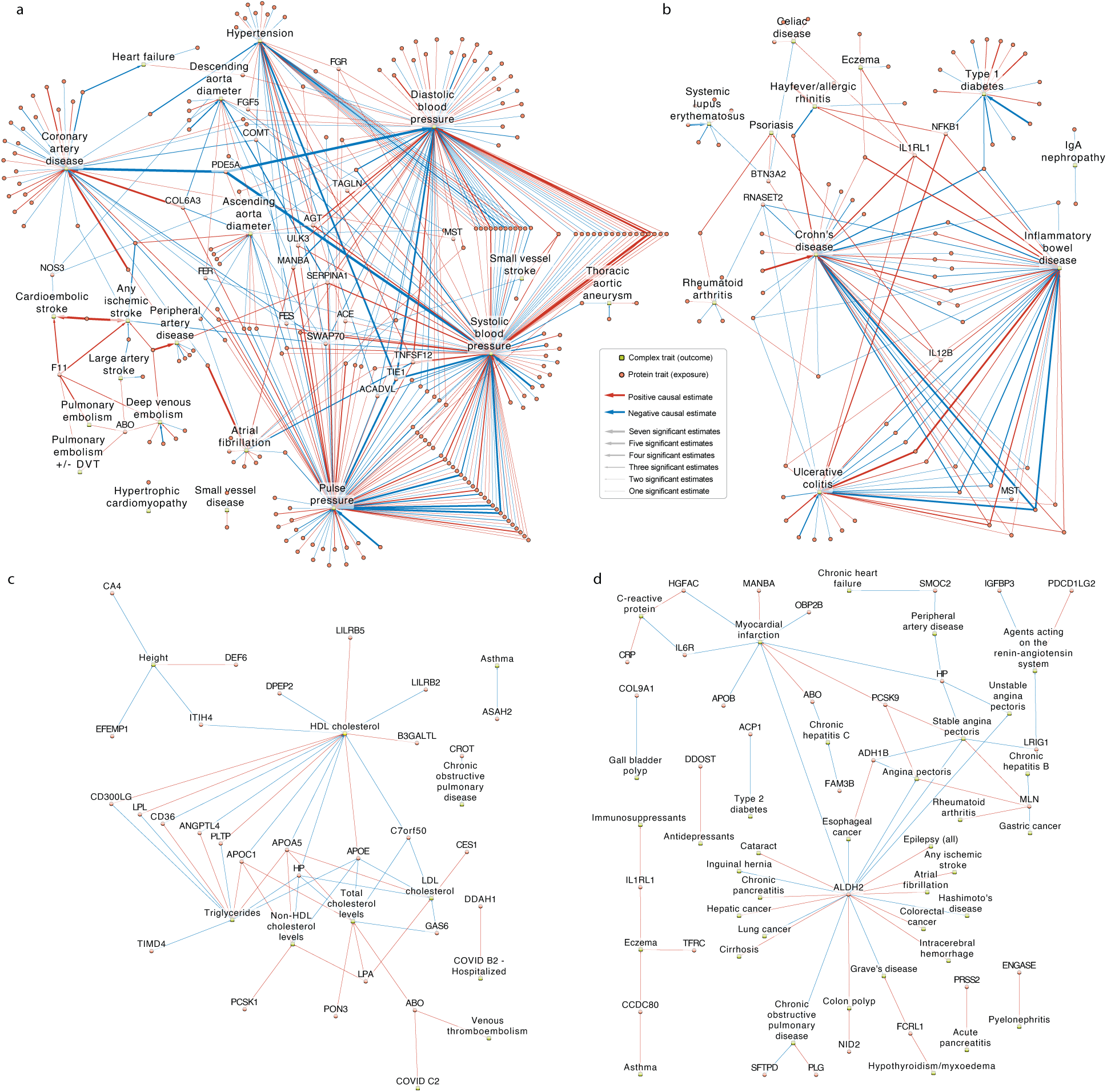
Protein-phenotype network plots. Red arrows indicate a positive causal estimate of the protein on the phenotype while blue arrows indicate a negative causal estimate of the protein on the phenotype. (a) Significant estimates between proteins (orange circles) and cardiovascular traits (green rectangles) both derived from European ancestry individuals. All cardiovascular traits are named, as well as the protein traits that have a significant effect on 4 or more cardiovascular traits. Arrow thickness representing number of significant estimates indicates how often a protein measurement has a causal effect on the outcome trait. The maximum number of significant estimates is 7 which is larger than the total number of European ancestry proteomics studies, 4, due to the presence of more than one SomaScan aptamer for that protein. For simplicity, we only depict protein-phenotype pairs in which all European ancestry cohorts showed concordant direction of effect estimates. (b) Significant estimates between proteins (orange circles) and autoimmune traits (green rectangles) both derived from European ancestry individuals. All autoimmune traits are named, as well as the protein traits that have a significant effect on 4 or more autoimmune traits. Arrow thickness representing number of significant estimates indicates how often a protein measurement has a causal effect on the outcome trait. For simplicity, we only depict protein-phenotype pairs in which all European ancestry cohorts showed concordant direction of effect estimates. (c) Significant estimates between proteins (orange circles) and all traits (green rectangles) both derived from African ancestry individuals. All arrows are the same thickness to indicate support of the association in a single study. (d) Significant estimates between proteins (orange circles) and binary traits (green rectangles) both derived from East Asian ancestry individuals. All arrows are the same thickness and indicate support of the association in a single study.

Similarly, we found high interconnectedness between autoimmune phenotypes and proteins (**Fig. 4b**). Autoimmune phenotypes were influenced by a median of 7 proteins; the highest number of associations were for Crohn’s disease (CD) (39 proteins), inflammatory bowel disease (IBD) (35 proteins), and ulcerative colitis (UC) (28 proteins). Proteins, on the other hand, influenced a median of 2 autoimmune phenotypes. IL1RL1 and IL12B influenced the largest number of outcomes (6 and 4 outcomes, respectively) (**Fig. 4b**) with both increasing risk of IBD, CD, and UC. IL12B is targeted by a commercially available drug ustekinumab for CD^50^; however, there are currently no approved drugs targeting IL1RL1, although there has been increasing interest in modulating IL1RL1 for treating various conditions^51,52^. For example, tozorakimab targets IL-33, the interleukin that binds IL1RL1 (ST2)^53^ and anakinra targets IL-1^54^, a closely related interleukin.

Other potentially novel findings involving other disease categories in European ancestries are highlighted in **Supplementary Note 5B**.

#### 5.2. Protein-phenotype associations in African and East Asian ancestries

In African ancestries, we identified 56 unique protein-phenotype associations involving 28 proteins and 11 phenotypes (**Fig. 4c** and **Supplementary Table 15**). Of these 56 pairs, 55 (98.2%) have not been previously reported by earlier proteome-phenome wide MR studies in African ancestries^19^. Notably, 11 (19.6%) protein-phenotype pairs involving four proteins, APOE, C7orf50, CD300LG, and PON3, were uniquely instrumentable in African ancestries (**Extended Data Fig. 6a**). For instance, increased PON3 levels was associated with increased total cholesterol (β_UKB-PPP_ = 0.03, 95% CI = 0.02–0.05, *P =* 1.1 × 10^-5^, PP_max_ = 0.95). *PON3* is highly expressed in the liver and previously implicated in cholesterol metabolism^55,56^ and atherosclerosis progression. This is notable as PON3 was not instrumentable in European ancestries, thus, African ancestries are uniquely suitable to identify biologically plausible protein-phenotype associations.

Furthermore, the Million Veteran Program^57^ recently released summary statistics for additional traits in up to 635,969 individuals. Using this resource, we tested the causal effect of the 119 proteins uniquely instrumentable in African ancestries on cardiovascular and autoimmune-related binary outcomes. Using the African ARIC proteomics cohort, we identified 7 associations with cardiovascular outcomes and no associations with autoimmune-related outcomes (**Supplementary Note 5C**). Notably, increased PCYOX1 levels were associated with a reduced risk of coronary atherosclerosis, atrial fibrillation, and flutter, indicating its protective effect. PCYOX1 plays a role in oxidative stress and lipid metabolism and has been implicated in atherosclerosis in rodent studies^58^, supporting our findings. This suggests that as more outcomes with larger sample size, particularly binary disease outcomes, become available in African ancestries, we can better leverage uniquely instrumentable proteins to uncover additional protein-disease associations.

In East Asian ancestries, 339 putatively causal associations were identified with 325 unique protein-phenotype pairs involving 110 proteins and 86 phenotypes (**Fig. 4d** and **Supplementary Table 16**). Among them, increased SMOC2 level was associated with decreased risk of peripheral artery disease (PAD) (OR = 0.82, 95% CI = 0.74–0.90, *P =* 5.5 × 10^-5^, PP_max_ = 0.97). *SMOC2* is highly expressed in arteries and plays roles in endothelial cell proliferation, angiogenesis, and matrix assembly^59^. Notably, in European ancestries, increased SMOC2 was associated with decreased pulse pressure, implicating favorable cardiovascular effects across ancestries.

We also found 8 proteins which were only instrumentable in East Asian ancestry due to lack of genome-wide significant *cis*-pQTLs in other ancestries or stringent strict V2G instrument selection (**Supplementary Note 5D**). These 8 proteins were ALDH2, ANXA7, APOA1, DDOST, GSS, PLA2G7, PRSS2, and UGT1A1 which together accounted for 67 (20.6%) protein-phenotype associations (**Extended Data Fig. 6b** and **Supplementary Note 5D**). For instance, increased PRSS2 levels in East Asian ancestries was associated with an OR of 2.05 for acute pancreatitis (95% CI: 1.42–2.98, *P =* 1.43 × 10^-4^, PP_max_ = 0.95) which has been previously validated in European only proteome-wide analyses^60^ suggesting concordant effects across ancestries.

### 6. Concordant effects across ancestries

We evaluated the direction of effect for protein-phenotype associations that passed MR and colocalization analyses across ancestries since concordance across ancestries may strengthen the evidence of broader applicability of therapeutic targets. We found 186 total protein-phenotype/outcome pairs with 63 unique pairs between 43 proteins and 13 outcomes (**Supplementary Table 17** and **Fig. 5**). Among these, 51 pairs (81.0%) had concordant effects across ancestries, while 12 pairs (19.0%) had inconsistent effects (**Supplementary** Figure 3 and **Supplementary Note 6**). Notable concordant associations included PCSK9 and LDL cholesterol in European and East Asian ancestries, and haptoglobin (HP), which binds free hemoglobin to prevent oxidative damage, with LDL cholesterol and total cholesterol across all three ancestries. Angiopoietin-related protein 4 (ANGPTL4) was negatively associated with HDL cholesterol and positively with triglycerides in European and African ancestries, while lipoprotein-lipase (LPL) showed opposite associations in these ancestries. ANGPTL4 has been shown to act as a local inhibitor of LPL^61^ which serves as the rate-limiting enzyme in the degradation of triglycerides^62^, concordant with our findings. While no approved drugs exist for ANGPTL4, inhibition of a closely related protein ANGPTL3 through an RNA interference therapy zodasiran is currently undergoing clinical trials for cholesterol lowering^63^. Thus, many of these biological effects were validated in our multi-ancestry analyses. We note that while concordant effects across ancestries provide strong evidence of support, discordant effects in MR do not automatically indicate biologically discordant effects across ancestries, which could be due to epitope-binding effects or other technical variations^64^.

**Figure 5.**
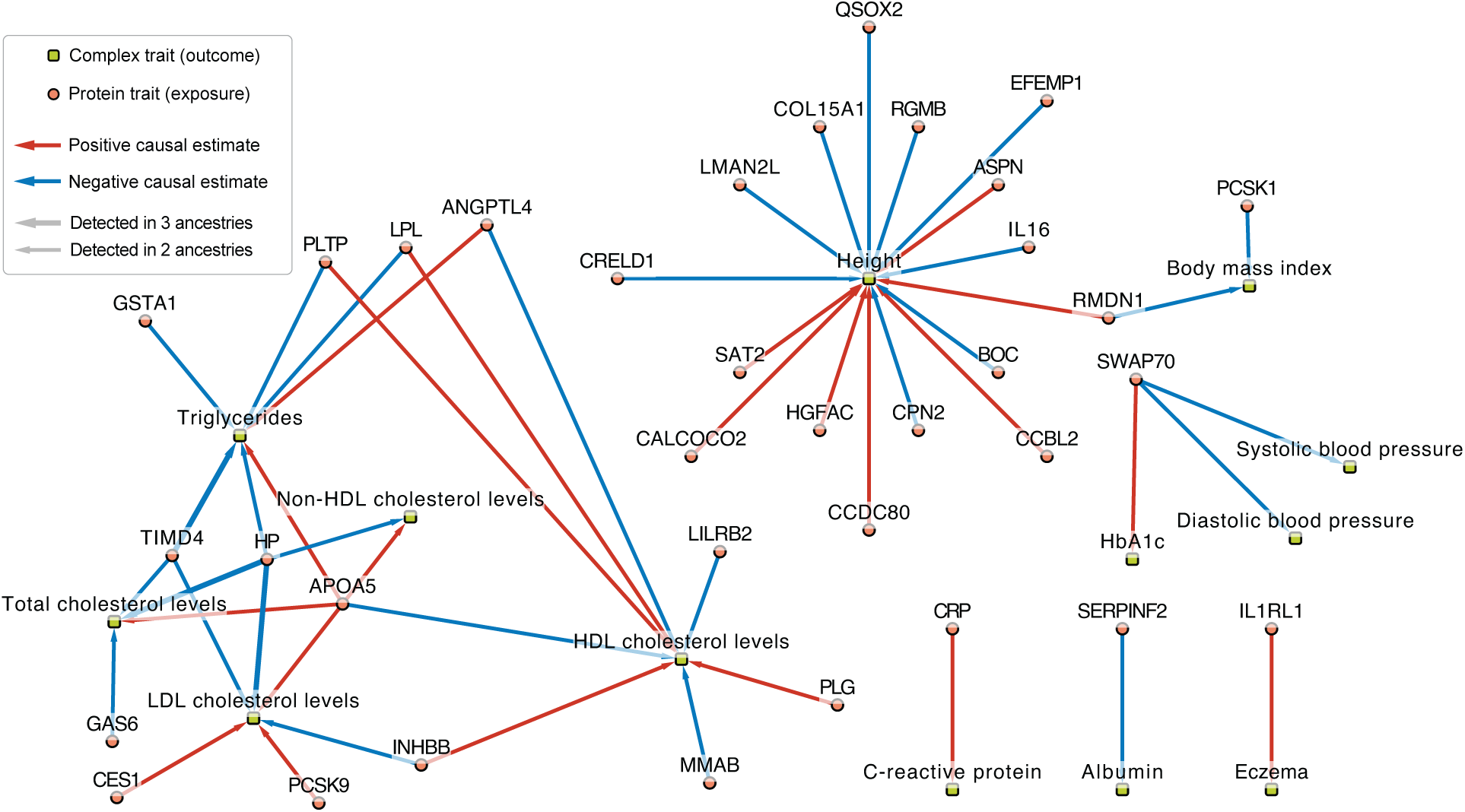
Multi-ancestry network plot for protein-phenotype pairs present in two or more ancestries. All estimates shown had evidence in European ancestry and at least one other ancestry (African or East Asian ancestry). Significant estimates between proteins (orange circles) and traits (green rectangles). Arrow thickness indicates the number of ancestries in which a protein measurement has a causal effect on the phenotype. For simplicity, we only depict estimates when cohorts within the same ancestry and across different ancestries showed concordant direction of effect estimates. Red arrows indicate a positive causal estimate of the protein on the phenotype while blue arrows indicate a negative causal estimate of the protein on the phenotype.

### 7. Druggability

#### 7.1. Druggability of the instrumentable protein-coding genes

Across all ancestries, between 60% to 67% of instrumentable protein-coding genes overlapped with the druggable genome^38^, which classifies genes into Tier 1, 2, or 3 according to druggability, and between 20.5% and 23.6% overlapped with Tiers 1 and 2 (**Supplementary Table 18** and **Supplementary Note 7A**). Druggability tiers for instrumentable proteins are in **Supplementary Tables 19–25**.

Next, we incorporated the druggable genome^38^, DrugBank^39^, and Open Targets Platform^40^ to determine which instrumentable proteins overlapped at least one database. Proteins overlapping with DrugBank^39^ had approved or investigational drugs available while those overlapping Open Targets have information on their clinical development phase and status of protein-drug-disease combinations. Cross-ancestry comparison stratified by proteomics platform for instrumentable proteins overlapping at least one database shows that in SomaScan v4, African ancestry adds 68 additional targets beyond European ancestry cohorts (**Extended Data Fig. 7a**), while East Asian ancestry contributes 34 more targets (**Extended Data Fig. 7b**). In Olink Explore 3072, including African ancestry presents 62 additional targets (**Extended Data Fig. 7c**). These findings suggest that data from African and East Asian ancestries could enhance drug development by offering more potential therapeutic targets.

#### 7.2. Druggability of protein-phenotype pairs by integrating the druggable genome, DrugBank, and Open Targets Platform

Across three ancestries, among the 1,037 number of protein-coding genes that have at least one protein-phenotype association, 669 (64.5%) were present in at least one database. Specifically, 579 (55.8%) protein-coding genes overlapped with the druggable genome^38^, 350 (33.8%) had approved or investigational drugs in DrugBank^39^, and 191 (18.4%) overlapped with the Open Targets Platform^40^ (**Supplementary Table 26**). Notably 32 (3.1%) were unique to non-Europeans. This highlights that multi-ancestry inclusion can expand the list of actionable druggable associations. Overlap with the druggable genome and DrugBank stratified by ancestry is presented in **Supplementary Note 7B**.

We found that higher levels of MANBA, a Tier 2 target, increased risk of atrial fibrillation in European ancestry (**Fig. 6a**) and myocardial infarction in East Asian ancestry (**Fig. 6b**). Currently no drugs exist for MANBA but its role in lysosomal metabolism suggests that modulating its activity could have therapeutic potential. Further, increased ANGPTL4, a Tier 3 target, leads to increased triglycerides and decreased HDL cholesterol levels in African ancestries (**Fig. 6c**) and increased CAD risk in European ancestries (**Supplementary Note 7C**) supporting its potential as a therapeutic target. Notably, we found that increased STAT3, a Tier 1 target, increased risk of IBD and its subtypes, CD, and UC (**Fig. 6d**). Danvatirsen, a STAT-3 mRNA 3’UTR antisense inhibitor has been undergoing phase 1 and 2 clinical trials for multiple cancer types and may be potentially repurposed for IBD. Currently, astegolimab, an inhibitor of IL1RL1, a Tier 3 target, has completed phase 2 trials for eczema and asthma; in our study, we find genetic support where increased IL1RL1 increased risk of eczema in East Asian ancestry (**Fig. 6e**) and was concordant in European ancestries (**Supplementary Table 17**). Druggability visualization for remaining diseases for European and East Asian ancestries are provided in **Supplementary Note 7C**.

**Figure 6.**
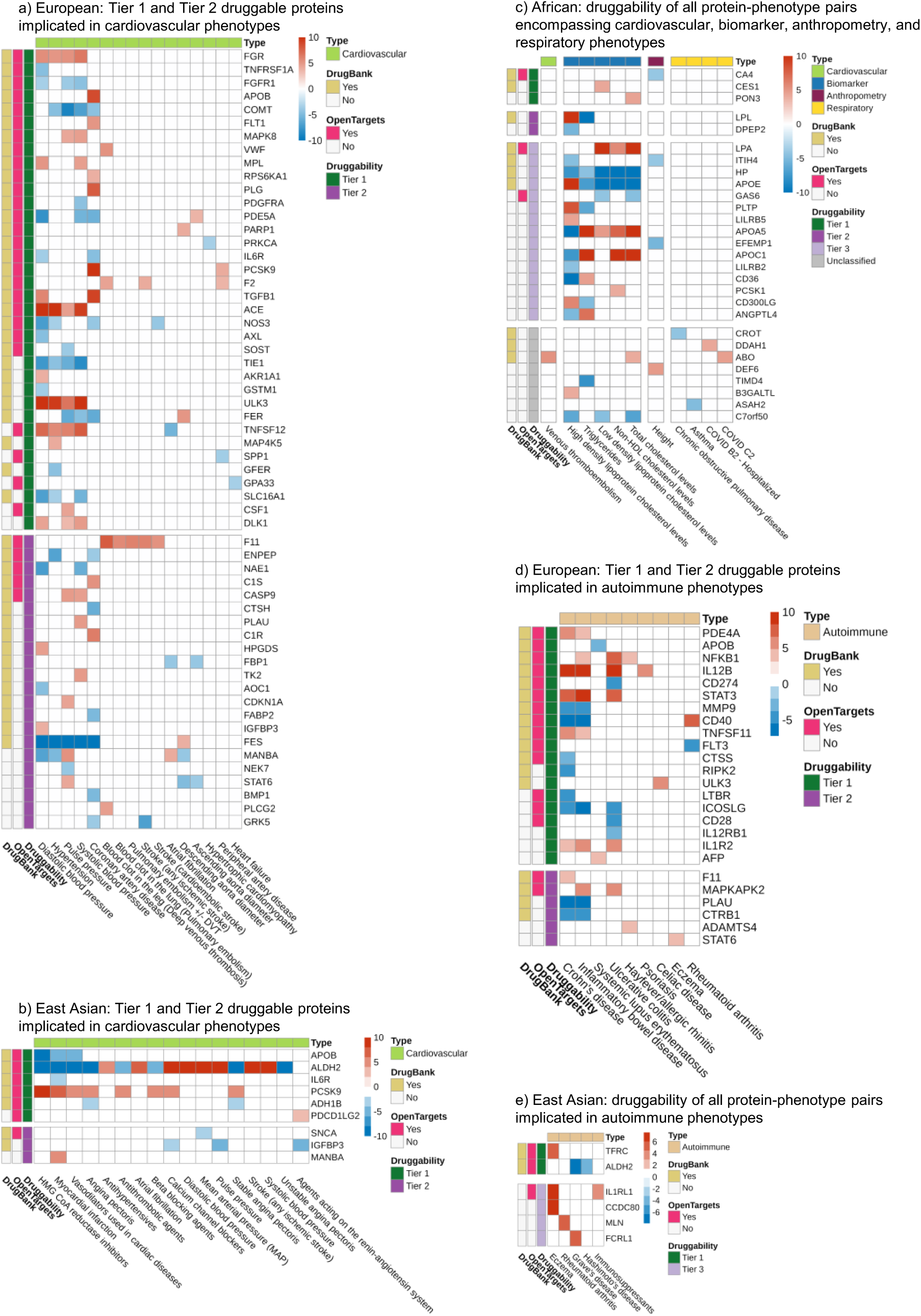
Integrating proteome-phenome-wide MR with the druggable genome, DrugBank, and clinical trial information from the Open Targets Platform. Legend information: Each cell displays a putatively causal protein-phenotype association. Cell color displays the MR effect estimate based on Z score averaged across cohorts capped at -10 to +10 with red showing a positive Z score indicating a positive MR effect of the protein (displayed as the gene name) on the phenotype and blue showing a negative Z score indicating a negative MR effect of the protein on the phenotype. For simplicity, in European ancestries, we only display protein-phenotypes with consistent effect across European cohorts. The y-axis shows the three drug databases. DrugBank (yellow square): DrugBank^39^ shows whether the protein has an available drug in the database. Open Targets (pink square): Open Targets Platform^40^ shows whether the protein has available clinical trial information. Druggability: The druggable genome from Finan et al.^38^ is shown for Tiers 1 (dark green, representing direct targets of approved small molecules and biotherapeutic drugs), Tier 2 (dark purple, representing proteins closely related to approved drug targets or which have associated drug-like compounds), Tier 3 (light purple, representing secreted or extracellular proteins, those distantly related to approved drug targets, and members of important druggable gene families not covered in Tier 1 or Tier 2), and Unclassified (gray, all other proteins not in Tiers 1 to 3). Proteins on the y-axis within each tier are sorted based on the number of supported databases. (a) European ancestry putatively causal protein-phenotype pairs stratified to Tier 1 and Tier 2 druggable proteins and cardiovascular phenotypes. (b) East Asian ancestry putatively causal protein-phenotype pairs stratified to Tier 1 and Tier 2 druggable proteins and cardiovascular phenotypes. (c) All of the African ancestry putatively causal protein-phenotype pairs with no druggability stratification. (d) European ancestry putatively causal protein-phenotype pairs stratified to Tier 1 and Tier 2 druggable proteins and autoimmune phenotypes. (e) East Asian ancestry putatively causal protein-phenotype pairs stratified to autoimmune phenotypes.

### 8. Converging evidence of the causal effect of IL1RL1 on IBD

Upon further stringent filtering to find protein-phenotype pairs with strong evidence (see **Methods**), we found that increased IL1RL1 levels was associated with increased risk of IBD, CD, and UC in European ancestries across three cohorts for IBD and CD and two cohorts for UC with consistent directions, which we validated using the largest available East Asian ancestry GWAS for IBD (14,393 cases and 15,456 controls), CD (7,372 cases and 15,456 controls), and UC (6,862 cases and 15,456 controls) from Liu et al.^65^ (**Fig. 7a**). In African ancestries, the number of cases were limited in the largest publicly available GWAS for IBD (1,285 cases and 119,314 controls)^57^ and UC (857 cases and 119,909 controls)^57^, thus estimates were not significant likely due to insufficient power.

**Figure 7.**
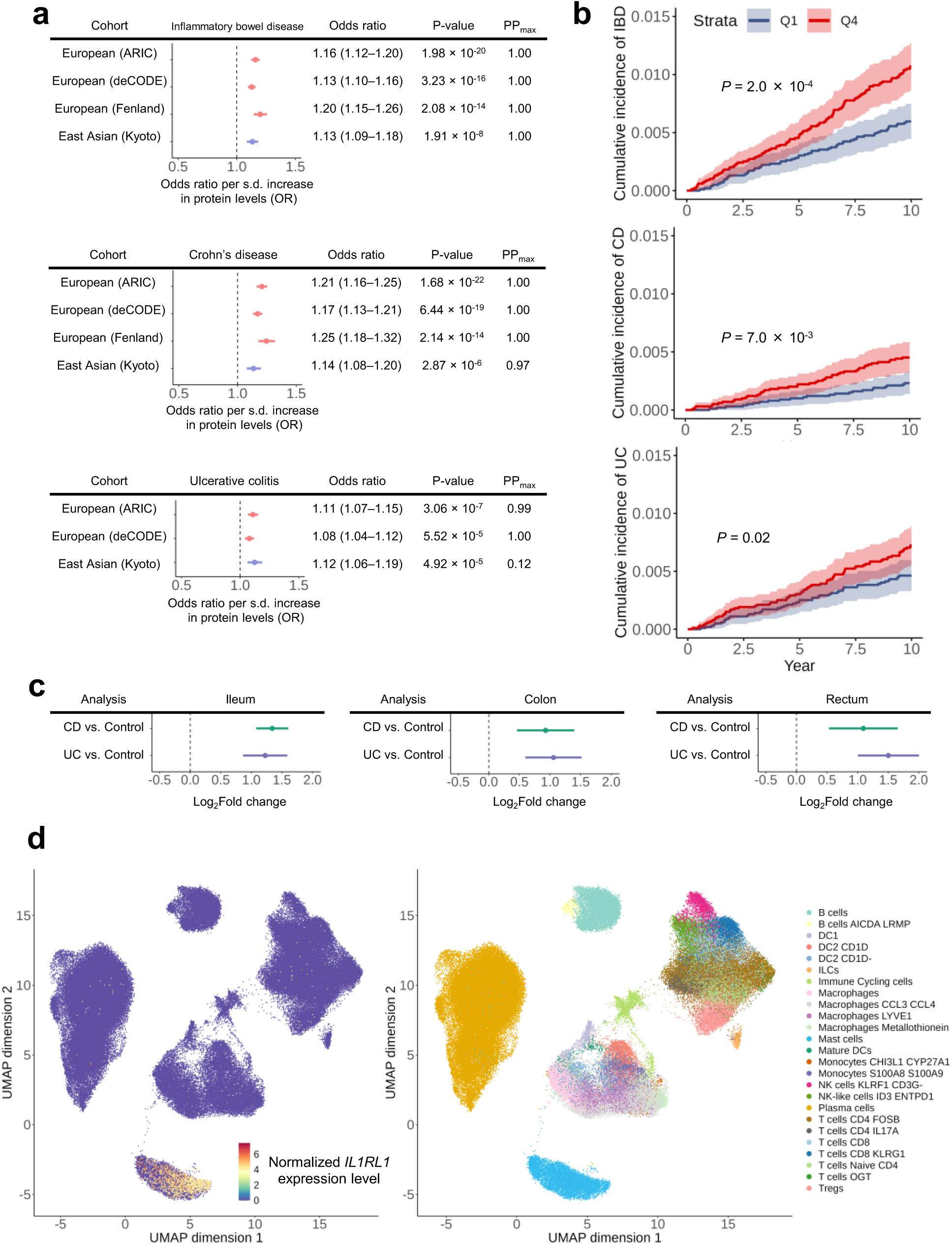
IL1RL1 in IBD, CD, and UC. a) MR for the effect of IL1RL1 and IBD (top), CD (middle), and UC (bottom) in European and East Asian ancestries. PP_max_ is the maximum colocalization posterior probability between PWCoCo and SharePro. b) Kaplan Meier estimates for cumulative incident of IBD (top), CD (middle), and UC (bottom) by baseline IL1RL1 level quantiles in the UK Biobank. *P* values were computed using the log-rank test. c) Bulk RNA sequencing of IL1RL1 in the ileum (left), colon (middle), and rectum (right). d) Single-cell RNA sequencing analyses of *IL1RL1*. *IL1RL1* expression patterns showing 720,633 cells collected from the terminal ileum and colon of 71 donors with different levels of inflammation. Single-cell transcriptomic data was obtained from Kong et al.^66^ (SCP1884 https://singlecell.broadinstitute.org/).

To triangulate the evidence, we performed supplementary observational analysis in the UK Biobank using Cox proportional hazards models. We adjusted for age, sex, recruitment center, Olink measurement batch, Olink processing time, and the first 10 genetic principal components. Notably, none of the genetic principal components were significant, suggesting that the association is not specific to ancestry. Over 10 years of follow-up, a one s.d. increase in IL1RL1 was associated with elevated risk of IBD (hazard ratio, HR = 1.18; 95% CI: 1.05–1.33; *P* = 6.6 × 10^-3^), CD (HR = 1.21; 95% CI: 1.00–1.47; *P* = 0.047), and UC (HR = 1.15; 95% CI: 1.00–1.33; *P* = 0.048), consistent with MR findings (**Supplementary Table 27**). Kaplan-Meier estimates for cumulative incidence of disease stratified by baseline IL1RL1 level (lowest 25% versus highest 25% in the UK Biobank population) also showed differences in IBD (log-rank test *P* = 2.0 × 10^-4^), CD (log-rank test *P* = 7.0 × 10^-3^), and UC (log-rank test *P* = 0.02) (**Fig. 7b**). We also performed an alternative, less stringent filter and prioritized proteins involved in CAD and type 2 diabetes which we provide in **Supplementary Note 8**.

#### 8.1. *IL1RL1* expression analyses

To further assess the role of IL1RL1 in IBD, we used the IBD Transcriptome and Metatranscriptome Meta-Analysis (IBD TaMMA) platform^67^ to compare expression of *IL1RL1* transcripts between IBD patients and healthy controls in the ileum, colon, and rectum. We found significantly higher *IL1RL1* gene expression in all three tissues (**Fig. 7c**), suggesting that increased *IL1RL1* expression is a consistent feature of IBD regardless of the specific location within the gastrointestinal tract.

To gain further insights into the role of IL1RL in IBD, CD and UC, we analyzed single-cell *IL1RL1* expression in 720,633 cells from the terminal ileum and colon of 71 participants with different levels of inflammation status from Kong et al.^66^ (SCP1884 https://singlecell.broadinstitute.org/). In single-cell RNA sequencing, *IL1RL1* showed significant enrichment in mast cells compared to 24 other cell types (permutation *P* < 2.0 × 10^-4^) (**Fig. 7d**). Mast cells, key players in allergic reactions and inflammation and a key cell type involved in the pathogenesis of IBD^68,69^, may contribute to chronic IBD by releasing inflammatory mediators like histamine and cytokines when activated by IL1RL1 in inflamed tissues. These findings align with our MR analyses showing that increased IL1RL1 leads to increased risk of IBD, CD and UC.

## Discussion

In this study, we conducted comprehensive multi-ancestry proteome-phenome analyses across three ancestries. Using seven large proteomics cohorts including European, African, and East Asian ancestries, we analyzed 355 complex traits or diseases, and identified 3,949, 56, and 325 putative causal effects of protein abundance on diseases and traits, respectively. By integrating data from druggable genomes and drug databases, we prioritized potential protein targets for drug development. Our findings offer a comprehensive atlas of protein-phenotype associations and an evidence-based resource to support drug discovery and development, expand insight into disease, and highlight potential targets for therapeutic intervention.

Our study provides an updated map to earlier phenome-wide MR studies of the human plasma proteome on complex diseases which were either limited to European ancestries^18^ or considered only a few diseases in European and African ancestries^19^. The significance of incorporating multiple ancestries is underscored by our identification of several proteins that are uniquely instrumentable by each ancestry due to allele frequency differences. Specifically, we instrumented an additional 119 proteins exclusively in African ancestry, 17 in East Asian ancestry, and 19 shared between African and East Asian ancestries. Moreover, the finding that a significant proportion of population-specific genetic variants—68.5% in African and 72.2% in East Asian— have a MAF below 0.01 in European ancestries highlights the potential for missed genetic discoveries when studies focus on a single ancestry. Research on the genetic architecture across different ancestries reveals both commonalities and differences, influenced by evolutionary history, genetic diversity, and population-specific factors^26,30^. Thus, by including diverse African and East Asian ancestries in proteomic analyses, we were able to instrument more proteins by leveraging common genetic variants in these underrepresented populations which were mostly rare in European ancestries, enhancing the potential for novel discoveries and exemplifying the value of including non-European individuals for comprehensive and inclusive proteomic and genetic analyses.

The inclusion of African and East Asian ancestries allowed discovery of protein-phenotype associations. We emphasize that these findings were attributable to uniquely instrumentable proteins in each ancestry and do not necessarily indicate ancestry-specific biological mechanisms.

As an illustrative example of the value of our atlas, we found that increased circulating abundances of IL1RL1 was causal for IBD, CD, and UC in European and East Asian ancestries, which was supported by observational analyses and gene expression analyses. IL1RL1 could be a promising therapeutic approach for IBD, potentially reducing mast cell-driven inflammation. Potential drugs targeting IL1RL1 include astegolimab which has completed phase 2 trials for eczema and asthma, tozorakimab which neutralizes IL-33, the interleukin that binds IL1RL1 (ST2), and anakinra which targets IL-1, a closely related interleukin. However, further research is required to assess the safety and efficacy of potential IL1RL1 inhibition. Many such findings may exist, and this atlas may be used as a tool to facilitate the selection of targets during primary or pre-clinical drug development, exploring drug repurposing opportunities, and improve understanding of proteins implicated in complex traits and diseases.

Our study has several key strengths. We curated GWAS for a wide range of complex traits and diseases, decreasing the overlap and redundancy and increasing the power for discovery in MR. Second, our study included diverse proteomics cohorts, including African ancestry and a new East Asian ancestry cohort, the Kyoto University Nagahama cohort. This latter cohort, which used the SomaScan v4 platform, is the largest to date aside from the China Kadoorie Biobank^25,26^. Further, our study combined both ARIC SomaScan and UKB-PPP Olink African proteomics cohorts at a phenome-wide scale and to include three ancestries. Notably, we identified uniquely instrumentable proteins in African and East Asian ancestries that were not found in European ancestries, highlighting the value of including cohorts from diverse ancestral backgrounds. Third, we performed extensive stringent filtering on genetic instruments with strict V2G criteria. Fourth, we harnessed novel state-of-the-art colocalization methods to reduce the risk of confounding from LD while increasing the statistical power to support more protein-phenotype associations with colocalization evidence^37^.

This study has several limitations. First, while two separate proteomics cohorts were included for African ancestries, the number of outcomes considered was still limited. Additionally, African ancestry phenotypes were curated from various cohorts with potentially finer genetic architecture differences than controlled for by using continental ancestries, thereby potentially biasing our analyses. Second, differences in measurement units make direct comparison of MR effect size estimates for continuous outcomes difficult. Nevertheless, direction of effect should be robust to this limitation. Third, while we reduced the risk of horizontal pleiotropy by using strict V2G *cis*-pQTLs, this resulted in many proteins being instrumented by a single *cis*-pQTL, limiting the ability to perform MR sensitivity analyses. Nonetheless, we used robust colocalization methods to mitigate risk of reporting false positives. Fourth, although we used assays measuring nearly 5,000 proteins from SomaScan and 3,000 proteins from Olink, coverage is still limited with regard to the entire proteome. Lastly, while we analyzed three diverse ancestries, the sample sizes for both proteomic GWAS and outcome GWAS were much larger for European ancestry, leading to differences in the number of associations. Greater coverage of proteins and larger sample sizes in non-European ancestries are needed.

In conclusion, through integrative multi-ancestry plasma proteome-phenome MR and extensive sensitivity analyses, we provided a comprehensive atlas of protein-phenotype associations across three ancestries and highlighted the value of multi-ancestry inclusion, as illustrated by uniquely instrumentable proteins in non-European ancestries. This study serves as a valuable resource for understanding disease mechanisms and prioritizing potential new targets.

## Methods

### 1. Proteomics cohorts

We analyzed proteomics cohorts from three ancestries consisting of European (four cohorts: ARIC, deCODE, Fenland, and UKB-PPP), African (two cohorts: ARIC and UKB-PPP), and East Asian (one cohort: Kyoto University Nagahama East Asian cohort). All cohorts had proteomics measured on the aptamer-based SomaScan assay v4 except for the UKB-PPP study, which used the antibody-based Olink Explore 3072 platform.

#### 1.1. European ancestry cohorts

We analyzed the GWAS of protein levels in individuals of European ancestry using four different studies. Three of these four studies (ARIC, deCODE, and Fenland described below) had proteomics measurements from the aptamer-based SomaScan assay v4 from SomaLogic (Boulder, Colorado, USA). In brief, SomaScan assay v4 uses aptamers, which are single-stranded oligonucleotides that have specific binding affinities to protein targets and can measure up to 5,000 unique proteins. The UKB-PPP study had proteomics measurements from the antibody-based Olink Explore 3072 platform, which measures up to 3,000 proteins. Briefly, Olink (Uppsala, Sweden) uses proximity extension assay (PEA) technology, which detects proteins through the binding of two separate antibodies carrying complementary oligonucleotide tags, which hybridize to the protein target. We restricted our analyses to proteins encoded by autosomal genes and analyzed a list of 4,687 proteins from SomaLogic and 2,823 proteins from Olink.

##### 1.1.1. ARIC

The Atherosclerosis Risk in Communities (ARIC)^13^ measured protein levels from 9,084 American participants of European and African ancestries using the SomaScan assay v4. Of these participants, 4,657 plasma proteins were measured for 7,213 European American individuals.

##### 1.1.2. deCODE

The deCODE study^12^ provided 4,907 aptamers that measure 4,719 proteins in 35,559 Icelandic individuals of European ancestry using SomaScan assay v4.

##### 1.1.3. Fenland

The Fenland study^11^ measured 4,775 proteins in 10,708 individuals of European ancestry using SomaScan assay v4.

##### 1.1.4. UKB-PPP

The UK Biobank Pharma Proteomics Project (UKB-PPP)^14^ conducted proteomic profiling on 54,219 individuals of multiple genetic ancestries in the UK Biobank using the Olink Explore 3072 platform. From this cohort, 34,557 European individuals, each with 2,923 unique proteins measured, were utilized in the UKB-PPP as the discovery cohort, and we used this discovery cohort in our study.

#### 1.2. African ancestry cohorts

For African ancestries, we used GWAS of protein levels from two different studies (ARIC and UKB-PPP) measured on SomaScan assay v4 and Olink Explore 3072, respectively.

##### 1.2.1. ARIC

The ARIC cohort was previously described in the European cohort section and consists of 9,084 European American and African American individuals. Of these participants, 4,657 proteins from the SomaScan v4 assay were measured for 1,871 African American individuals.

##### 1.2.2. UKB-PPP

We used the African ancestry individuals from the UKB-PPP study, which consists of 931 individuals with 2,923 unique proteins measured with the Olink Explore 3072 platform.

#### 1.3. East Asian ancestry cohort

##### Kyoto University Nagahama cohort

The Nagahama Primary Prevention Cohort Project (Kyoto-Nagahama cohort) is a joint project between the Kyoto University Graduate School of Medicine and Nagahama City, Shiga Prefecture that involved 10,000 residents of Nagahama (https://w3.genome.med.kyoto-u.ac.jp/en/nagahama-project/). Data generation was performed at the Kyoto University Center for Genome Medicine, where 1,823 Japanese individuals of East Asian ancestry were whole genome-sequenced and had 4,196 proteins measured using the SomaScan assay v4. Further details can be found in **Supplementary Note 9**.

### 2. Identification of strict variant-to-gene (v2g) *cis*-protein quantitative trait loci (pQTLs)

Each cohort had had different *cis*-pQTL definitions. For example, the ARIC^13^ study defined *cis*-pQTL as those within ±500 kb of the TSS of the protein-coding gene with FDR < 5%. The deCODE study^12^ defined *cis*-pQTL as those within ±1 Mb of the TSS of the protein-coding gene with *P* < 1.8 × 10^−9^. The Fenland Study^11^ defined *cis*-pQTL as those within ±500 kb of the protein-coding gene with *P* < 1.004 × 10^-11^. The UKB-PPP^14^ noted that of their identified pQTLs, 66.9% of proteins tested (1,954 of 2,922 proteins) had a *cis*-pQTL within ±1 Mb of the protein-coding gene with *P* < 1.7 × 10^−11^. Thus, we created a common *cis* definition as follows.

#### 2.1. Linkage disequilibrium (LD) clumping

We performed LD clumping (clumping window of 1 Mb, significance level of 5 × 10^−8^, and clumping *r*^2^ threshold of 0.001) on each proteomic GWAS in each ancestry cohort. For European proteomic GWAS, we used a reference panel composed of 50,000 randomly sampled unrelated UK Biobank^70^ individuals of European ancestry (UKB 50k). For African proteomic GWAS, we curated an LD reference panel from the Human Genome Diversity Project and 1000 Genomes Project (HGDP + 1kGP) reference panel^71^ for 994 African ancestry individuals. In East Asian ancestries, we used the 1000 Genomes East Asian (1kGP EAS) reference panel. We retained variants with a MAF > 0.01 in all reference panels.

#### 2.2. Identification of cis- and trans-pQTLs

To determine *cis*-pQTLs, we used the Ensembl BioMart^72^ package version Ensembl 105: Dec 2021 (Genome Reference Consortium Human Build 38, GRCh38.p13) to generate a human protein-coding genes file. We considered relevant attributes such as the canonical TSS, gene start, and gene end. Since Fenland proteomic GWAS were in GRCh37 coordinates, we separately curated a protein-coding genes file using BioMart on the GRCh37 assembly in Ensembl using version 110. For analysis, we excluded protein-coding genes located on sex chromosomes and those located within the major histocompatibility complex (MHC) (GRCh38: chr6 28,510,020– 33,480,577) due to the complex LD structure, high allelic diversity, and strong pleiotropy in this region^73^. We defined a *cis*-pQTL as a pQTL within ± 500 kb of the TSS of the protein-coding gene. All other pQTLs were *trans*. We used *cis*-pQTLs since they are more likely to directly impact the transcription and translation of the protein of interest.

#### 2.3. Strict variant-to-gene cis-pQTL definition

To minimize potential horizontal pleiotropic effects, we defined a unique strict V2G definition for *cis*-pQTLs whereby a *cis*-pQTL is a *strict V2G cis-pQTL* if it is a *cis*-pQTL for only one protein-coding gene (*strict*) and it has the strongest link to the corresponding protein-coding gene based on multiple sources of evidence and concomitantly has the highest Open Targets Genetics V2G score (**Extended Data Fig. 1**). We outline these steps in the following two sections.

##### 2.3.1. Strict cis-pQTL

For genome-wide significant independent *cis*-pQTLs in each cohort, we retained those associated with a single protein-coding gene. We defined this as a “strict” *cis*-pQTL definition. Here, we used *protein-coding genes* instead of *proteins* (*aptamers*) for the strict *cis* definition due to SomaScan assay v4 having multiple instances where two or more aptamers target a single protein, which would result in the unwarranted scenario where pQTLs that were *cis* for more than one aptamer of the same protein were removed.

##### 2.3.2. Open Targets Genetics Variant-to-Gene (V2G)

We used the Open Targets Genetics^35^ database (https://genetics.opentargets.org/) to determine whether each strict *cis*-pQTL held the highest V2G score for its corresponding associated protein-coding gene. This ensures that the variant is a suitable proxy for the plasma levels of the protein-coding gene. For instance, the variant may directly impact the protein-coding gene, potentially by altering its transcription, thereby influencing its plasma abundance. Briefly, Open Targets Genetics V2G scores are generated through a model trained on molecular QTLs (eQTL, sQTL, pQTL), chromatin interaction experiments such as promoter capture Hi-C (PCHi-C), *in silico* functional predictions such as Ensembl Variant Effect Predictor (VEP), and the distance between variants and genes’ canonical transcription start sites. This composition of evidence enables accurate assignment of variants to genes. We note that the pQTL datasets utilized by Open Targets Genetics for model training are from earlier studies and do not overlap with the proteomics cohorts we analyzed in this study, mitigating the risk of overfitting.

Together, we combined the strict *cis* definition and V2G score from Open Targets Genetics to curate strict V2G *cis*-pQTLs, which decreases the chance of horizontal pleiotropy in MR.

### 3. Protein altering variant (PAV) and expression quantitative trait loci (eQTL)

Since pQTLs altering the binding epitope of a protein may reflect assay specificity instead of the true biological function of the protein^1,64,74^, we annotated strict V2G *cis*-pQTLs as protein-altering variants (PAVs) of moderate or high impact if they or any variants in high LD (r^2^ > 0.8) were identified as PAVs using VEP^75^ (**Supplementary Tables 2–8**). VEP annotates the impact of variants into four categories: Modifier, Low, Moderate, or High (https://useast.ensembl.org/info/genome/variation/prediction/predicted_data.html). “Modifier” impact refers to variants in non-coding regions or affecting non-coding genes in which evidence of impact is hard to predict or limited. “Low” impact refers to variants that may not change protein behavior. “Moderate” impact variants are non-disruptive and may change protein effectiveness and include missense variants. “High” impact variants are disruptive and can cause truncation of proteins, loss of function, or trigger nonsense-mediated decay. If a strict V2G *cis*-pQTL and all of its LD proxies were labelled as Modifier or Low impact, we considered this strict V2G *cis*-pQTL to have no PAV. If a strict V2G *cis*-pQTL or any of its LD proxies were labeled Moderate or High, we considered this strict V2G *cis*-pQTL to be a PAV of -Moderate or -High impact, respectively.

We also conducted a *cis*-eQTL enrichment analysis using 49 tissues from GTEx v8 in European ancestries since if a *cis*-pQTL overlaps with the *cis*-eQTL of the same gene, it strengthens the evidence that the *cis*-pQTL acts directly on the gene products, reducing the risk of horizontal pleiotropy. To do so, we determined whether the strict V2G *cis-*pQTL was a *cis*-eQTL for the protein-coding gene of interest with *P* < 1 × 10^-5^ by querying 49 tissues in GTEx v8 European eQTLs. Querying was performed based on CHR:POS:EA:NEA and CHR:POS:NEA:EA (CHR: chromosome; POS: position; EA: effect allele; NEA: non-effect or other allele). In order to compare effect sizes across different cohorts, we aligned the strict V2G *cis*-pQTL effect allele in each cohort to the corresponding ancestry-specific reference panel’s alternative allele (UKB 50k for European, HGDP + 1kGP for African, and 1kGP EAS for East Asian ancestries). We note that pQTLs in the Fenland cohort were based on GRCh37 coordinates and were lifted to GRCh38 prior to querying for *cis*-eQTLs across GTEx v8 tissues. After synchronizing the genome assembly, 7 out of 3,045 strict V2G *cis*-pQTLs in Fenland were automatically labeled as having no eQTL since no corresponding GRCh38 coordinate existed for these variants, and 3,038 *cis*-pQTLs were analyzed.

### 4. GWAS outcome curation and selection

We manually curated the latest and largest GWAS (as of February 2024) for European and African ancestry outcomes. East Asian ancestry outcomes were selected from BioBank Japan^15^. All GWAS in GRCh37 were lifted to GRCh38 with the liftOver tool. We outline the curation steps for each ancestry’s GWAS outcomes in detail:

#### 4.1. European ancestry outcomes

During the curation, we considered 510 outcomes downloaded from 50 studies and one database. We removed outcomes that were duplicated and had a larger GWAS available, had ambiguous or broad definitions, were likely heterogeneous, were sex-specific, had missing relevant columns, had no download link available, and were not relevant to our outcomes of interest. We retained 179 outcomes for analysis (**Supplementary Table 10**). We labeled rsids, chromosome, and position if they were missing. Cases and sample sizes for each outcome were manually extracted from the original manuscript or the supplementary tables of each corresponding study, and we further categorized each outcome into one of 23 “Type” categories pertaining to the human system the outcome was based on or most likely to fall under.

#### 4.2. African ancestry outcomes

We manually curated 26 of the most up-to-date and publicly available African ancestry GWAS summary statistics (**Supplementary Table 11**). Restricted access GWAS from dbGap were not considered due to data access difficulties. Missing sample sizes were manually annotated with the sample size by inspecting the original manuscript and Supplementary Tables, while missing rsids were labeled using the HGDP + 1kGP reference panel. We annotated missing chromosomes and positions and used VEP to annotate variants missing effect allele frequency with gnomADg_AFR_AF (gnomAD genomes for African/American populations). We categorized each outcome into one of 8 “Type” categories, including Respiratory, Musculoskeletal, Cardiovascular, Eye, Anthropometry, Biomarker, Psychiatric, and Metabolic/endocrine.

As exploratory analyses, we included 114 binary cardiovascular and 9 binary autoimmune-related outcomes from the Million Veteran Program (*n* = 635,969) for African individuals.

#### 4.3. East Asian ancestry outcomes

We curated 220 outcomes from Biobank Japan (https://pheweb.jp/)^27^. We excluded 14 sex-specific outcomes, including Abortion, Breast cancer, Cervical cancer, Cesarian section, Ectopic pregnancy, Endometriosis, Endometrial cancer, Mastopathy, Ovarian cancer, Ovarian cyst, Pre-eclampsia, Prostate cancer, Uterine fibroid, and Uterine prolapse. We analyzed 206 outcomes (**Supplementary Table 12**). We also provide “Type” labels for each outcome, denoting the human system the outcome was based on or was most likely related to.

### 5. Two-sample Mendelian randomization

To assess the putative causal effect of protein abundance on outcomes in European, African, and East Asian ancestries, we performed two-sample MR using TwoSampleMR v.0.5.7^76^. To mitigate horizontal pleiotropy, we used strict V2G *cis*-pQTLs as instrumental variables to proxy protein-level exposures, as defined in the earlier sections of the methods. We harmonized exposure and outcome GWAS using the harmonise_data() function and performed a proxy search if an instrument was absent in the outcome GWAS. For European, African, and East Asian ancestries, we used the UKB 50k, HGDP + 1kGP, and 1kGP East Asian reference panels that were previously used for LD clumping for the proxy search, respectively. We searched for proxies using PLINK v.1.9^77^ parameters --ld-window=5000, --ld-window-kb=5000, --ld-window-r2=0.8 and retained proxies with minor allele frequencies ≤ 0.42.

MR analyses were performed using the mr() function. For proteins with a single genetic instrument, the association between the protein and outcome was evaluated using a Wald ratio estimate. For proteins with ≥ 2 genetic instruments, we used an inverse variance weighted random effects estimate. We determined whether genetic instrumental variables had F-statistics > 10^43,78^, indicating strong associations with the exposure and thus less chance of weak instrument bias, which may bias the causal effect estimates towards the null in two-sample MR. F-statistics are shown in **Supplementary Table 9**. We corrected for multiple testing per cohort in each ancestry by applying a Benjamini-Hochberg-corrected *P* threshold (FDR)^45^ of 0.05 (5%) as done previously^19^.

We note that beta estimates from MR for continuous traits are not directly comparable across different outcomes because the units used in GWAS vary. For instance, some GWAS use clinical units, whereas others use standardized and/or residualized values.

### 6. MR sensitivity analyses

To increase the robustness of MR findings, we further filtered MR results based on multiple sensitivity analyses, including heterogeneity tests, MR using alternative approaches (including weighted median, weighted mode, MR-Egger), and Steiger directionality test^79^ to assess reverse causality. Following these filtering steps, we refer to retained protein-phenotype associations as “MR-passing”. The sensitivity analyses are described for proteins with ≥ 2 instruments and proteins with ≥ 3 instruments below:

#### 6.1. Sensitivity analyses for proteins with two or more instruments

The heterogeneity test was performed for proteins with ≥ 2 instruments and describes whether strict V2G *cis*-pQTLs of the same protein are likely to show comparable effects on the tested outcomes. For heterogeneity testing, we used the mr_heterogeneity() function to compute a heterogeneity *P* value (Q_pval), and we calculated *I*^2^ statistics using the “Isq()” function. If an association had an *I*^2^ threshold ≥ 0.5 and a heterogeneity *P* value (Q_pval) < 0.05, this indicated considerable heterogeneity.

#### 6.2. Sensitivity analyses for proteins with three or more instruments

For proteins with ≥ 3 genetic instruments, we performed additional sensitivity analyses with alternative MR methods such as MR weighted median, MR weighted mode, and MR-Egger methods, as well as Steiger directionality testing. To check the consistency of MR estimates, we required the MR estimate, as well as the sensitivity analyses estimates from the MR weighted median, MR weighted mode, and MR-Egger approaches, to all have the same sign. For directional pleiotropy, we used the mr_pleiotropy_test() function to perform the MR-Egger intercept test and considered a *P* value < 0.05 as a statistically significant deviation from the null and an indication of directional pleiotropy. We also performed Steiger filtering on all proteins (directionality_test() function). Any pQTLs that explain more variance in the outcome than in the exposure potentially indicate reverse causation and were removed from further analysis.

### 7. Colocalization of proteomic GWAS with outcome GWAS

To assess whether plasma protein levels share the same causal variant with GWAS outcomes, we employed two colocalization methods to ensure the robustness of our findings. We performed PWCoCo^18,36^ and SharePro^37^ for MR associations that passed all MR sensitivity analyses described in the previous section. PWCoCo and SharePro are recent methods allowing multiple independent associations to be assessed. Both improve on the original coloc method^80^, which was limited by the assumption that a single variant exists per GWAS, wherein the method only considers the strongest of these distinct association signals when multiple independent associations exist. A detailed description of both methods is provided in **Supplementary Note 10**. Colocalization analyses were performed around a 1-Mb region centered on the lead (lowest *P* value) *cis*-pQTL. We set a colocalization posterior probability (PP) of a shared causal variant ≥ 0.8 in any of PWCoCo or SharePro as evidence of colocalization. For simplicity, we report the maximum PP between PWCoCo and SharePro (PP_max_) in the main text. We reported putatively causal associations (**Supplementary Tables 14–16**) as associations which pass all MR sensitivity analyses, Steiger filtering, and also colocalized with a PP ≥ 0.8 in any one of PWCoCo or SharePro. Due to the difficulty in verifying the corresponding Olink assay target for each SomaScan aptamer, we counted unique protein-phenotype associations using protein-coding genes to harmonize between proteomics platforms. **Supplementary Tables 14–16** contain MR and colocalization summaries, summaries of effect direction consistency across cohorts, flags of proteins instrumented by PAVs of high impact, and whether the protein-phenotype association came from a protein uniquely instrumentable in that ancestry. We also annotate whether protein-phenotype associations passed the most stringent Bonferroni correction for the total number of MR tests across three ancestries (*P* < 0.05 / 874,465).

### 8. Distinguishing between previously reported and unreported protein-phenotype pairs

#### 8.1. Comparing against earlier studies identifying putatively causal protein-phenotype pairs with MR and colocalization evidence

To identify the status of protein-phenotype associations as reported or unreported (not found from pre-existing proteome-phenome-wide MR studies) in European ancestries, we overlapped our associations with recent proteome-phenome-wide MR analyses from Zheng et al. 2020^18^ and multi-ancestry proteome-wide MR analyses from Zhao et al. 2022^19^. We used the 111 identified putatively causal associations (65 proteins on 52 phenotypes) from “Table S7” of Zheng et al.^18^ and the 45 associations from “ST7A” of Zhao et al.^19^.

We note that Zhao et al. used three colocalization methods and a more relaxed threshold of PP > 0.7 as evidence of colocalization. To do so, we harmonized the outcomes from Zheng and Zhao to our outcomes and matched protein-phenotype pairs using ensembl ID and outcome name for the Zheng study and UniProt ID and outcome name for the Zhao study. Any of our identified putatively causal European ancestry association pairs not from these two studies were identified as unreported.

For African ancestries, we overlapped our protein-phenotype pairs with the single protein-phenotype pair passing FDR correction identified in “ST8A” of Zhao et al^19^. Any pairs that did not overlap were considered unreported.

### 9. Protein-phenotype network plots

We used Python igraph (v.0.10.8) to generate networks. Placement of nodes was generated in Cytoscape (v.3.10.2) and manual editing was performed in Adobe Illustrator (v.28.2).

### 10. Effect concordance within-ancestry

To determine the concordance of MR effect estimates within European and African ancestries, which both included more than one proteomics cohort, we compared protein-coding genes to harmonize assay names across SomaScan assay v4 and Olink 3072 Explore platforms. However, we could not verify whether Olink assays for a particular protein targeted the same domain as its corresponding SomaScan assay, which may lead to differences in MR estimates. Moreover, since some proteins were instrumented by *cis*-pQTLs that may be PAVs of high impact, which could also lead to discordance in effects, we annotated these associations with a flag and advise caution in the interpretation of these flagged results (**Supplementary Tables 14–16**).

### 11. Druggability assessment

We performed a druggability assessment on instrumentable protein-coding genes and protein-phenotype associations that showed putatively causal relationships.

#### 11.1. Druggability assessment of instrumentable proteins

For instrumentable protein-coding genes, we determined druggability based on Finan et al.^38^, as described below.

#### 11.1.1. Finan et al

Finan et al.^38^ considered 20,300 protein-coding genes annotated using Ensembl version 73 and classified 4,479 (22%) into three tiers (Tier 1, 2, and 3) as drugged or druggable. Tier 1 (1,427 genes) encompasses the primary targets of approved small molecules and biotherapeutic drugs, along with those influenced by clinical-phase drug candidates. Tier 2 (682 gene) involves proteins closely associated with drug targets or linked to drug-like compounds. Meanwhile, Tier 3 (2,370 genes) comprises secreted or extracellular proteins that are distantly related to approved drug targets, and those in important druggable gene families not covered in Tiers 1 or 2. We denoted all other protein-coding genes that did not fall in Tier 1, 2, or 3 categories as “Unclassified”. To check the overlap of protein-coding genes across all cohorts when stratifying into Finan et al. tiers (**Supplementary** Figure 5 and 6), we used the UpSetR^81^ package v.1.4.0 (https://github.com/hms-dbmi/UpSetR).

### 11.2. Druggability assessment and enrichment of protein-phenotype pairs

We first assessed how many proteins from the identified pairs of putatively causal protein-phenotype associations had existing drugs by querying DrugBank^39^. We used Ensembl ID to match protein targets in DrugBank. Next, we created heatmaps using the pheatmap package v.1.0.12 in R incorporating the druggable genome, Drugbank, and Open Targets Platform (described below). We overlapped protein-phenotype pairs with the DrugBank database to determine whether any drugs existed for these disease-implicated proteins while Open Targets was used to determine whether any clinical trial information existed for these proteins.

#### 11.2.1. DrugBank

We used DrugBank database v.5.1.12 (https://go.drugbank.com/releases/latest) and R package dbparser v.2.0.2 to parse the DrugBank database xml file. We aggregated drugs and target information and overlapped this with putatively causal protein-phenotype associations to determine which proteins had an available drug.

#### 11.2.2. Open Targets

We used Open Targets v.24.03 (https://platform.opentargets.org/downloads). We used the knownDrugsAggregated dataset, which provides information on known drugs for a given disease and contains protein target information. Open Targets also includes information on clinical trials and its phases, which we used to determine the status of a protein target and its corresponding drug. We overlapped this dataset by matching Ensembl ID with our identified protein-phenotype associations.

### 12. Follow-up analyses showing evidence for IL1RL1

We filtered protein-phenotype associations in European ancestries by ensuring that the protein was instrumented in two or more ancestries, was targeted by both SomaScan and Olink assays, had an MR effect that was concordant across all cohorts, had MR and colocalization evidence in three or more cohorts for the protein-phenotype pair, and was implicated in at least one binary phenotype. Following these filtering steps, we identified 53 candidate protein-phenotype pairs and highlighted IL1RL1 in IBD, CD, and UC as an illustrative example.

#### 12.1. MR and colocalization of IL1RL1 in East Asian ancestry with IBD, CD, and UC

To validate that IL1RL1 was also putatively causal for IBD, CD, and UC in non-European ancestries, we performed two-sample MR and colocalization with PWCoCo and SharePro using the IL1RL1 strict V2G *cis*-pQTL (rs12712135) identified in the Kyoto University Nagahama East Asian ancestry proteomics cohort. We used the largest East Asian ancestry GWAS for IBD (14,393 cases and 15,456 controls), CD (7,372 cases and 15,456 controls), and UC (6,862 cases and 15,456 controls) from Liu et al.^65^. Harmonization was performed similarly to the primary analyses, and we used the Wald ratio to obtain MR effect estimates. Colocalization was performed as described in earlier sections of the methods, and we used PP ≥ 0.8 as the threshold for evidence of colocalization in any of PWCoCo or SharePro.

#### 12.2. MR of IL1RL1 in African ancestry with IBD and UC

We estimated the causal effect of IL1RL1 on IBD (1,285 cases and 119,314 controls) and UC (857 cases and 119,909 controls) in African ancestry using outcome GWAS from the Million Veteran Program^57^. We performed MR using the IL1RL1 strict V2G cis-pQTL (rs1420101) in the ARIC and UKB-PPP African ancestry cohorts. The Wald ratio was used to obtain MR effect estimates.

#### 12.3. Cox regression analysis for 10-year cumulative events of IBD, CD, and UC in the UK Biobank

We used multivariable Cox proportional hazards regression to determine whether baseline plasma IL1RL1 protein level was associated with cumulative events of IBD, CD, or UC. We adjusted for age, sex, recruitment center, Olink measurement batch, Olink processing time, and the first 10 genetic principal components (UKB field: 22009) to adjust for genetic ancestry while protein levels were rank-based inverse normal transformed. We used the coxph() function from the survival R package v.3.2.13 and considered *P* < 0.05 as nominal significance of association. None of the genetic principal components were significantly associated with the IBD, CD, and UC outcome in each analysis.

We checked the proportional hazards assumption using the cox.zph() function for the IL1RL1 association analysis for IBD, CD, and UC for the rank-based inverse normal transformed IL1RL1 covariate. The proportional hazards assumption tests the null hypothesis that each covariate’s effect estimate does not vary with time. We used the “GLOBAL” variable from cox.zph() which tests the null hypothesis of whether all inputted covariates meet the proportional hazards assumption. We considered *P* < 0.05 as evidence that the proportional hazards assumption was not fulfilled.

We defined IBD using ICD10 codes K50-K51, CD using K50, and UC using K51. We calculated the time to event by subtracting the date of event registration from the date of enrollment (data field: 53), focusing on events occurring within 10 years of enrollment. We excluded cases of prevalent IBD that met these criteria before enrollment, and those without a recorded event date for IBD, and performed the same steps for CD and UC. Controls for the IBD analysis were defined as individuals without an IBD, UC, or CD record based on self-reported medical history. Controls for the CD analysis were defined as individuals without a CD record, and controls for the UC analysis were defined as individuals without a UC record, based on self-reported medical history. We analyzed 333 cases and 40,001 controls for IBD, 130 cases and 40,388 controls for CD, and 240 cases and 40,178 controls for UC.

We plotted Kaplan-Meier curves by stratifying individuals into the bottom 25% and the top 25% based on baseline plasma IL1RL1 levels. We performed a log-rank test to assess whether there is a statistically significant difference in survival between these two groups, with a nominal *P* < 0.05.

#### 12.4. Bulk RNA-sequencing

We used the IBD Transcriptome and Metatranscriptome Meta-Analysis (IBD TaMMA) platform^67^ (https://ibd-meta-analysis.herokuapp.com) to evaluate changes in *IL1RL1* gene expression. IBD TaMMA encompasses 3,853 RNA-Seq datasets from 26 studies on IBD and control samples across various tissues. All datasets were processed using a uniform computational pipeline and underwent batch correction for harmonizing data, enabling consistent comparison across studies. Differential expression results for ileum, colon, and rectum biopsies from CD and UC patients versus healthy controls were downloaded from IBD TaMMA. We assessed the log_2_ fold change to create forest plots.

#### 12.5. Single-cell RNA-sequencing

To gain a better understanding of the enrichment of *IL1RL1* in specific cell types, we obtained single-cell RNA sequencing data from Kong et al.^66^ (SCP1884 from https://singlecell.broadinstitute.org/), which profiled 720,633 cells from the terminal ileum and colon of 71 CD individuals with different levels of inflammation. Specific details of sample collection, data processing, and single-cell profiling have been described previously^66^. We evaluated the normalized gene expression levels of *IL1RL1* in 25 different cell types and replotted the first two dimensions of Uniform Manifold Approximation and Projection (UMAP) coordinates to visualize the cell clusters. To determine if *IL1RL1* was more significantly expressed in certain cell types, we conducted 5,000 permutations of the cell type labels. We assessed how often a specific cell type had the same or a higher proportion of cells expressing *IL1RL1* compared to all the cells in the overall population (permutation *P* value).

### 13. STROBE-MR statement

Our study closely adheres to the STROBE-MR guidelines and the STROBE-MR checklist is attached in **Supplementary Note 11**.

### 14. Ethics declarations

All contributing cohorts obtained ethical approval from their institutional ethics review boards. The contributing proteomics cohorts include the Atherosclerosis Risk in Communities (ARIC) Study, deCODE study, Fenland study, UK Biobank, and Kyoto University Nagahama study. The UK Biobank has approval from the North West Multi-centre Research Ethics Committee as a Research Tissue Bank.

### 15. Data availability

We will provide unfiltered proteome-phenome wide MR results for European (ARIC, deCODE, Fenland, UKB-PPP), African (ARIC, UKB-PPP), and East Asian (Kyoto University Nagahama cohort) ancestries on FigShare upon publication. We caution against directly comparing MR effect estimates across continuous outcomes, as the outcomes were collected from various sources and may not be scaled to the same units.

#### 15.1. Proteomic GWAS

ARIC summary statistics (EUR and AFR): http://nilanjanchatterjeelab.org/pwas/

deCODE summary summary statistics (EUR): https://www.deCODE.com/summarydata/

Fenland summary statistics (EUR): https://omicscience.org/apps/pgwas/

UKB-PPP summary statistics (EUR, AFR): http://ukb-ppp.gwas.eu/

Kyoto University Nagahama cohort summary statistics (EAS): Available through contacting the authors of this study.

#### 15.2. Outcome GWAS summary statistics

Information on the 179 European outcomes used in this study and the link to the original summary statistics is available in **Supplementary Table 10**.

The 26 African outcomes are available in **Supplementary Table 11**.

The 206 East Asian outcomes are available in **Supplementary Table 12**.

Million Veteran Program outcomes can be found in the original study^57^.

The largest East Asian ancestry IBD, CD, and UC GWAS are publicly available from Liu et al.^65^ and were downloaded from https://www.ibdgenetics.org/

#### 15.3. Variant-to-gene score

Open Targets Genetics^35^ (https://genetics-docs.opentargets.org/data-access/data-download),

#### 15.4. Reference panels

European ancestries: UKB 50k (https://www.ukbiobank.ac.uk/)

East Asian ancestries: 1000 Genomes Project (https://www.internationalgenome.org/data)

African ancestries: HGDP+1KG (https://gnomad.broadinstitute.org/news/2020-10-gnomad-v3-1-new-content-methods-annotations-and-data-availability/#the-gnomad-hgdp-and-1000-genomes-callset)

#### 15.5. Druggability

We used Finan et al. 2017^38^ for the list of 4,479 protein-coding genes in each druggability tier, DrugBank database v.5.1.12 (https://go.drugbank.com/releases/latest) for information on drugs targeting specific proteins, and Open Targets Platform database v.24.03 (https://platform.opentargets.org/downloads) for clinical trial phase and status information for protein-drug-disease triplets.

#### 15.6. Expression analyses

For gene expression data, we used data from Kong et al.^66^ (SCP1884 at Single Cell Portal https://singlecell.broadinstitute.org/).

### 16. Code availability

We used R v.4.1.2 (https://www.r-project.org/),

Python 3.10 (https://www.python.org/downloads/release/python-3100/)

PLINK v.1.9^77^ (http://pngu.mgh.harvard.edu/purcell/plink/),

TwoSampleMR v.0.5.6 (https://mrcieu.github.io/TwoSampleMR/),

coloc v.5.2.3^80^ (https://chr1swallace.github.io/coloc/),

PWCoCo^18,36^ (https://github.com/jwr-git/pwcoco),

SharePro v.5.0.0^37^ (https://github.com/zhwm/SharePro_coloc/),

Cytoscape v.3.10.2 (https://cytoscape.org/),

LocusZoom^82^ (https://my.locuszoom.org/)

Code used in this study will be made available at https://github.com/chenyangsu/pQTL-MR upon publication.

## 17. Acknowledgments

This research has been conducted using the UK Biobank Resource under Application Number 27449. C.-Y.S. is supported by a CIHR Canada Graduate Scholarship Doctoral Award (Funding Reference Number: 187673), an FRQS doctoral training scholarship, and a Lady Davis Institute/TD-Bank Scholarship. T.L. has been supported by start-up funding from the Office of the Vice Chancellor for Research and Graduate Education, School of Medicine and Public Health, and Department of Population Health Sciences at the University of Wisconsin-Madison. S.Y. is supported by the Japan Society for the Promotion of Science.

The funders had no role in the study design, data collection and analysis, decision to publish, or preparation of the manuscript. We acknowledge Servier Medical Art (https://smart.servier.com/) for providing images that were used to create diagrams in this study. We thank the Digital Research Alliance of Canada for providing computing resources.

## 18. Author contributions

Conception and design: C.-Y.S., T.L., S.Y. Methodology: C.-Y.S., W. Z., T.L., S.Y.

Data curation: C.-Y.S., S.S.-H., T.-Y.Y., K.Y.H.L., Y.C., F. M., T.L., S.Y.

Data Analysis: C.-Y.S., T.L., S.Y. Visualization: C.-Y.S., A.v.d.G., T.L., S.Y.

Knowledge portal: C.-Y.S., D.-K.J., M.C., S.Y. Writing—Original Draft: C.-Y.S.

Writing—Review and Editing: all authors Supervision: T.L., S.Y.

Project administration: T.L., S.Y.

Funding acquisition: V.M., S.Z., T.L., S.Y.

## Extended Data Figures

**Extended Data Fig. 1.**
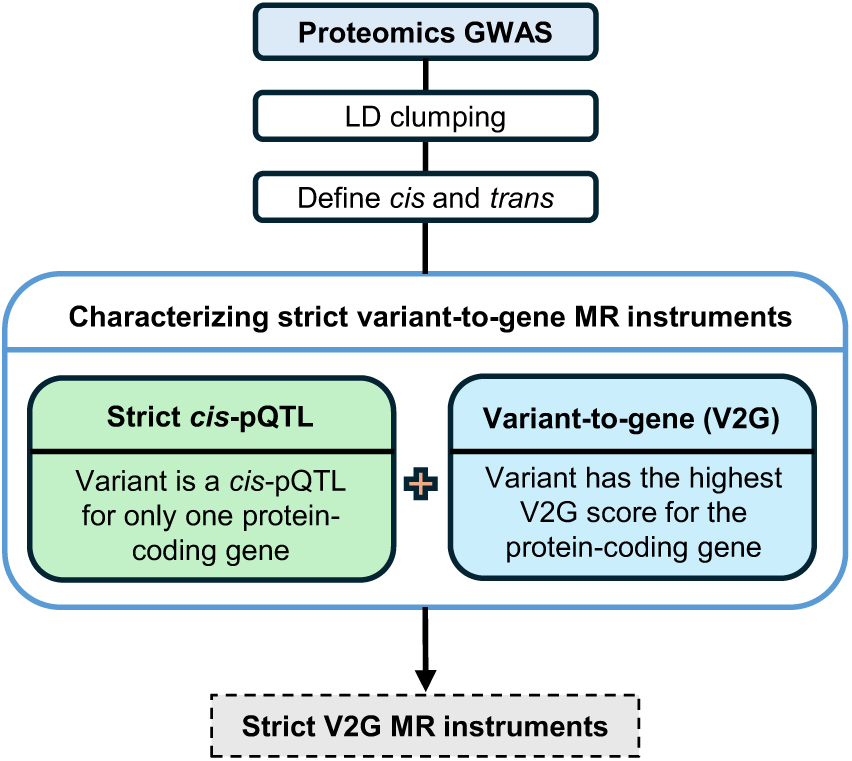
Flow diagram showing the definition of strict variant-to-gene *cis*-pQTLs. Flow diagram showing the selection of strict variant-to-gene (V2G) *cis*-pQTLs used as instruments for MR starting from the proteomic GWAS.

**Extended Data Fig. 2.**
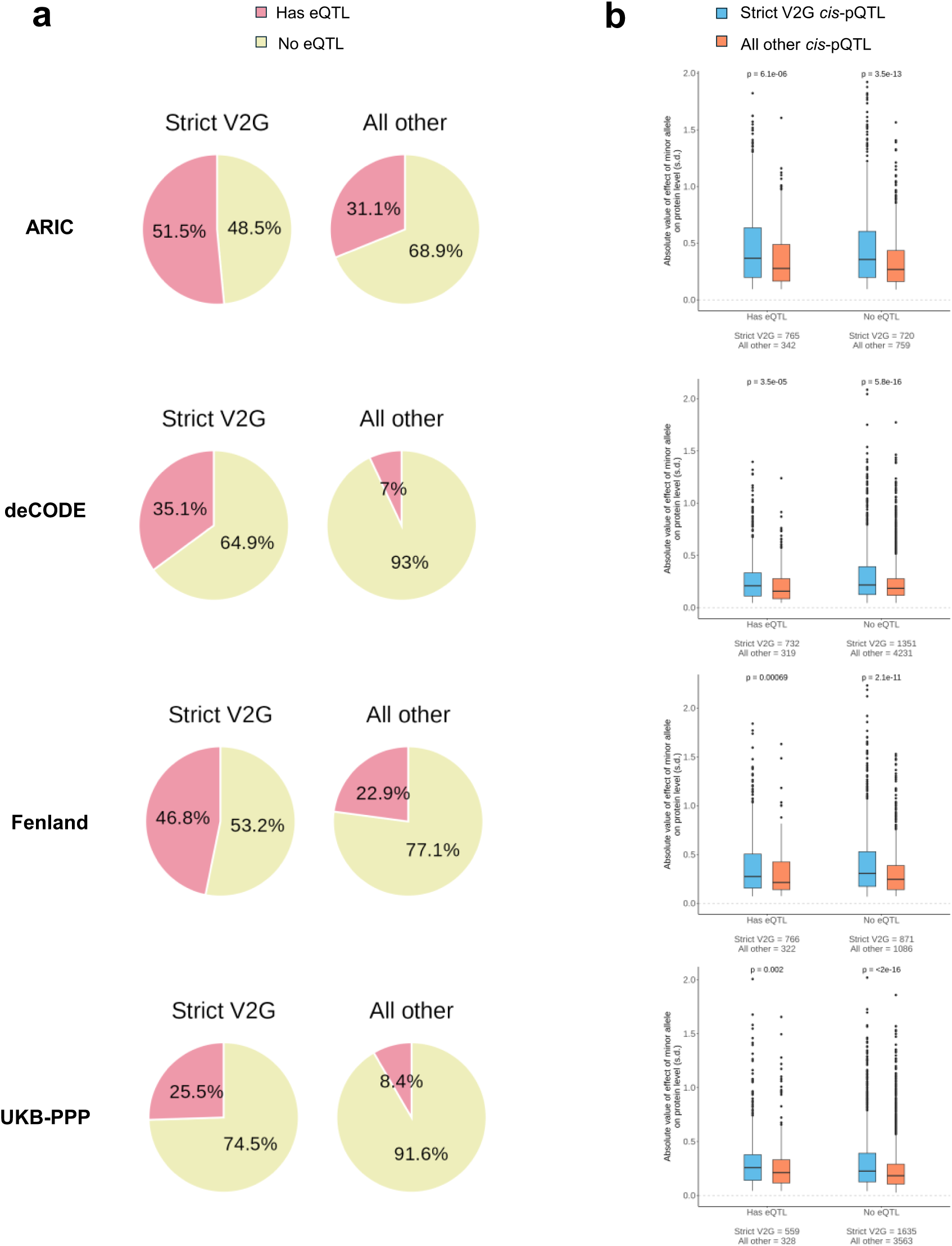
eQTL enrichment analysis comparing strict V2G *cis*-pQTLs against all other *cis*-pQTLs. (a) Pie charts showing proportion of eQTL enrichment in the ARIC, deCODE, Fenland, and UKB-PPP European ancestry cohorts for strict V2G *cis*-pQTLs compared to all other *cis*-pQTLs. Red: presence of a *cis*-eQTL; Yellow: absence of a *cis*-eQTL. (b) Absolute value of effect of the minor allele on protein level broken down by the presence or absence of *cis*-eQTLs in the ARIC, deCODE, Fenland, and UKB-PPP European ancestry cohorts. Strict V2G, strict variant-to-gene *cis*-pQTLs; All other, all other *cis*-pQTLs that were removed due to strict V2G filtering. Boxplots show the median, lower, and upper quartiles; whiskers end at 1.5 times the interquartile range from the top and bottom of the box; points outside the whisker boundaries are plotted individually; smaller black dots represent individual points and are used to show the number of samples included in each boxplot; significance level *P* value is based on the Mann-Whitney U test.

**Extended Data Fig. 3.**
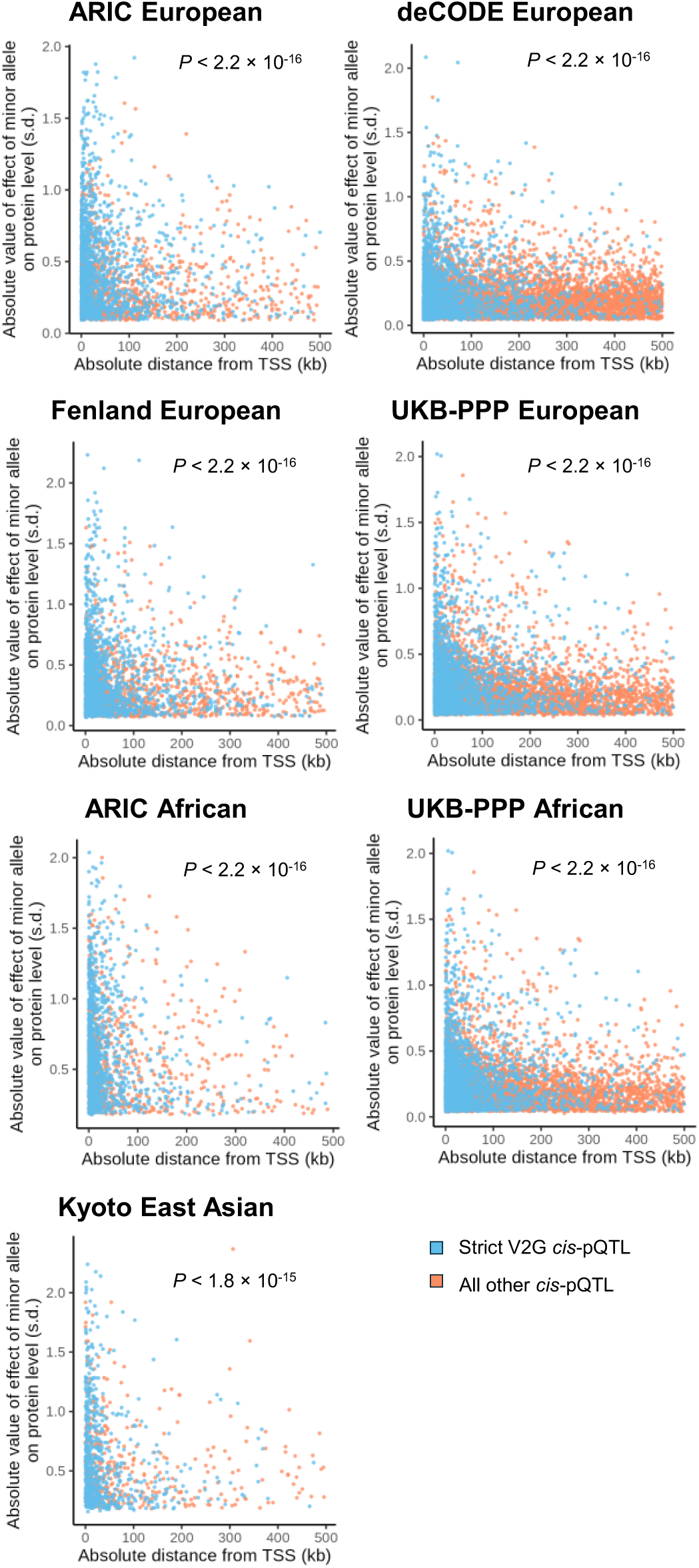
Absolute distance from transcription start site versus effect size for strict V2G *cis*-pQTLs and all other *cis*-pQTLs. The x-axis shows the distance of the pQTL from the canonical transcription start site of the associated protein-coding gene while the y-axis shows the absolute value of the effect size estimate of the effect allele aligned to the minor allele of each ancestry’s respective reference panel. (Note, in European, African, and East Asian ancestry proteomics cohorts, the effect allele of *cis*-pQTLs in each cohort was aligned to the minor allele of the corresponding variant in their respective reference panels—UKB 50k for European, HGDP+1kGP for African, and 1kGP for East Asian ancestry—to harmonize alleles across each ancestral cohort for plotting). Strict V2G *cis*-pQTLs are highlighted in blue while all other *cis*-pQTLs are highlighted in orange. Note that Fenland European *cis*-pQTLs are presented in GRCh37 coordinates while all other cohorts are presented in GRCh38 coordinates. *P* values show a one-sided t-test testing whether strict V2G *cis*-pQTLs have smaller absolute distance to the TSS compared to all other *cis*-pQTLs.

**Extended Data Fig. 4.**
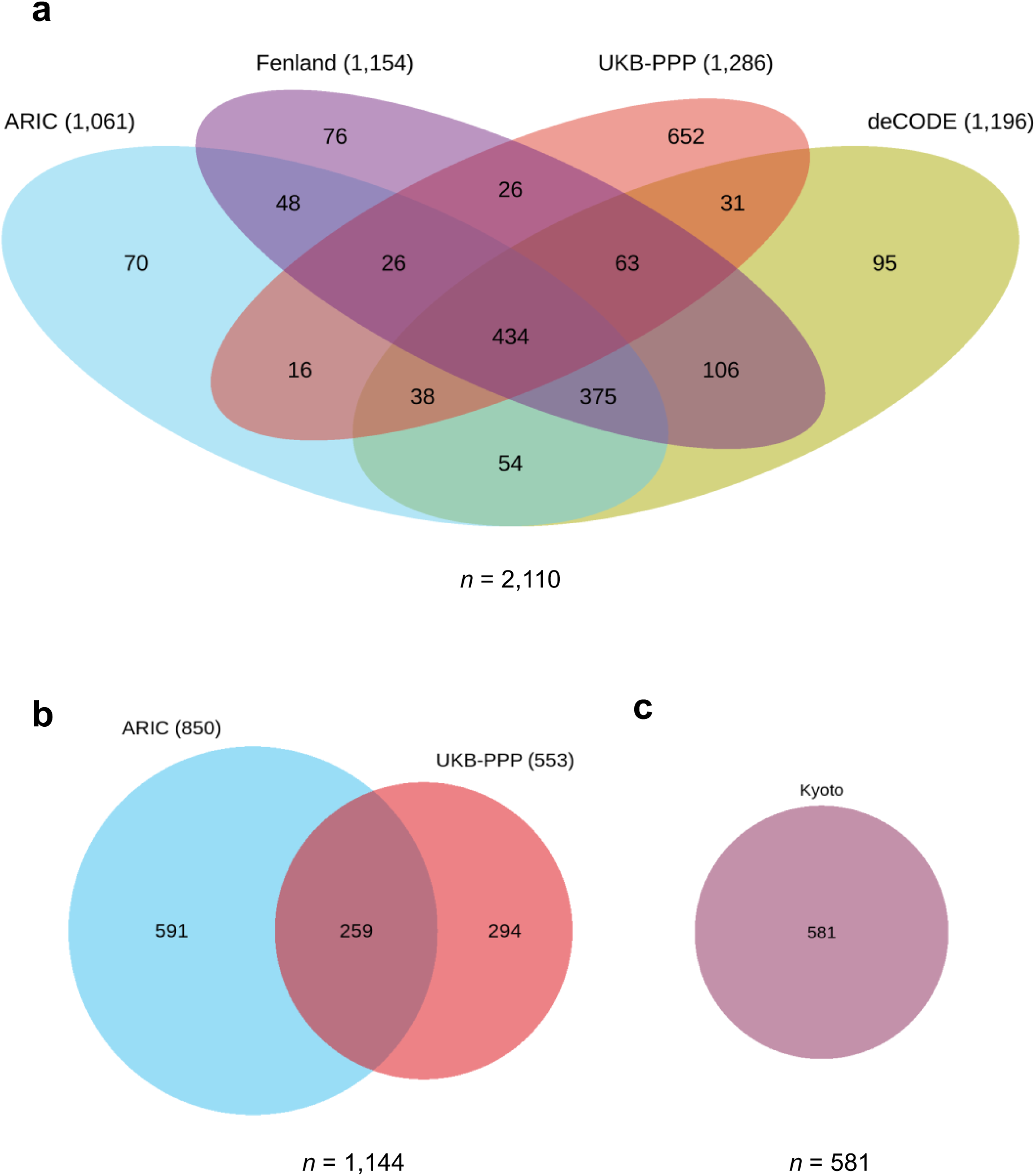
Within ancestry comparison of instrumentable proteins. Venn diagrams of overlapping instrumentable proteins within ancestries. (a) European cohorts involving ARIC, deCODE, Fenland, and UKB-PPP (4 cohorts). (b) African cohorts involving ARIC and UKB-PPP (2 cohorts). (c) East Asian ancestry cohort from Kyoto University Nagahama (single cohort).

**Extended Data Fig. 5.**
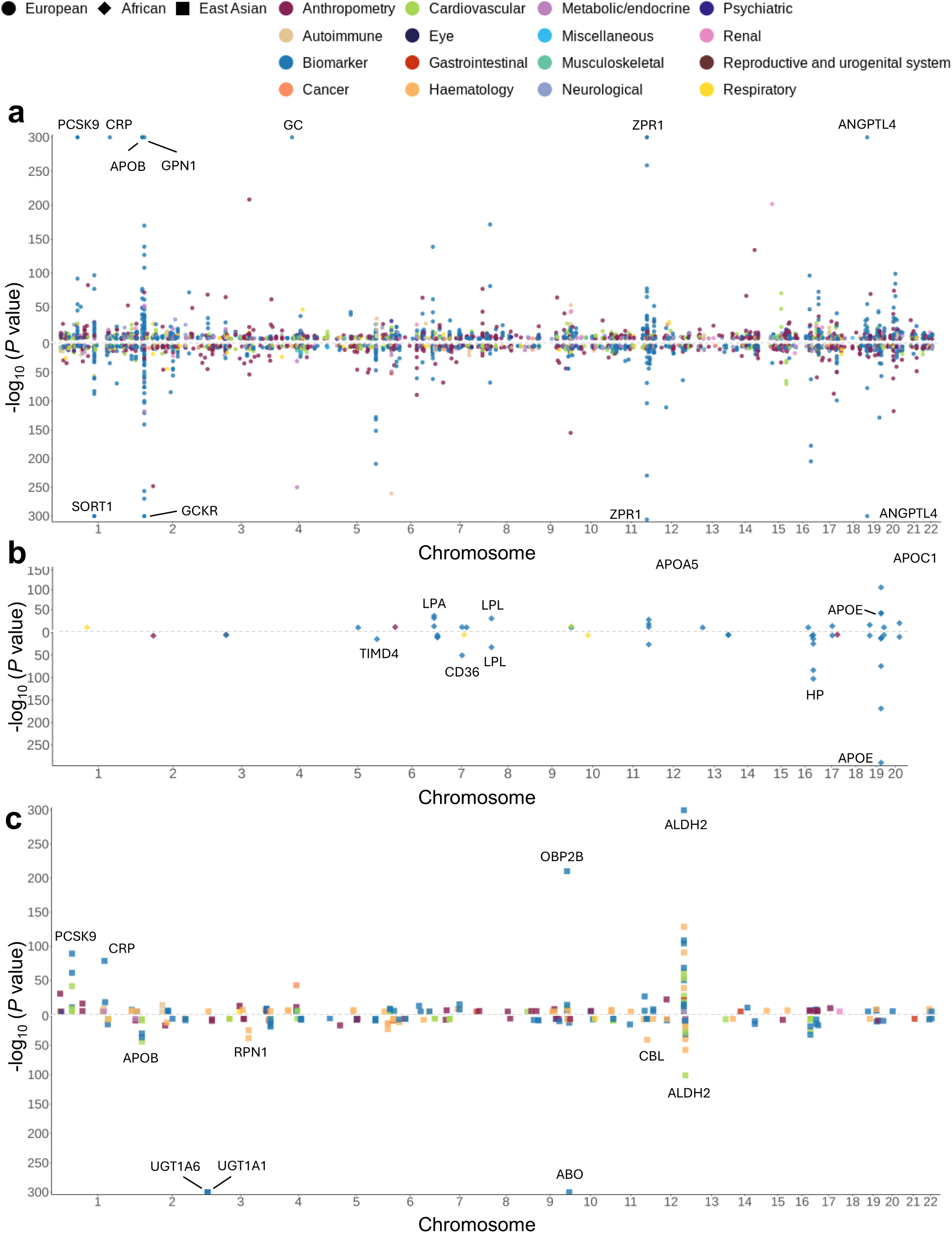
Putatively causal protein-phenotype associations across European, African, and East Asian ancestries. Miami plots displaying chromosomal position (*x*-axis) of significant putatively causal protein-outcome associations (MR-passing and colocalized with PP_max_ ≥ 0.8) in (a) European, (b) African, (c) East Asian ancestry. The *y*-axis shows *P* values from the MR causal estimates where the exposure is protein level and outcome is the complex trait or disease. Colors indicate the type of complex trait or disease. Cancer types were harmonized under a single “Cancer” group. Ancestry is denoted by filled circle (European), filled diamond (African), and filled square (East Asian). Each data point is plotted based on the chromosome and transcription start-site of the protein-coding gene. For simplicity, Z scores and *P* values are averaged across cohorts in the European ancestry plot and the African ancestry plot and shown as a single data point. In European ancestry, only associations that were consistent across all cohorts are shown.

**Extended Data Fig. 6.**
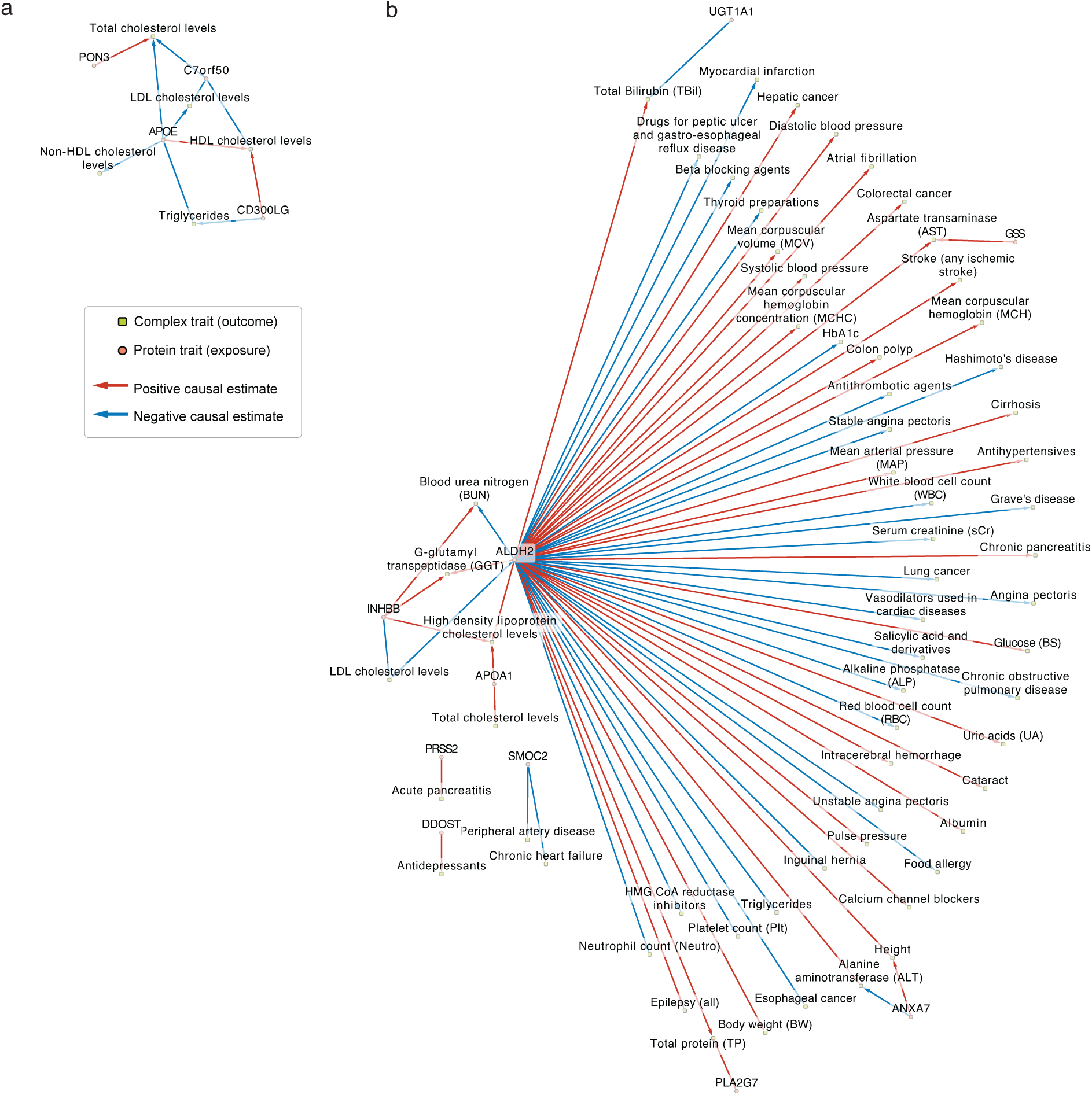
Uniquely instrumentable protein-phenotype pairs in African and East Asian ancestries. Red arrows indicate a positive causal estimate of the protein on the outcome while blue arrows indicate a negative causal estimate of the protein on the outcome. (a) Protein-phenotype pairs from 4 proteins uniquely instrumentable in African ancestry. Significant estimates between proteins (orange circles) and traits (green rectangles). (b) Protein-phenotype pairs from 8 proteins uniquely instrumentable in East Asian ancestry. Significant estimates between proteins (orange circles) and traits (green rectangles).

**Extended Data Fig. 7.**
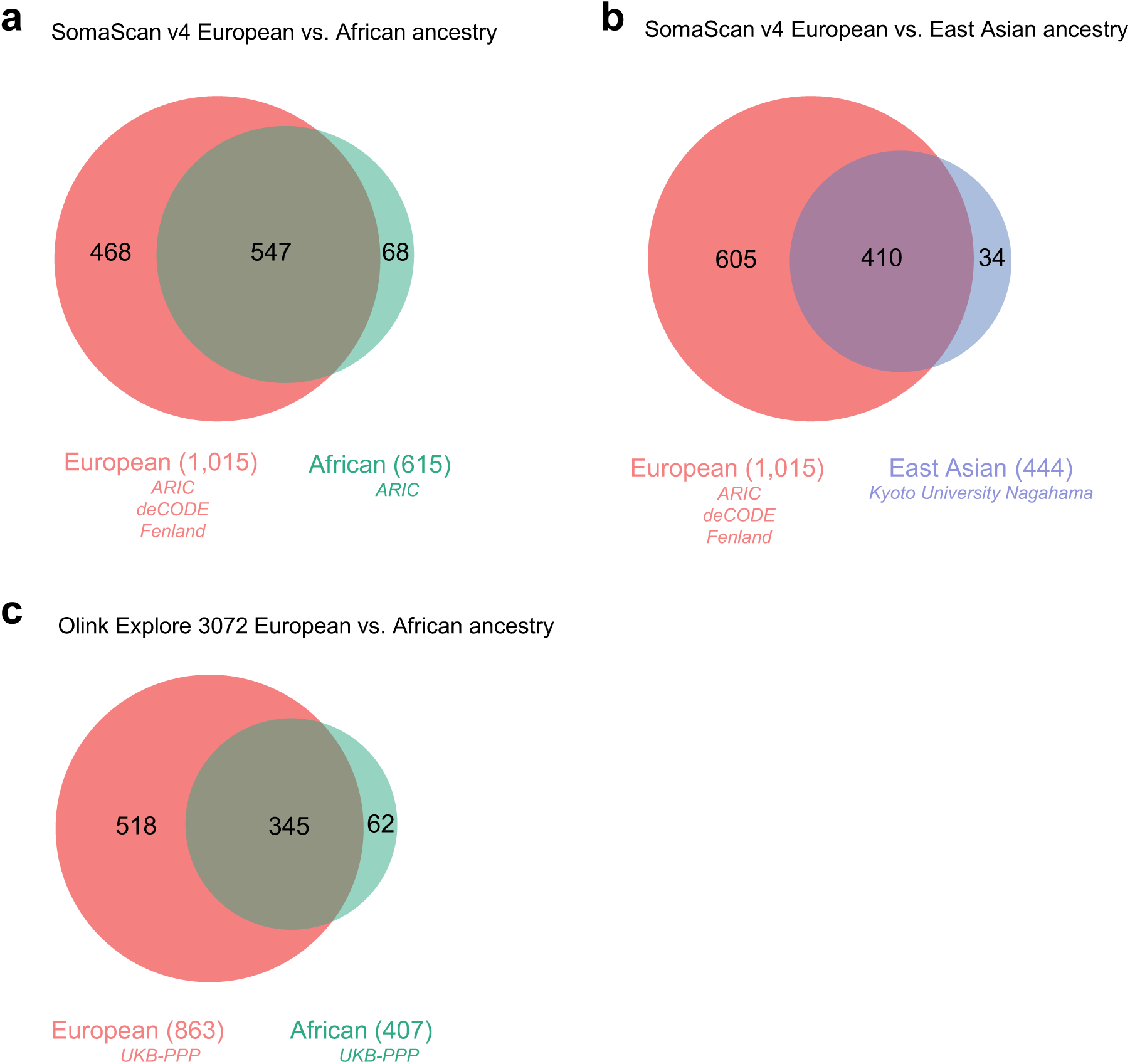
Cross-ancestry comparison stratified by proteomics platform of instrumentable proteins overlapping at least one drug database (druggable genome, DrugBank, or Open Targets Platform). (a) Comparison of SomaScan v4 platform instrumentable proteins overlapping at least one drug database between three European cohorts (ARIC, deCODE, and Fenland) and one African cohort (ARIC). (b) Comparison of SomaScan v4 platform instrumentable proteins overlapping at least one drug database between three European cohorts (ARIC, deCODE, and Fenland) and one East Asian cohort (Kyoto University Nagahama). (c) Comparison of Olink Explore 3072 platform instrumentable proteins overlapping at least one drug database between one European cohort (UKB-PPP) and one African cohort (UKB-PPP).

