## Supplementary Info and Supplementary Figures for "Multi-ancestry proteome-phenome-wide Mendelian randomization offers a comprehensive protein-disease atlas and potential therapeutic targets"

### Supplementary Information

This file contains **Supplementary Note 1-11** and **Supplementary Figures 1-9** followed by the **Supplementary Note-only References.**

The Table of Contents is as follows:

**Supplementary Note 1-11:**


**Supplementary Note 1A:** *Characterizing newly defined cis-pQTLs within European and African ancestries*

**Supplementary Note 1B:** *Further details on defining "strict variant-to-gene" (strict V2G) cis-pQTLs*

**Supplementary Note 1C:** *Further details on instrumentable proteins in each cohort*

**Supplementary Note 2:** *Assessing sample overlap in two-sample MR for UK Biobank GWAS outcomes and proteomics cohort*

**Supplementary Note 3:** *Cohort level description of putatively causal protein-phenotype pairs in European and African ancestry*

**Supplementary Note 4:** *Proteins involved in a multitude of traits and diseases*

**Supplementary Note 5A:** *Validating cardiovascular related putatively causal protein-phenotype pairs from prior studies in European ancestries*

**Supplementary Note 5B:** *Validating non-cardiovascular disease related putatively causal protein-phenotype pairs from prior studies in European ancestries*

**Supplementary Note 5C:** *Exploratory analyses using ARIC African proteomics cohort to assess causal effects on binary cardiovascular and autoimmune-related outcomes in the Million Veteran Program*

**Supplementary Note 5D:** *Additional note on uniquely instrumentable proteins in East Asian ancestry and putatively causal protein-phenotype pairs from these proteins*

**Supplementary Note 6:** *Discordant effects across ancestries for putatively causal protein-phenotypes pairs*

**Supplementary Note 7A:** *Druggability of instrumentable protein-coding genes at the cohort level*

**Supplementary Note 7B:** *Overlap of protein-phenotype pairs stratified by ancestry in the druggable genome and DrugBank*

**Supplementary Note 7C:** *Druggability of protein-phenotype pairs by integrating the druggable genome, DrugBank, and Open Targets Platform*

**Supplementary Note 8:** *Prioritization of targets for CAD and T2D*

**Supplementary Note 9:** *Detailed description of Kyoto University Nagahama East Asian cohort*

**Supplementary Note 10:** *Detailed description of PWCoCo and SharePro*

**Supplementary Note 11:** *STROBE-MR checklist*

**Supplementary Figure 1-9:**

**Supplementary Figure 1. Within-ancestry *cis*-pQTL effect size concordance.**

**Supplementary Figure 2. Protein-phenotype network plots for other phenotypes in European ancestry; six phenotype categories)**

**Supplementary Figure 3. Protein-phenotype pairs with discordant direction across ancestries.**

**Supplementary Figure 4. The overlap between instrumentable protein-coding genes and the druggable genome from Finan et al.**

**Supplementary Figure 5. UpSet plot showing the overlap between instrumentable proteins and the druggable genome across three ancestries.**

**Supplementary Figure 6. UpSet plot showing the overlap between instrumentable proteins and the druggable genome across 7 cohorts.**

**Supplementary Figure 7. European ancestry druggability heatmaps for 12 disease categories.**

**Supplementary Figure 8. East Asian ancestry druggability heatmaps for 11 disease categories.**

**Supplementary Figure 9. Prioritizing proteins for coronary artery disease and type 2 diabetes.**

**Supplementary Note-only References**

**Supplementary Note 1-11:**

**Supplementary Note 1A:** *Characterizing newly defined cis-pQTLs within European and African ancestries*

To verify that newly defined *cis*-pQTLs were comparable, we assessed the within-ancestry concordance of *cis*-pQTL effect sizes in European (**Supplementary Figure 1a**) and in African ancestries (**Supplementary Figure 1b**). We found high concordance of effects suggesting that within each ancestry, the newly defined *cis*-pQTLs are comparable.

Methods**:**

We assessed the within-ancestry concordance between European ancestry proteomics cohorts by aligning the effect allele of *cis*-pQTLs in each cohort to the minor allele in the UKB 50k reference panel. Similarly, when comparing the within-ancestry concordance between African ancestry proteomics cohorts, we first aligned effect alleles of *cis*-pQTLs in each cohort to the minor allele of the corresponding variant in the African ancestry HGDP+1kGP reference panel.

**Supplementary Note 1B:** *Further details on defining "strict variant-to-gene" (strict V2G) cis-pQTLs*

Upon defining *cis*-pQTLs in each cohort, we performed two additional steps to select genetic instruments which we term strict V2G *cis*-pQTLs.

First, to minimize potential horizontal pleiotropic effects, we removed *cis*-variants associated with two or more protein-coding genes (**Supplementary Note Table 1**), thus retaining *cis*-pQTLs associated with a single protein-coding gene, which we term "strict" *cis*-pQTLs.

Second, we leveraged multiple sources of evidence to assign variants to genes using Open Targets Genetics V2G score. As expected, upon performing strict V2G filtering within each cohort, we found that strict V2G *cis*-pQTLs had a larger proportion of proteins with a single associated *cis*-pQTL compared to all *cis*-pQTLs (**Supplementary Note Table 2**), and the maximum number of *cis*-pQTLs associated with a single protein was either the same or lower across all cohorts in all ancestries (**Supplementary Note Table 3**). The number of strict V2G *cis*-pQTLs for each protein is shown in **Supplementary Note Table 4**. The maximum number of strict V2G *cis*-pQTLs per protein was larger in all European ancestry cohorts compared to African ancestry cohorts and the Kyoto University Nagahama East Asian ancestry cohort (**Supplementary Note Table 3**). For example, the UKB-PPP European ancestry cohort had a range of 1 up to 21 strict V2G *cis*-pQTLs per protein while the number of strict V2G *cis*-pQTLs per protein in the Kyoto University Nagahama East Asian ancestry cohort ranged from 1 to 3.

**Supplementary Note 1C:** *Further details on instrumentable proteins in each cohort*

In European ancestries, we identified 1,485 instruments for 1,102 proteins for ARIC (*n* = 7,213; **Supplementary Table 2**), 2,083 instruments for 1,243 proteins for deCODE (*n* = 35,559; **Supplementary Table 3**), 1,637 instruments for 1,194 proteins for Fenland (*n* = 10,708; **Supplementary Table 4**), and 2,194 instruments for 1,292 proteins for UKB-PPP (*n* = 34,557; **Supplementary Table 5**). In African ancestries, we identified 1,080 instruments for 877 proteins for ARIC (*n* = 1,871; **Supplementary Table 6**), and 604 instruments for 554 proteins for UKB-

PPP ($n$ = 931; **Supplementary Table 7**). Finally, in the Kyoto University Nagahama East Asian ancestry cohort, 663 instruments were identified from 602 proteins ($n$ = 1,823; **Supplementary Table 8**).

We also assessed whether any instrumentable proteins were shared across cohorts within European and African ancestries. Here, we use proteins to refer to the protein-coding gene name in order to avoid double counting SomaScan v4 aptamers and to enable harmonization with Olink assays. Protein-coding genes were quantified based on Ensembl gene IDs. Within European ancestries, 434 proteins were shared across all fours cohorts while 375 were unique to the ARIC, deCODE, and Fenland cohorts measured with the SomaScan v4 platform, and 652 were unique to the UKB-PPP cohort which measured proteins using Olink Explore 3072 (**Extended Data Fig. 4a**), highlighting the value of using two proteomics platforms. In African ancestries, 259 proteins were jointly instrumentable by the ARIC and UKB-PPP cohorts while 591 were unique to ARIC and 294 to UKB-PPP (**Extended Data Fig. 4b**) which emphasizes the value of having two cohorts from two separate proteomics platforms. The Kyoto University Nagahama East Asian cohort ($n$ = 1,823) was able to instrument 581 proteins (**Extended Data Fig. 4c**).

**Supplementary Note 2:** *Assessing sample overlap in two-sample MR for UK Biobank GWAS outcomes and proteomics cohort*

To estimate the extent of bias in two-sample MR causal estimates for European exposures that use proteomics GWAS from the UKB-PPP and European GWAS outcomes that were generated from UK Biobank individuals, we calculated relative bias[1] based on the equation:

$$Relative\ bias = \phi \times \frac{1}{F}$$

The proportion of sample overlap, $\phi$, ranges between 0 (no sample overlap) and 1 (complete sample overlap), while the F-statistic of the exposure is denoted by $F$. Assessments of sample overlap for the European UKB-PPP and African UKB-PPP proteomics cohorts and corresponding UKB outcomes are shown in **Supplementary Note Table 5**.

Since 51 of 179 European GWAS outcomes were based on the UK Biobank, we estimated the extent of potential bias towards the null due to sample overlap in two-sample MR causal estimates for European exposures that used proteomics GWAS from the UKB-PPP ($n$ = 34,557). The relative bias[1] when assuming maximum overlap between proteomics GWAS and the 51 UKB outcome GWAS using the minimum F-statistic of 29.8 was estimated to be between 0.250% and 0.769% (**Supplementary Note Table 5**). However, we note that we utilized three additional proteomics cohorts from ARIC, deCODE, and Fenland which may consequently provide further levels of support for these protein-phenotype associations and mitigate this bias.

**Supplementary Note 3:** *Cohort level description of putatively causal protein-phenotype pairs in European and African ancestry*

Our analysis involves two different levels of resolution when describing the results. At the ancestry level, this pertains to European, African, and East Asian ancestries. At the cohort level this involves four cohorts (ARIC, deCODE, Fenland, and UKB-PPP) for European ancestries, two cohorts (ARIC and UKB-PPP) for African ancestries, and the Kyoto University Nagahama cohort for individuals of East Asian ancestry. In the main text, we focus findings at the ancestry level to avoid complicating the results and describe the cohort level details here:

In European ancestries, we identified a total of 6,771 putatively causal protein-phenotype associations (1,764 pairs in ARIC; 1,731 in deCODE, 1,788 in Fenland, and 1,488 in UKB-PPP) pertaining to 3,949 unique pairs (**Supplementary Table 14** and **Extended Data Fig. 5a**). In African ancestries, we identified a total of 72 associations involving 35 associations in ARIC and 37 associations in UKB-PPP, with 56 unique protein-phenotype associations across both cohorts involving 28 proteins and 11 phenotypes (**Supplementary Table 15** and **Extended Data Fig. 5b**). The direction of effect was consistent among all common associations between ARIC and UKB-PPP including well known proteins known to affect HDL cholesterol such as APOC1, CD36, DPEP2, and ITIH4 serving as positive controls.

**Supplementary Note 4:** *Proteins involved in a multitude of traits and diseases*

When quantifying the number of associated phenotypes that each protein was associated with, we found pleiotropic proteins associated with up to 49, 5, and 58 unique phenotypes in European, African, and East Asian ancestries, respectively (**Supplementary Note Table 6**). In European ancestries, 13 proteins were putatively causal for 20 or more outcomes including GCKR, GPN1, BTN3A2, SORT1, RSPO3, HP, RAB21, TIMD4, MST1, APOB, EFEMP1, PCSK9, and PLCG1. Since some phenotype categories had a greater number of outcomes than other categories, we quantified the number of associated phenotype categories each protein was implicated in as well (**Supplementary Note Table 7**). These targets present complex scenarios and may have effects in multiple tissues or organs.

For example, in European ancestries, GCKR (implicated in 10 phenotype categories) increases the risk of type 2 diabetes as previously reported[2] but our study shows that it also decreases the risk of inflammatory bowel disease (IBD) and cholelithiasis. MST1 (9 phenotype categories) is a macrophage-stimulating protein and hepatocyte growth factor-link protein highly expressed in the liver, suggesting its involvement in immune-related and liver diseases. MST1 has been implicated in IBD[3] and its pQTLs have also been found to associate with Crohn's disease, ulcerative colitis, IBD, and primary sclerosing cholangitis (PSC)[4], a rare liver condition associated with IBD and causing severe liver scarring. We found that increased genetically predicted MST1 levels were protective against IBD, including both Crohn's disease and ulcerative colitis, and PSC. However, despite these protective effects, our results suggest that higher MST1 protein levels may potentially lead to harmful cardiovascular events, such as increased systolic blood pressure and diastolic blood pressure and a higher risk of coronary artery disease.

In African ancestries, APOA5, APOE, and HP had effects on the most phenotypes with both being associated with 5 outcomes (**Supplementary Note Table 6**). All proteins had effects on phenotypes within a single category aside from ABO and ITIH4 which were causal for outcomes from three and two phenotype categories, respectively (**Supplementary Note Table 7**).

In East Asian ancestries, ALDH2 was the most pleiotropic protein and putatively causal for 58 phenotypes (from 12 phenotype categories) including multiple diseases such as stroke (any ischemic stroke), epilepsy, colorectal cancer, esophageal cancer, hepatic cancer, lung cancer among many others. Notably, ALDH2 was not instrumented in European nor African and was uniquely instrumented in East Asian ancestry. The next most pleiotropic protein in East Asian ancestries was ABO associated with 14 phenotypes (4 phenotype categories) as expected due to known pleiotropy at this locus. MLN, a small peptide hormone secreted by cells in the small intestine which regulates gastrointestinal contractions and motility was causal for 10 phenotypes (6 phenotype categories) including cardiovascular outcomes (angina pectoris and stable angina pectoris), gastrointestinal (chronic hepatitis B), autoimmune (rheumatoid arthritis), cancer (gastric cancer), and various biomarkers implicating its involvement in multiple biological processes and influence in a multitude of health conditions.

To summarize, of our putatively causal findings, we identified many proteins influencing traits or diseases in the same categories revealing the common mechanistic interplay between specific outcomes. On the contrary, many proteins also demonstrated effects on various traits or diseases that may not be directly linked to each other, underscoring the complexity of protein functions and the intricate network of biological pathways involved in health and disease. For instance, a higher genetically predicted level of MST1 decreased the risk of IBD and its subtypes including both Crohn's disease and ulcerative colitis. This protective effect is likely due to MST1's role in modulating immune responses and inflammation, which are central to the pathogenesis of these conditions. Meanwhile, increased MST1 was associated with adverse cardiovascular outcomes including higher risk of coronary artery disease. In the context of IBD, MST1 may enhance mucosal healing and modulate inflammatory pathways, contributing to its protective effects. Conversely, the impact of MST1 on the cardiovascular system may involve mechanisms related to vascular inflammation and endothelial function, leading to increased blood pressure and atherosclerosis, highlighting the importance of understanding context-specific regulation of these potential targets.

**Supplementary Note 5A:** *Validating cardiovascular related putatively causal protein-phenotype pairs from prior studies in European ancestries*

We validated many previously known findings for cardiovascular phenotypes in European ancestries (**Fig. 4a** and **Supplementary Figure 2a**). For instance, COL6A3 (SomaScan aptamer: 11196-31, Olink assay: OID20292) was positively associated with coronary artery disease (CAD) in all four European ancestry cohorts. This effect is concordant with our previous extensive work on COL6A3[5]. In addition, we replicated previously reported findings of MMP12 on stroke by Sun et al.[6] that was also further validated by Zheng et al.[7] who extended to stroke subtypes. Similar to Zheng's study, European ancestry MMP12 pQTLs from deCODE and UKB-PPP were associated with lower risk of any ischemic stroke (deCODE: OR = 0.92, 95% CI: 0.89–0.95, $P = 1.9 \times 10^{-7}$, $PP_{max} = 0.98$; UKB-PPP: OR = 0.92, 95% CI: 0.90–0.95, $P = 1.9 \times 10^{-7}$, $PP_{max} = 1$) and large artery stroke (deCODE: OR = 0.77, 95% CI: 0.70–0.86, $P = 6.0 \times 10^{-7}$, $PP_{max} = 1$; UKB-PPP: OR = 0.79, 95% CI: 0.72–0.87, $P = 6.0 \times 10^{-7}$, $PP_{max} = 0.99$).

Additionally, ITIH4 was shown in a prior study using mouse lines and colocalization to act as a novel vascular smooth muscle cell-expressed gene implicated in atherosclerotic plaques[8]. However, genetic evidence of causality was not determined. Here, we found a positive association between ITIH4 with CAD (deCODE: OR = 1.43, 95% CI: 1.27–1.60, $P = 1.5 \times 10^{-9}$, $PP_{max} = 1$; Fenland: OR = 1.18, 95% CI: 1.12–1.25, $P = 1.5 \times 10^{-9}$, $PP_{max} = 1$), pulse pressure (deCODE: $\beta = 1.46$, 95% CI: 1.00–1.93, $P = 5.5 \times 10^{-10}$, $PP_{max} = 1$; Fenland: $\beta = 0.69$, 95% CI: 0.47–0.91, $P = 5.5 \times 10^{-10}$, $PP_{max} = 0.99$), and systolic blood pressure (deCODE: $\beta = 1.66$, 95% CI: 0.98–2.34, $P = 1.8 \times 10^{-6}$, $PP_{max} = 0.97$; Fenland: $\beta = 0.78$, 95% CI: 0.46–1.11, $P = 1.8 \times 10^{-6}$, $PP_{max} = 0.94$), consistent with and confirming the findings of earlier research using animal studies.

Interestingly, we also identified a few proteins that were protective against cardiovascular events. For example, SWAP70 was protective against coronary artery disease (ARIC: OR = 0.95, 95% CI: 0.93–0.96, $P = 5.7 \times 10^{-11}$, $PP_{max} = 1$; deCODE: OR = 0.92, 95% CI: 0.89–0.96, $P = 1.5 \times 10^{-9}$, $PP_{max} = 0.94$), small vessel stroke (deCODE: OR = 0.78, 95% CI: 0.68–0.89, $P = 2.6 \times 10^{-4}$, $PP_{max} = 0.84$), and any ischemic stroke (ARIC: OR = 0.94, 95% CI: 0.92–0.97, $P = 4.5 \times 10^{-6}$, $PP_{max} = 0.98$; deCODE: OR = 0.90, 95% CI: 0.86–0.95, $P = 3.0 \times 10^{-5}$, $PP_{max} = 0.92$).

PTN was protective against peripheral artery disease in two cohorts (Fenland: OR = 0.79, 95% CI: 0.70–0.89, $P = 7.5 \times 10^{-5}$, $PP_{max} = 0.97$, and UKB-PPP: OR = 0.49, 95% CI: 0.37–0.65, P = $8.5 \times 10^{-7}$, $PP_{max} = 0.89$), mood swings (deCODE: OR = 0.98, 95% CI: 0.97–0.99, $P = 1.4 \times 10^{-5}$, $PP_{max} = 0.81$; Fenland: OR = 0.97, 95% CI: 0.96–0.99, $P = 1.4 \times 10^{-5}$, $PP_{max} = 0.84$), and anthropometric outcomes such as waist-to-hip ratio (deCODE: $\beta = -0.04$, 95% CI: -0.06, -0.03, $P = 2.3 \times 10^{-6}$, $PP_{max} = 0.97$; Fenland: $\beta = -0.04$, 95% CI: -0.07, -0.03, $P = 4.0 \times 10^{-6}$, $PP_{max} = 0.95$).

Further, DKKL1 was previously reported as putatively causal for multiple sclerosis[9] which we also found (ARIC: OR = 0.46, 95% CI: 0.37–0.58, $P = 3.5 \times 10^{-11}$, $PP_{max} = 1$; Fenland: OR = 0.28, 95% CI: 0.19–0.41, $P = 4.3 \times 10^{-11}$, $PP_{max} = 1$), but here we also identified it as protective against risk of large artery stroke (ARIC: OR = 0.59, 95% CI: 0.44–0.79, $P = 4.2 \times 10^{-4}$, $PP_{max} = 0.87$; Fenland: OR = 0.42, 95% CI: 0.26–0.67, $P = 3.0 \times 10^{-4}$, $PP_{max} = 0.89$) as well as metabolic/endocrine disorders such as hypothyroidism/myxoedema (ARIC: OR = 0.99, 95% CI: 0.98–0.99, $P = 1.2 \times 10^{-5}$, $PP_{max} = 0.95$; Fenland: OR = 0.98, 95% CI: 0.97–0.99, $P = 2.6 \times 10^{-5}$, $PP_{max} = 0.84$).

**Supplementary Note 5B:** *Validating non-cardiovascular disease related putatively causal protein-phenotype pairs from prior studies in European ancestries*

We also validated findings in other non-cardiovascular disease categories in European ancestries. Network plots for these associations are shown for autoimmune (**Supplementary Figure 2b**), neurological (**Supplementary Figure 2c**), psychiatric (**Supplementary Figure 2d**), metabolic/endocrine (**Supplementary Figure 2e**), and gastrointestinal phenotypes (**Supplementary Figure 2f**).

For example, a one standard deviation increase in genetically predicted NPNT levels was associated with a decreased risk of asthma which is consistent with findings in our previous work[10].

Notably, increased levels of genetically predicted RAB21 was protective against Parkinson's disease and associated with increased cognitive performance and educational attainment consistent with findings of the involvement of RAB21 in neuronal development[11] (**Supplementary Figure 2c**). Further, RAB21 has been implicated in obesity in concord with our findings showing protective effects of RAB21 on visceral (VAT), abdominal subcutaneous (ASAT), and gluteofemoral (GFAT) adipose tissue volumes (**Supplementary Figure 2e**).

INHBB has previously been implicated in serum urate levels[12] and in our analysis was found to be positively associated with urate and negatively associated with estimated glomerular filtration rate in all four European ancestry cohorts. In addition, we found a positive effect of genetically predicted INHBB levels on risk of type 2 diabetes in the UKB-PPP cohort (OR: 1.09, 95% CI: 1.04–1.13 per standard deviation (s.d.) increase in the protein level, $P = 9.8 \times 10^{-5}$, $PP_{max} = 1$) which had not previously been reported.

Lastly, STAT3 was positively associated with inflammatory bowel disease but negatively associated with multiple sclerosis which is concordant with recent findings showing opposite effects across these two diseases[13] and supported by an earlier randomized, placebo-controlled multicenter study showing divergent outcomes of anti-TNF therapies, which are effective for inflammatory bowel disease but worsen multiple sclerosis[14].

**Supplementary Note 5C:** *Exploratory analyses using ARIC African proteomics cohort to assess causal effects on binary cardiovascular and autoimmune-related outcomes in the Million Veteran Program*

We identified 7 putatively causal associations for binary cardiovascular outcomes which are shown in **Supplementary Note Table 8**. The protein PCYOX1 was implicated in all diseases.

**Supplementary Note 5D:** *Additional note on uniquely instrumentable proteins in East Asian ancestry and putatively causal protein-phenotype pairs from these proteins*

Of the 325 unique protein-phenotype pairs identified in the Kyoto University Nagahama East Asian ancestry cohort, we found that 67 (20.6%) protein-phenotype associations were from 8 proteins (ALDH2, ANXA7, APOA1, DDOST, GSS, PLA2G7, PRSS2, UGT1A1) specific to East Asian and not instrumentable by European nor African ancestries. For instance, in European ancestry, ALDH2 had no genome-wide significant pQTLs in ARIC, and upon LD clumping, only had *trans*-pQTLs in UKB-PPP, while in both Fenland and deCODE, the strict *cis*-pQTL did not have the highest V2G score and was filtered out given the risk of horizontal pleiotropy. Similarly, in African ancestries, ALDH2 had no genome-wide significant pQTLs upon LD clumping.

An example of one association from uniquely instrumentable proteins in East Asian ancestry is APOA1 and cholesterol levels. APOA1 was positively associated with both HDL cholesterol and total cholesterol levels concordant with its function as the main protein in high density lipoproteins mediating efflux of cholesterol.  Additionally, causal effects of UGT1A1 on total bilirubin has been previously identified in African ancestries[15] and our study supports these findings in East Asian ancestries ($\beta = -0.44$, 95% CI: -0.46, -0.43, $P = 1.00 \times 10^{-300}$, $PP_{max} = 1$).

**Supplementary Note 6:** *Discordant effects across ancestries for putatively causal protein-phenotypes pairs*

For putatively causal associations with inconsistent MR effect estimates across ancestries, we identified 12 protein-phenotype pairs involving 9 proteins and 6 phenotypes, composed of lipid and anthropometric traits (**Supplementary Figure 3**). One pair showed discordant MR estimates across all three ancestries, four were discordant between European and African ancestries, and seven between European and East Asian ancestries. For instance, ABO was negatively associated with total cholesterol levels in Europeans but positively associated in African and East Asian ancestries. This discrepancy may require further study due to the high-impact PAV instrumental variable used for ABO in African ancestries (**Supplementary Table 15**). Similar discordance was seen with ABO and LDL cholesterol: negative in Europeans and positive in East Asian ancestries, likely due to ABO's pleiotropic nature. CD36 also showed discordant associations: positively associated with HDL cholesterol in Europeans (deCODE, Fenland, UKB-PPP) but negatively in Africans (ARIC, UKB-PPP), and negatively associated with triglycerides in Europeans but positively in Africans. The *cis*-pQTL proxying CD36 in African ancestries was a high-impact PAV. While increased CD36 typically decreases HDL and increases triglycerides, a previous study showed an inverse relationship between monocyte CD36 and HDL in African ancestries[16], aligning with our findings. Other discordant associations included DEF6 with height (negative in Europeans, positive in Africans) and CA4 (positive in Europeans, negative in Africans). Between European and East Asian ancestries, discordance was found for GHR and AOC1 with height, APOB with LDL and total cholesterol, and ACP1 with body mass index. Notably, GHR and height associations were inconsistent within European cohorts (deCODE and UKB-PPP), likely due to differences in proteomics platforms.

We also identified discordant effects for phenotypically related outcomes that were not exact matches and highlight an example using APOB. APOB plays a predominant role in the etiology of coronary artery disease as shown in recent studies[17–19]. Likewise, our results showed that APOB was positively associated with coronary artery disease (CAD) in European ancestries with evidence in deCODE ($\beta$ = 0.65, 95% CI: 0.51–0.80, $P = 3.4 \times 10^{-18}$, $PP_{max}$ = 1), and Fenland ($\beta$ = 0.43, 95% CI: 0.33–0.53, $P = 2.9 \times 10^{-18}$, $PP_{max}$ = 1). However, APOB in East Asian ancestries was found to be negatively associated with myocardial infarction ($\beta$ = -0.08, 95% CI: -0.11,-0.05, $P = 1.4 \times 10^{-8}$, $PP_{max}$ = 1), LDL cholesterol ($\beta$ = -0.06, 95% CI: -0.07, -0.05, $P = 1.3 \times 10^{-30}$, $PP_{max}$ = 1), HMG CoA reductase inhibitors ($\beta$ = -0.14, 95% CI: -0.16, -0.12, $P = 3.1 \times 10^{-44}$, $PP_{max}$ = 1), and vasodilators used in cardiac diseases ($\beta$ = -0.06, 95% CI: -0.08, -0.03, $P = 5.0 \times 10^{-6}$, $PP_{max}$ = 0.98). When querying the instruments used for APOB in European ancestries in Open Targets Genetics we found that the single *cis*-pQTL proxying APOB in deCODE, rs563290, and Fenland, rs541041, were intergenic variants (Ensembl VEP impact: Modifier). In contrast, the most severe consequence of the pQTL for APOB in East Asian ancestries, rs13306194, was missense (Ensembl VEP impact: Moderate) and could potentially be altering epitope binding rather than being a true biological signal. Nonetheless, this pQTL may still hold biological significance due to being the top hit from variant-to-gene mapping, although further investigation may be required.

In summary, while inconsistent effects were found across ancestries for a few protein-phenotype associations we highlight that in three associations, ABO with total cholesterol levels, CD36 with HDL cholesterol levels, and CD36 with triglycerides, the protein level in African ancestries was a PAV of high impact so we advise caution in the interpretation of these results. Further, ABO is known to play a multi-faceted role in diseases and pQTLs at this locus have been associated with many proteins[6]. However, since we used variant-to-gene mapping which leverages biological evidence to select instruments, these PAVs of high impact may potentially still be functionally relevant. Notably, we found that decreased CD36 in African ancestries was associated with increased HDL cholesterol which was consistent with findings from a previous study[16]. Thus, further exploration may be required with larger sample sizes to elucidate whether these findings are biologically plausible.

**Supplementary Note 7A:** *Druggability of instrumentable protein-coding genes at the cohort level*

We compared instrumentable protein-coding genes against the druggable genome composed of 4,479 genes from Finan et al.[20] which classifies genes into Tier 1, 2, or 3 according to druggability. Tier 1 refers to efficacy targets of approved small molecules, biotherapeutic drugs, and clinical-phase drug candidates; Tier 2 includes proteins closely associated with drug targets or linked to drug-like compounds; Tier 3 encompasses secreted or extracellular proteins, those distantly related to approved drug targets, and proteins from important druggable gene families not covered in Tier 1 or Tier 2. All ancestries had proportionally comparable number of instrumentable protein-coding genes in Tier 1 and 2 (**Supplementary Figure 4**).

We found that 71, 29, and 210 proteins were shared in Tier 1, Tier 2, and Tier 3 across three ancestries, respectively (**Supplementary Figure 5**) while 26, 10, and 78 proteins were shared in Tier 1, Tier 2, and Tier 3 among all seven cohorts across three ancestries, respectively (**Supplementary Figure 6**).

**Supplementary Note 7B:** *Overlap of protein-phenotype pairs stratified by ancestry in the druggable genome and DrugBank*

*7B.1 Druggable genome*

We found that 57.7%, 78.5% and 74.2% of putatively causal protein-phenotype associations in European, African, and East Asian ancestries, respectively, overlapped with the druggable genome (**Supplementary Note Table 9**).

*7B.2. DrugBank*

Across ancestries, a similar proportion of proteins from the putatively causal protein-phenotype associations—33.6% in European, 35.7% in African and 35.5% in East Asian ancestry—had approved or investigational drugs available based on DrugBank[21] (**Supplementary Note Tables 10–12).**

**Supplementary Note 7C:** *Druggability of protein-phenotype pairs by integrating the druggable genome, DrugBank, and Open Targets Platform*

We provide druggability visualization for proteins implicated in various diseases (stratified by disease category) which may be potentially explored as opportunities for drug development for European (**Supplementary Figure 7**) and East Asian ancestries (**Supplementary Figure 8**). For instance, increased genetically predicted ANGPTL4 (Tier 3 target) leads to increased risk of CAD (**Supplementary Figure 7 - Cardiovascular**).

**Supplementary Note 8:** *Prioritization of targets for CAD and T2D*

We applied filtering steps to prioritize proteins involved in CAD (**Supplementary Figure 9a**) and T2D (**Supplementary Figure 9b**). See **Supplementary Note 8 Methods** below. In European ancestries, we found directional concordance between MR estimates and hazard ratios from Cox regression for incident CAD for 18 proteins, ANGPTL4, C1R, C1S, COL6A3, COMT, DDT, DUSP13, FES, FN1, IL6R, ITIH4, MST1, PCSK9, PDE5A, PLG, SCARF2, TGFB1, and TIMP2. Two of these proteins, ITIH4 and ANGPTL4 were putatively causal for more than one phenotype in African ancestries, albeit for different outcomes, while three proteins, IL6R, PCSK9, and PLG were putatively causal for more than one phenotype in East Asian ancestries (**Supplementary Figure 9a, Supplementary Note Table 13**).

We also found directional concordance between MR estimates and hazard ratios from Cox regression for incident T2D for 8 proteins, ACE, ANGPTL4, INHBB, LRIG1, MINDY1, PAM, PAPPA, and TFRC in European ancestries. ANGPTL4 was the only putatively causal protein from this list present in African ancestries while in East Asian ancestries, three of these proteins, INHBB, LRIG1, and TFRC were putatively causal (**Supplementary Figure 9b, Supplementary Note Table 14**).

We found that, in European ancestries, each standard deviation increase in ANGPTL4 levels was associated with increased odds of incident CAD (OR = 1.17, SE = 0.016, $P = 1.89 \times 10^{-22}$) and increased odds of incident T2D (OR = 1.21, SE = 0.023, $P = 4.02 \times 10^{-17}$). These results are consistent with our findings from MR, where increased circulating ANGPTL4 levels were associated with increased risk of CAD and T2D. Notably, MR estimates in African ancestries for ANGPTL4 also supported this concordance in European ancestries with ANGPTL4 being negatively associated with high density lipoprotein cholesterol levels and positively associated with triglycerides supporting ANGPTL4 as a potential therapeutic target for intervention.

In addition, each standard deviation increase in INHBB levels was associated with a 1.19-fold increased hazard of T2D (SE = 0.022, $P = 3.63 \times 10^{-14}$), aligning with MR findings of increased T2D risk. MR evidence in East Asian ancestries showed that higher genetically predicted INHBB levels increase the risk of blood urea nitrogen and G-glutamyl transpeptidase, both linked to T2D risk. However, in East Asian ancestries, reducing INHBB levels might lower HDL cholesterol and raise LDL cholesterol, potentially increasing cardiovascular risk. Thus, therapeutic strategies targeting INHBB must be carefully evaluated for adverse lipid profile effects. Comprehensive research is needed to ensure benefits outweigh risks and to develop strategies that selectively modulate INHBB without harming cardiovascular health.

We found corroborative evidence of causality between ANGPTL4 and coronary artery disease and type 2 diabetes which was validated through observational association analyses on incident coronary artery disease and incident type 2 diabetes risk in European ancestries. Additionally, through MR we also identified similar causal roles for ANGPTL4 in African ancestries for related biomarkers such as HDL cholesterol and triglycerides which were in directions congruent with our CAD and T2D findings. ANGPTL4 is an inhibitor of lipoprotein lipase (LPL) which leads to increased triglyceride levels and prior studies have shown that the ANGPTL4 coding variant E40K has been associated with lower plasma triglyceride levels[22] and lower risk of both CAD and T2D[23]. While no current therapies exist for ANGPTL4, a closely related protein ANGPTL3, which also inhibits LPL does and is targeted by zodasiran, an RNA interference therapy. Zodasiran targets ANGPTL3 expression in the liver and demonstrated efficacy in reducing triglyceride levels in patients with mixed hyperlipidemia[24]. Mechanistically, ANGPTL4 and ANGPTL3 act differently. Whereas ANGPTL3 inhibitory activity on LPL is hindered through binding to heparin, ANGPTL4 is unaffected by heparin binding suggesting that ANGPTL4 may employ distinct regulatory mechanisms in modulating LPL activity, potentially involving alternative molecular interactions or structural characteristics that confer resistance to heparin inhibition. Further research into these mechanisms could unveil novel therapeutic strategies for managing cardiovascular diseases.

#### Supplementary Note 8 Methods

##### *8.1. Prioritizing targets for CAD and T2D*

For European protein-phenotype associations, we first subsetted to those with consistent MR effect across all four cohorts (ARIC, deCODE, Fenland, and UKB-PPPP) then integrated Cox regression effects and determined directional concordance between MR effect estimates and observational association estimates. Those with discordance between the two estimates were removed and the remaining proteins were retained as potential candidates. We performed this procedure for both incident CAD and incident T2D. A detailed flow diagram is shown in **Supplementary Figure 9**.

We highlight that when comparing the direction of MR effect estimates with observational association estimates, we removed proteins with inconsistent direction and those which did not have an observational association estimate. Since Cox regression was performed on the UKB-PPP Olink Explore 3072 proteins, MR effect estimates from SomaScan v4 proteins would not have a corresponding observational association estimate meaning proteins exclusive to SomaScan v4 were not considered. Therefore, only proteins from Olink or common proteins between SomaScan and Olink were analyzed in this analysis.

##### *8.2. Observational associations between circulating protein abundances and incident CAD and T2D in the UK Biobank*

We assessed whether MR effect estimates were in alignment with observational associations as this can provide an additional source of evidence supporting the purported protein-phenotype association. To perform observational association analysis on incident CAD and T2D, we used individual level data from the UK Biobank to determine whether circulating protein abundances were able to predict future risk of these diseases based on 10 years of follow-up.

CAD was defined as in our previous study[5]. Briefly, we used three criteria: (i) a record of ICD-10 codes I20-I25 (ischemic heart disease), (ii) an operation record of percutaneous transluminal coronary angioplasty (PTCA) or coronary artery bypass grafting (CABG), and (iii) a death record associated with ICD-10 codes I20-I25. The time to event was determined by subtracting the event registration date from the enrollment date (data field: 53), focusing on events occurring within 10 years of enrollment. We excluded individuals with pre-existing CAD who met these criteria prior to enrollment, as well as those without a recorded event date. Controls were defined as individuals without a CAD record based on doctor diagnosis (data field: 6150), self-reported heart attack (data field: 20002), or an ICD-10 record of I20-I25.

T2D was defined using ICD-10 code E-11 while controls were defined as individuals without any type of diabetes based on self-reports.

We used Cox proportional hazards models (function: coxph()) adjusting for age, sex, BMI, recruitment center, time to Olink processing, batch, and the first 10 principal components to associate 2,922 Olink 3072 Explore platform proteins from 4,750 cases with incident CAD and 32,565 controls and 3,066 cases with incident T2D and 37,453 controls. Protein levels were inverse rank normal transformed prior to analysis.

**Supplementary Note 9:** *Detailed description of Kyoto University Nagahama East Asian cohort*

*9.1. Study Cohort*

Whole-genome sequencing (WGS) and proteome analysis were conducted using samples from the Nagahama Prospective Genome Cohort for Comprehensive Human Bioscience (Nagahama Study). A subset of 2,000 individuals (1,392 women, mean age 56.7 years; 608 men, mean age 62.0 years) was selected from 8,559 participants in the first follow-up health check (2012-2016). Ethical approval was obtained from the Kyoto University Graduate School of Medicine and the Nagahama Municipal Review Board (No. 278). All participants provided written informed consent.

*9.2. Plasma Samples and Protein Quantification*

Plasma was isolated from EDTA-treated blood by centrifugation and stored at −80 °C. Protein levels were measured using SomaScan assay v4, targeting 4,740 unique proteins with 5,284 SOMAmers. After quality control, 1,997 plasma samples and 4,392 SOMAmers (4,196 proteins) were retained. Data were normalized and used for protein quantitative trait locus (pQTL) analysis. Some proteins had multiple SOMAmers targeting different forms, distinguished by annotations in the SomaId and Target columns.

*9.3. Whole-Genome Sequencing*

WGS was performed on 1,573 samples using Illumina platforms and 385 samples using a DNBSEQ-G400 instrument, following standard protocols (GATK and DRAGEN). After quality control, including kinship analysis and variant concordance checks, 1,823 samples and 4,642,253 variants were retained for protein association analysis.

*9.4. pQTL Analysis*

Genetic associations with 4,392 SOMAmers were analyzed using PLINK (v.2.00a3LM) with age, sex, variant-calling pipeline, and the first five genetic principal components as covariates. The genome-wide significance threshold was set at 1.08 × 10−8 after Bonferroni correction.

*9.5. Linkage Disequilibrium (LD) Clumping*

LD clumping was performed using PLINK, defining clumping regions as 500 kb around index variants with an LD threshold of $R^2 \geq 0.8$. Each region was assigned a unique ID.

*9.6. Conditional Analysis*

Independent signals at the loci were identified through stepwise selection in GCTA-COJO with the following parameters: --maf 0.01 --cojo-slct, --cojo-collinear 0.9 –cojo-p 5e-8

**Supplementary Note 10:** *Detailed description of PWCoCo and SharePro*

We performed colocalization analyses using PWCoCo and SharePro as complementary methods to safeguard against potential putative causal associations confounded by LD and ensure higher confidence in our findings. Since differences in LD structures between populations under study can introduce bias in MR analyses, the presence of a shared causal variant between the exposure and outcome can help mitigate this issue and increase the robustness of the MR findings.

*10.1. Pairwise conditional and colocalization analysis (PWCoCo)*

Pairwise conditional and colocalization analysis (PWCoCo) (https://github.com/jwr-git/pwcoco) integrates methods from conditional analyses (GCTA-COJO)[25] and colocalization analyses (coloc)[26] which relaxes the simplified single causal variant assumption of coloc thereby allowing the assessment of whether multiple causal variants exist and colocalize within a region. Through conditional analyses, independent signals from both traits (here protein GWAS and outcome GWAS) can be identified and colocalization can be conducted on each pair of conditionally independent signals for the two GWAS while upholding the strict single variant assumption of coloc. PWCoCo has been shown to outperform existing methods in scenarios where the single variant assumption is violated[27]. It also enables the identification of previously missed disease-causing variants through its ability to perform independent colocalization of secondary signals and offers key improvements through its computational efficiency and ease-of-use. We performed PWCoCo using default settings and set the maximum number of causal variants in the region, *k*, to 5. PWCoCo, similar to coloc, reports five colocalization probabilities: H0 – no association with either trait; H1 – association with trait 1, not with trait 2; H2 – association with trait 2, not with trait 1; H3 – association with trait 1 and trait 2, two independent SNPs; H4 – association with trait 1 and trait 2, one shared SNP (i.e., the probability that both traits are associated through the sharing of a single causal variant). We considered the maximum H4 posterior probability (PPH4) across all tested pairs of conditionally independent signals and report evidence of colocalization if the maximum PPH4 ≥ 0.8. We note that this is different from $PP_{max}$ used in the main text which we use to denote the maximum PP between PWCoCo and SharePro colocalization methods.

*10.2. Shared sparse Projection for colocalization analysis (SharePro)*

Shared sparse Projection for colocalization analysis (SharePro) (https://github.com/zhwm/SharePro_coloc) is a novel colocalization method which extends upon the coloc framework. SharePro uses an efficient variational inference algorithm that leverages LD modelling and integration with colocalization assessment through grouping of correlated variants into effect groups to accurately estimate posterior colocalization probabilities which together overcome the aforementioned limitations. Further, SharePro has increased power for identifying biologically plausible signals in simulation analyses, outperforming coloc and PWCoCo while maintaining low computational cost and a low false positive rate[28]. We used SharePro (v.5.0.0) default settings which sets the maximum number of causal variants in the region, K, to 10.

We emphasize that for SharePro, when K is larger than the true number of causal variants, it consistently provides adequate results in comparison to setting K to the true number of causal variants. Namely, K = 10 should always yield results similar to, or better than K = 5 which was set in PWCoCo due to the COJO step requiring a substantial increase in computational time with a larger defined K, which is not the case in SharePro.

For the required LD input files to SharePro, we used the UKB 50k reference panel for European, the HGDP + 1kGP reference panel for African, and the 1kGP East Asian reference panel for East

Asian ancestries similar to what we utilized at the LD clumping stage previously described. SharePro reports the “share” column (colocalization probabilities) for all effect groups, and we reported evidence of colocalization for the protein GWAS and outcome GWAS pair if the maximum colocalization probability of any effect group was > 0.8. We also note that this is different from $PP_{max}$ used in the main text which we use to denote the maximum PP between PWCoCo and SharePro colocalization methods.

**Supplementary Note 11:**
**STROBE-MR checklist of recommended items to address in reports of Mendelian randomization studies**[1 2]
Note: Page number will be added at the proof-reading stage.

| Item No. | Section | | Checklist item | Page No. | Relevant text from manuscript |
|---|---|---|---|---|---|
| 1 | **TITLE and ABSTRACT** | | Indicate Mendelian randomization (MR) as the study's design in the title and/or the abstract if that is a main purpose of the study | 1, 2 | Specified in the title abstract |
| | **INTRODUCTION** | | | | |
| 2 | **Background** | | Explain the scientific background and rationale for the reported study. What is the exposure? Is a potential causal relationship between exposure and outcome plausible? Justify why MR is a helpful method to address the study question | 3 | Explained in paragraph 1 of the introduction section. |
| 3 | **Objectives** | | State specific objectives clearly, including pre-specified causal hypotheses (if any). State that MR is a method that, under specific assumptions, intends to estimate causal effects | 3 | Explained in paragraph 1 of the introduction section. |
| | **METHODS** | | | | |
| 4 | **Study design and data sources** | | Present key elements of the study design early in the article. Consider including a table listing sources of data for all phases of the study. For each data source contributing to the analysis, describe the following: | | Explained in the Methods section and sources of data are presented in supplementary table 1 (ST10-12). |
| | | a) | Setting: Describe the study design and the underlying population, if possible. Describe the setting, locations, and relevant dates, including periods of recruitment, exposure, follow-up, and data collection, when available. | | The study design and the underlying population are described in the Methods section. The remainder are described in the main text. |
| | | b) | Participants: Give the eligibility criteria, and the sources and methods of selection of participants. Report the sample size, and whether any power or sample size calculations were carried out prior to the main analysis | | (b)–(e) were described in the Methods section and Supplementary Note |
| | | c) | Describe measurement, quality control and selection of genetic variants | | |
| | | d) | For each exposure, outcome, and other relevant variables, describe methods of assessment and diagnostic criteria for diseases | | |
| | | e) | Provide details of ethics committee approval and participant informed consent, if relevant | | |
| 5 | **Assumptions** | | Explicitly state the three core IV assumptions for the main analysis (relevance, independence and exclusion restriction) as well assumptions for any additional or sensitivity analysis | | Explicitly stated in the introduction and in the Methods. |

| | | | | |
|---|---|---|---|---|
| 6 | **Statistical methods: main analysis** | | Describe statistical methods and statistics used | |
| | | a) | Describe how quantitative variables were handled in the analyses (i.e., scale, units, model) | (a)–(e) were described in the Methods as well as the Results section. |
| | | b) | Describe how genetic variants were handled in the analyses and, if applicable, how their weights were selected | |
| | | c) | Describe the MR estimator (e.g. two-stage least squares, Wald ratio) and related statistics. Detail the included covariates and, in case of two-sample MR, whether the same covariate set was used for adjustment in the two samples | |
| | | d) | Explain how missing data were addressed | |
| | | e) | If applicable, indicate how multiple testing was addressed | |
| 7 | **Assessment of assumptions** | | Describe any methods or prior knowledge used to assess the assumptions or justify their validity | 7–9 were described in the Methods section as well as the Results section. |
| 8 | **Sensitivity analyses and additional analyses** | | Describe any sensitivity analyses or additional analyses performed (e.g. comparison of effect estimates from different approaches, independent replication, bias analytic techniques, validation of instruments, simulations) | |
| 9 | **Software and pre-registration** | | | |
| | | a) | Name statistical software and package(s), including version and settings used | |
| | | b) | State whether the study protocol and details were pre-registered (as well as when and where) | |
| | **RESULTS** | | | |
| 10 | **Descriptive data** | | | |
| | | a) | Report the numbers of individuals at each stage of included studies and reasons for exclusion. Consider use of a flow diagram | Described in the Methods and Supplementary Tables. |
| | | b) | Report summary statistics for phenotypic exposure(s), outcome(s), and other relevant variables (e.g. means, SDs, proportions) | Described in the Methods and Supplementary Tables. |
| | | c) | If the data sources include meta-analyses of previous studies, provide the assessments of heterogeneity across these studies | Discussed in the original papers. We also evaluated the heterogeneity and horizontal pleiotropy in our analyses. |

| | | | | |
|---|---|---|---|---|
| | | d) | For two-sample MR:<br>i. Provide justification of the similarity of the genetic variant-exposure associations between the exposure and outcome samples<br>ii. Provide information on the number of individuals who overlap between the exposure and outcome studies | Described in the Methods, Results, and Supplementary Note. |
| 11 | **Main results** | | | |
| | | a) | Report the associations between genetic variant and exposure, and between genetic variant and outcome, preferably on an interpretable scale | (a)–(c) were described in the Results. |
| | | b) | Report MR estimates of the relationship between exposure and outcome, and the measures of uncertainty from the MR analysis, on an interpretable scale, such as odds ratio or relative risk per SD difference | |
| | | c) | If relevant, consider translating estimates of relative risk into absolute risk for a meaningful time period | |
| | | d) | Consider plots to visualize results (e.g. forest plot, scatterplot of associations between genetic variants and outcome versus between genetic variants and exposure) | Described in the Methods, Results, Main Figures and Supplementary Figures using forest plots. |
| 12 | **Assessment of assumptions** | | | |
| | | a) | Report the assessment of the validity of the assumptions | Described in the Methods and Results. |
| | | b) | Report any additional statistics (e.g., assessments of heterogeneity across genetic variants, such as $I^2$, Q statistic or E-value) | Described in the Methods and Results. |
| 13 | **Sensitivity analyses and additional analyses** | | | |
| | | a) | Report any sensitivity analyses to assess the robustness of the main results to violations of the assumptions | (a)-(d) were described in the Methods and Results. |
| | | b) | Report results from other sensitivity analyses or additional analyses | |
| | | c) | Report any assessment of direction of causal relationship (e.g., bidirectional MR) | |
| | | d) | When relevant, report and compare with estimates from non-MR analyses | |
| | | e) | Consider additional plots to visualize results (e.g., leave-one-out analyses) | We did not perform leave-one-out analyses but assessed the robustness of the analyses using two colocalization methods (PWCoCo and SharePro) |

| | | | | |
|---|---|---|---|---|
| | | | | which are described in the Methods, Results, and Supplementary Note. |
| | **DISCUSSION** | | | |
| 14 | **Key results** | | Summarize key results with reference to study objectives | Described in the Discussion. |
| 15 | **Limitations** | | Discuss limitations of the study, taking into account the validity of the IV assumptions, other sources of potential bias, and imprecision. Discuss both direction and magnitude of any potential bias and any efforts to address them | Described in the Discussion. |
| 16 | **Interpretation** | | | |
| | | a) | Meaning: Give a cautious overall interpretation of results in the context of their limitations and in comparison with other studies | (a)–(c) were described in the Results and Discussion. |
| | | b) | Mechanism: Discuss underlying biological mechanisms that could drive a potential causal relationship between the investigated exposure and the outcome, and whether the gene-environment equivalence assumption is reasonable. Use causal language carefully, clarifying that IV estimates may provide causal effects only under certain assumptions | |
| | | c) | Clinical relevance: Discuss whether the results have clinical or public policy relevance, and to what extent they inform effect sizes of possible interventions | |
| 17 | **Generalizability** | | Discuss the generalizability of the study results (a) to other populations, (b) across other exposure periods/timings, and (c) across other levels of exposure | Described in the limitations section in the Discussion. |
| | **OTHER INFORMATION** | | | |
| 18 | **Funding** | | Describe sources of funding and the role of funders in the present study and, if applicable, sources of funding for the databases and original study or studies on which the present study is based | Described in the Acknowledgments. |
| 19 | **Data and data sharing** | | Provide the data used to perform all analyses or report where and how the data can be accessed, and reference these sources in the article. Provide the statistical code needed to reproduce the results in the article, or report whether the code is publicly accessible and if so, where | Described in the Data Availability and Code availability. |
| 20 | **Conflicts of Interest** | | All authors should declare all potential conflicts of interest | Described in the Competing Interests |

This checklist is copyrighted by the Equator Network under the Creative Commons Attribution 3.0 Unported (CC BY 3.0) license.

1. Skrivankova VW, Richmond RC, Woolf BAR, Yarmolinsky J, Davies NM, Swanson SA, et al. Strengthening the Reporting of Observational Studies in Epidemiology using Mendelian Randomization (STROBE-MR) Statement. JAMA. 2021;under review.

2. Skrivankova VW, Richmond RC, Woolf BAR, Davies NM, Swanson SA, VanderWeele TJ, et al. Strengthening the Reporting of Observational Studies in Epidemiology using Mendelian Randomisation (STROBE-MR): Explanation and Elaboration. BMJ. 2021;375:n2233.

**Supplementary Figure 1. Within-ancestry *cis*-pQTL effect size concordance.**

*Cis*-pQTL effect size concordance for (a) European and (b) African ancestry cohorts. In European and African ancestry proteomics cohorts, the effect allele of *cis*-pQTLs in each cohort was aligned to the minor allele of the corresponding variant in their respective reference panels—UKB 50k for European and HGDP+1kGP for African ancestry—to harmonize alleles across each ancestral cohort for plotting. Red line is the diagonal while blue line is the best-fit line with standard errors shown by blue shading. Horizontal and vertical gray dashed lines show y = 0 and x = 0, respectively.

(a) European ancestry cohorts pairwise comparison

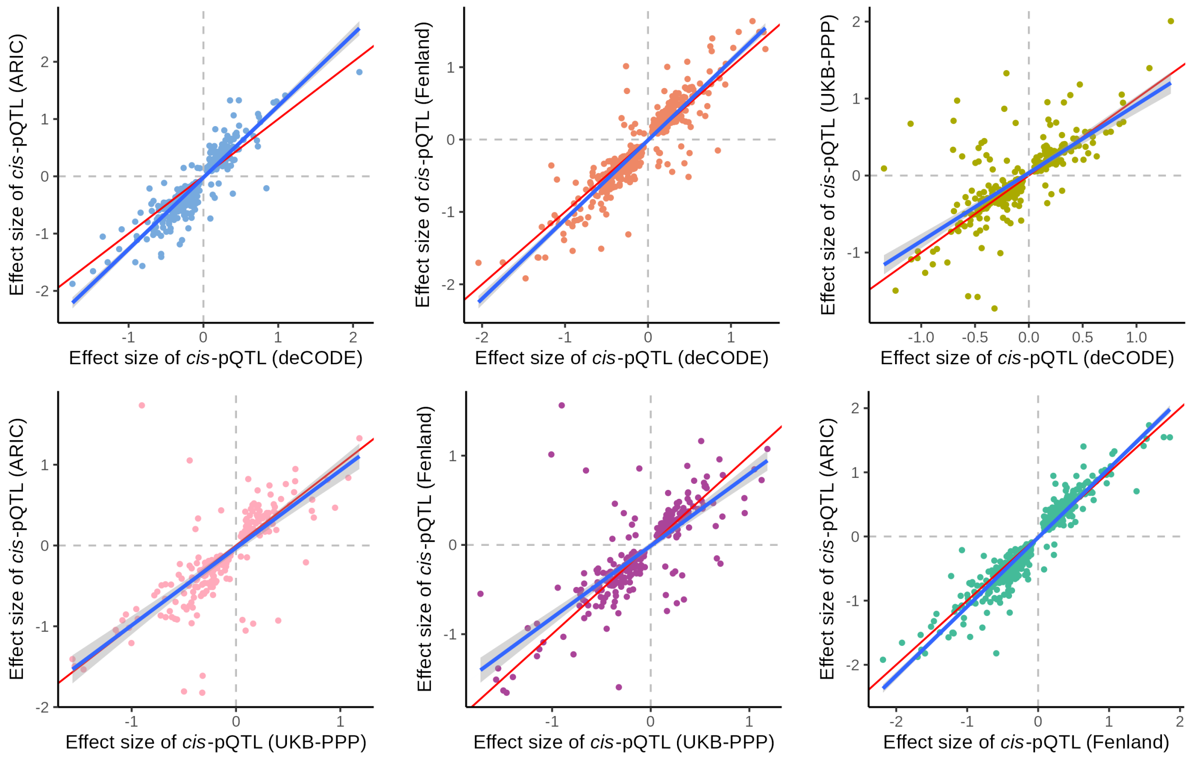


(b) African ancestry cohorts pairwise comparison

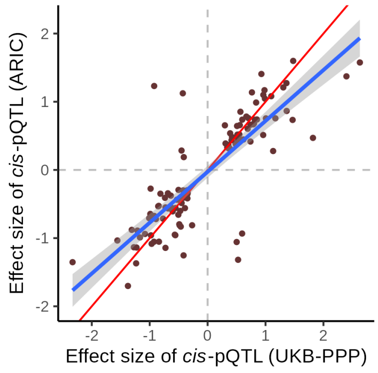

**Supplementary Figure 2. Protein-phenotype network plots for other phenotypes in European ancestry; six phenotype categories)**
Significant estimates between proteins (orange circles) and traits (green rectangles). Arrow thickness indicates how often a protein measurement has a causal effect on the outcome trait. For simplicity, we only depict protein-phenotype pairs in which all European cohorts showed concordant direction of effect estimates. Red arrows indicate a positive causal estimate of the protein on the outcome while blue arrows indicate a negative causal estimate of the protein on the outcome.

a. Cardiovascular (binary traits only)
b. Autoimmune
c. Neurological
d. Psychiatric
e. Metabolic/endocrine
f. Gastrointestinal

We note the following:
- Eczema appears in both Autoimmune and Skin network plots
- Type 1 diabetes appears in both Autoimmune and Metabolic/endocrine network plots
- Inflammatory bowel disease, Ulcerative colitis, Crohn’s disease, and Celiac disease appear in both Autoimmune and Gastrointestinal network plots

#### a. Cardiovascular (binary traits only)

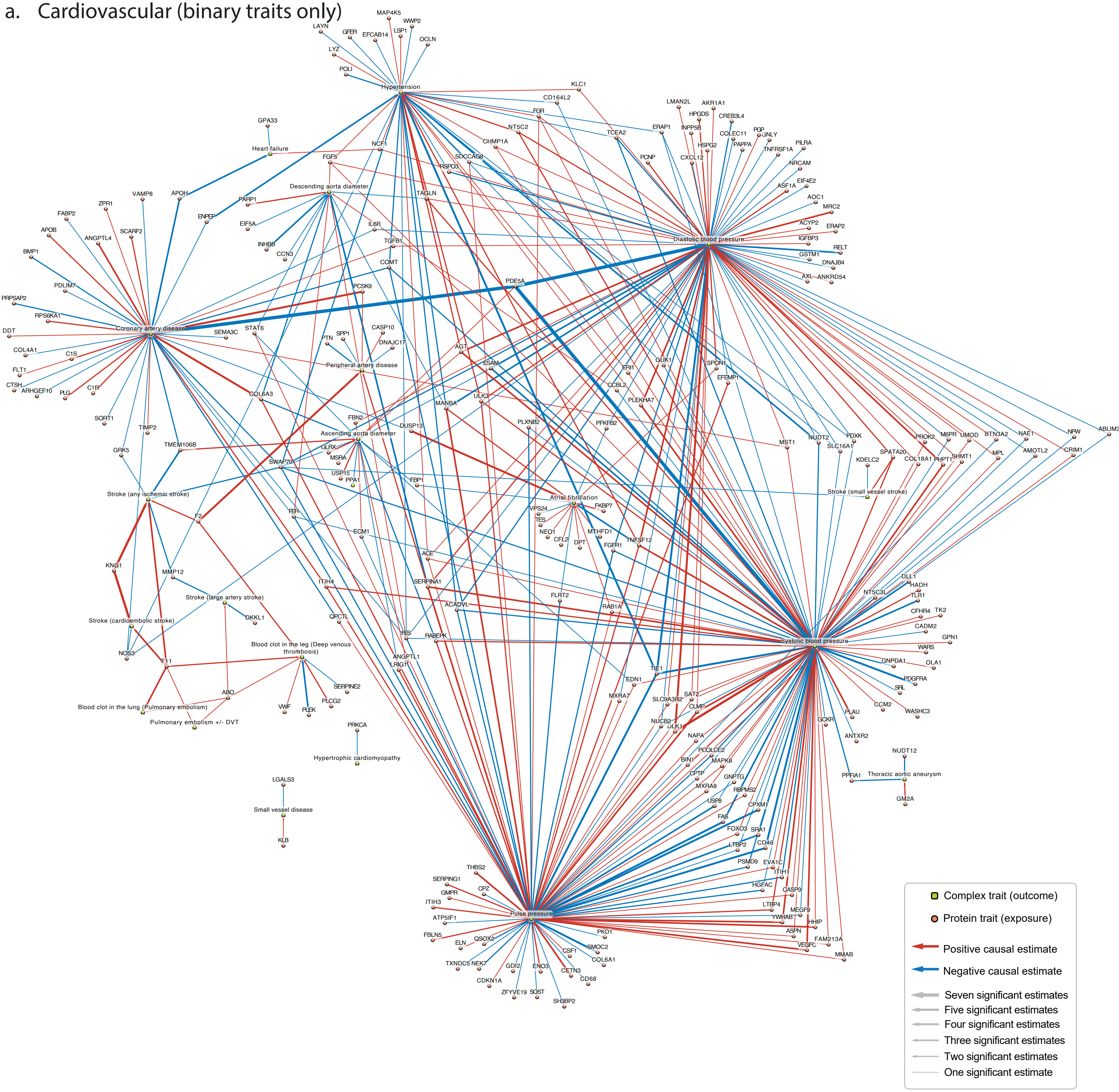

b. Autoimmune

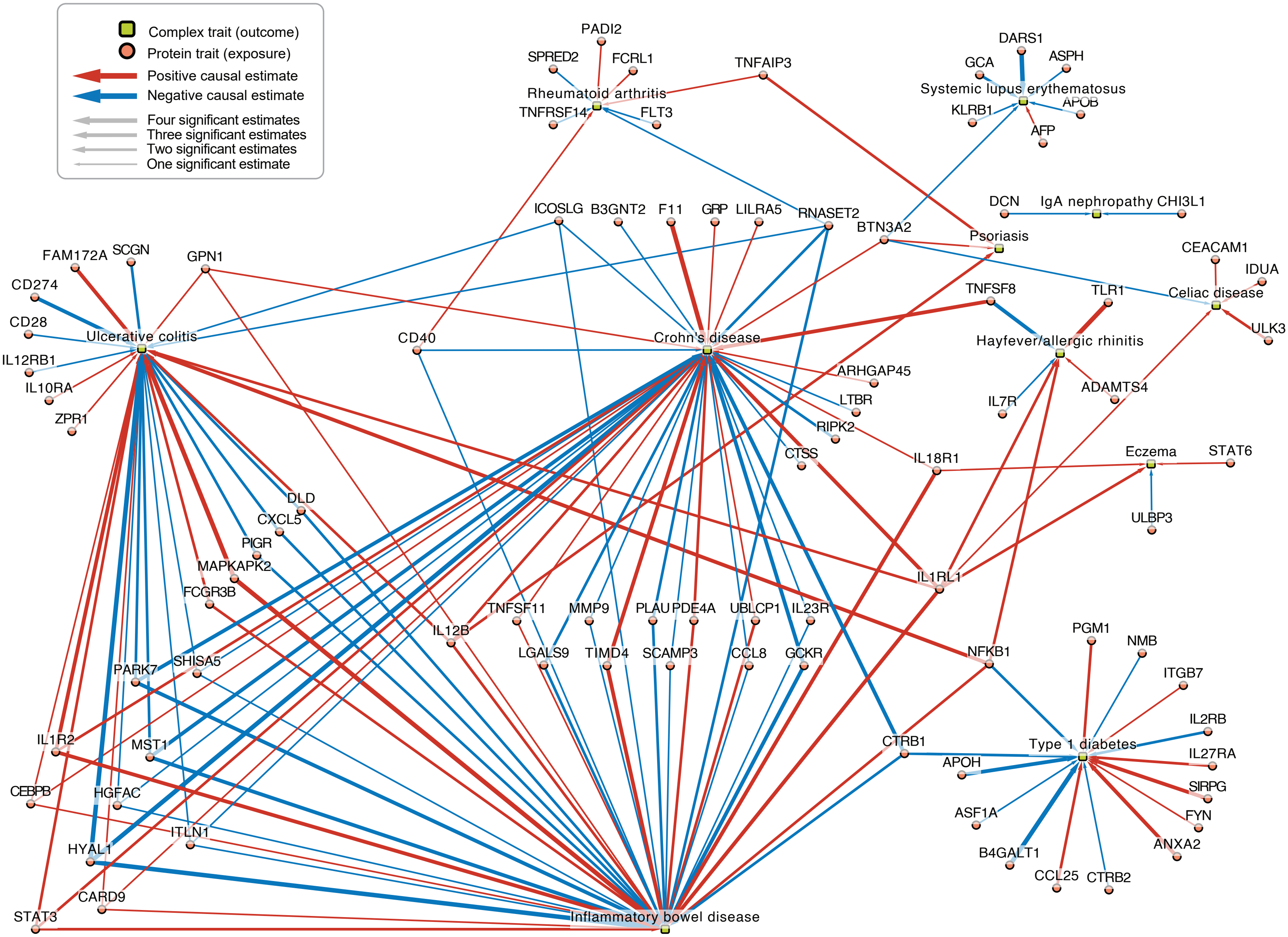

c. Neurological

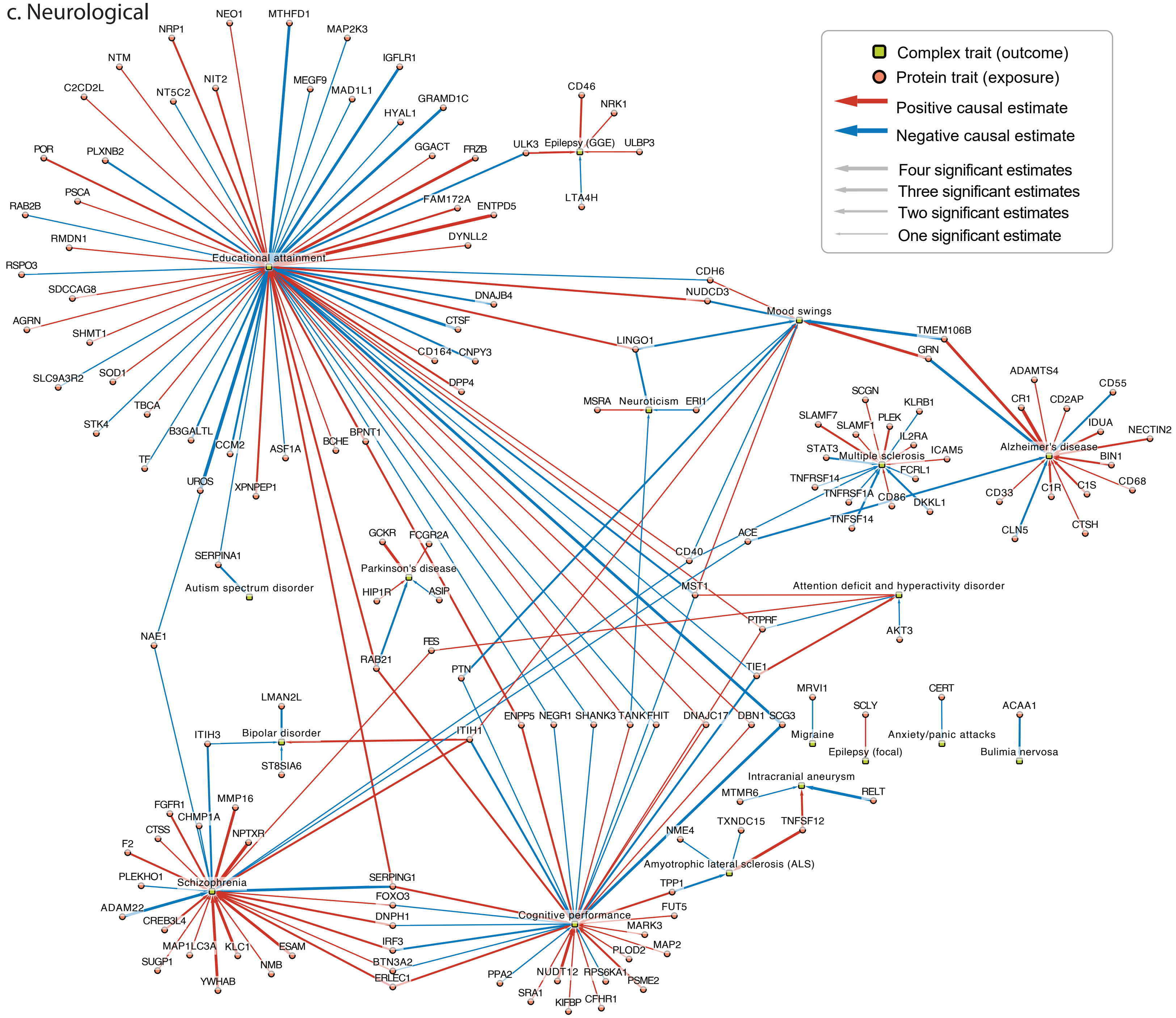

d. Psychiatric

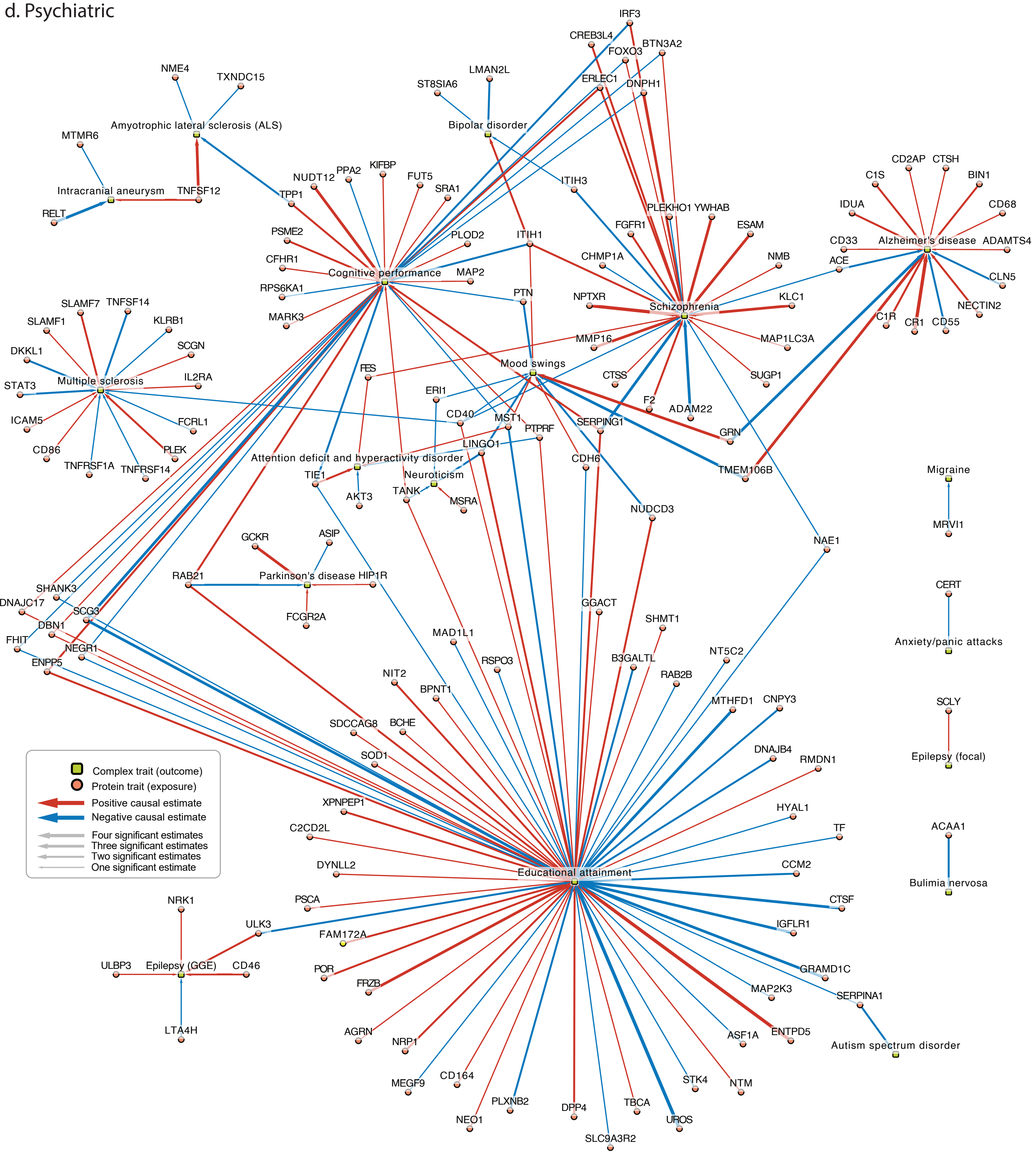

e. Metabolic/endocrine

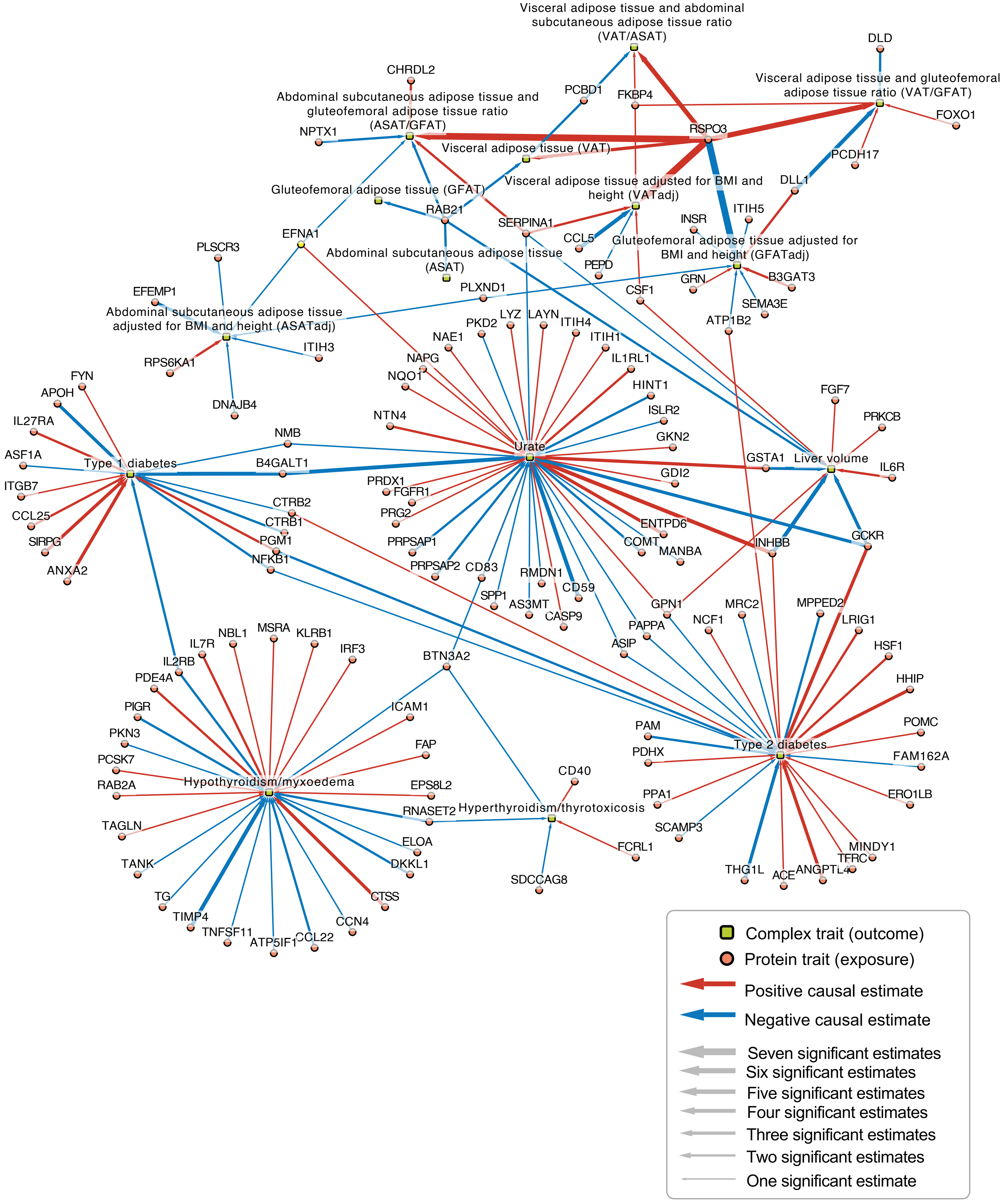

f. Gastrointestinal

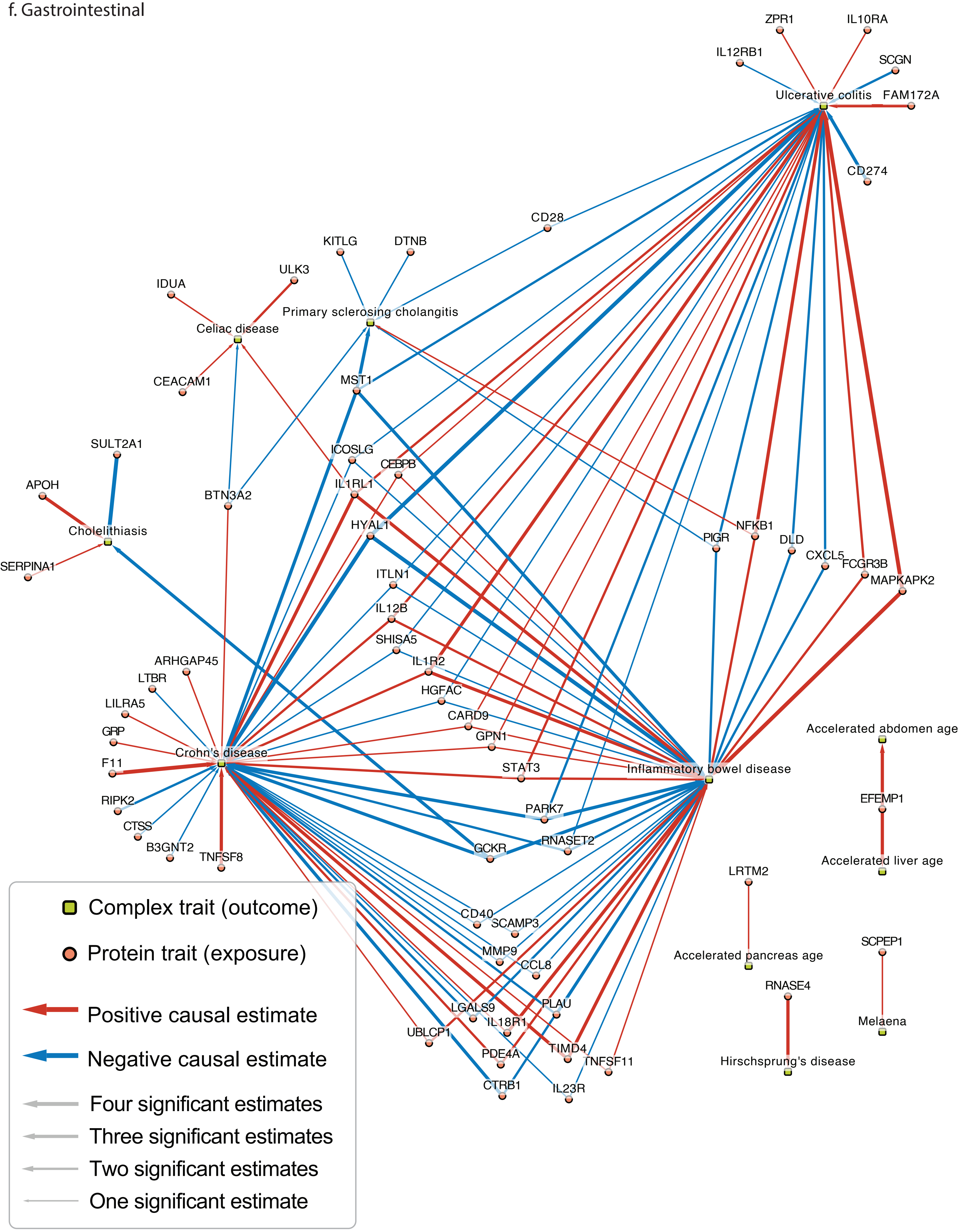

**Supplementary Figure 3. Protein-phenotype pairs with discordant direction across ancestries.** Protein-phenotype associations that have an estimated causal effect (FDR < 0.05), passed MR sensitivity analyses, and colocalized (PP.H4 > 0.8) in either PWCoCo or SharePro but had inconsistent direction of MR effect estimates across ancestries. Sample sizes for each outcome can be found in Supplementary Tables 13, 14, and 15 for European, African, and East Asian ancestries, respectively.

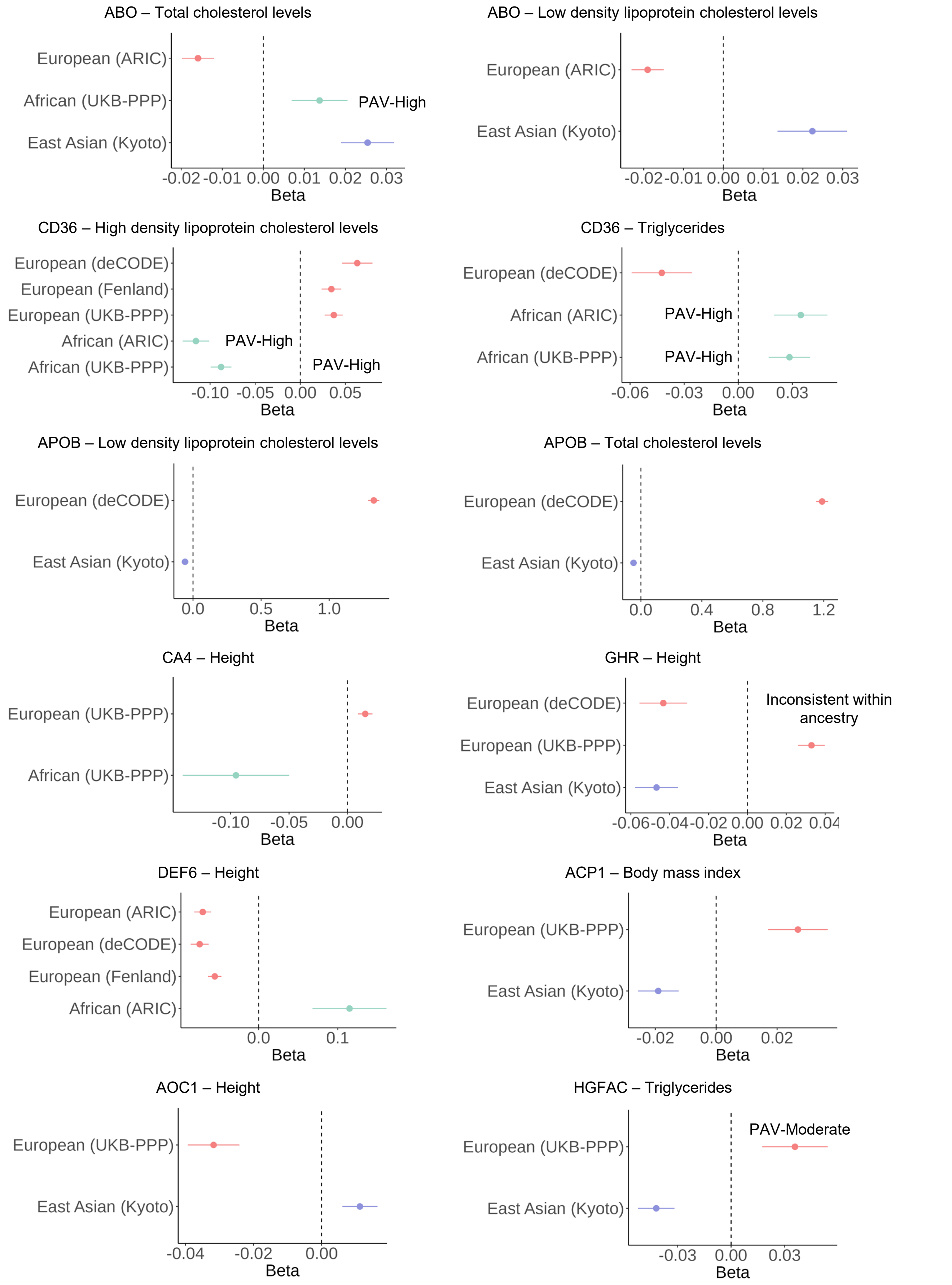

**Supplementary Figure 4. The overlap between instrumentable protein-coding genes and the druggable genome from Finan et al.**

The number of proteins classified into each tier when normalized by the number of instrumentable protein coding genes.

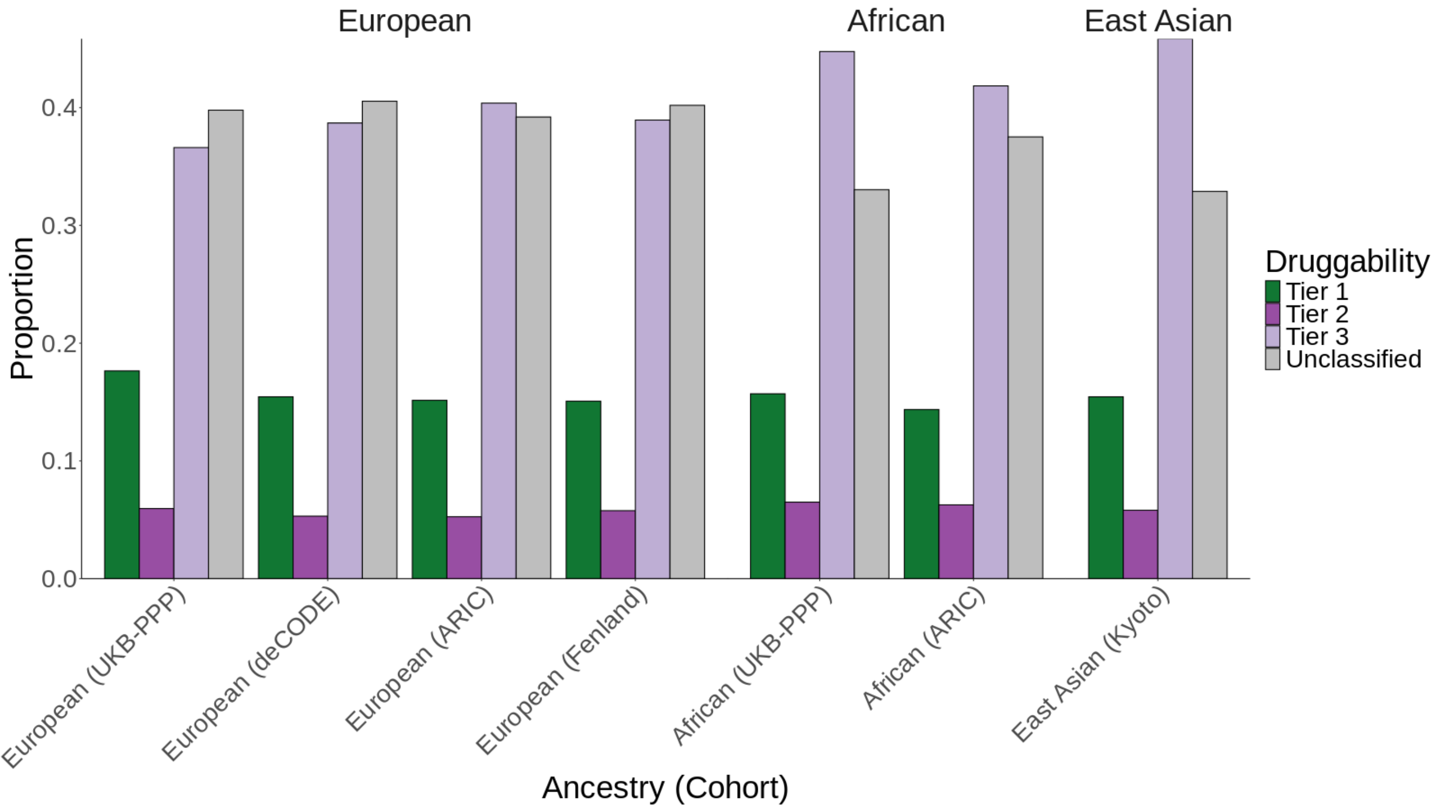

**Supplementary Figure 5. UpSet plot showing the overlap between instrumentable proteins and the druggable genome across three ancestries.**
(a) Tier 1, (b) Tier 2, (c) Tier 3, and (d) Unclassified groupings from Finan et al. The height of bars in the graphs display the number of instrumentable proteins common to a given cohort configuration. The x-axis shows the different cohort configurations. Horizontal bar graphs along with dots indicate a specific ancestry cohort and their frequency with ancestry denoted by shading (Green shading: East Asian ancestry cohort; Blue shading: African ancestry cohorts; Light red shading: European ancestry cohorts). The vertical red bar and red dotted line denote instrumentable proteins common to all cohorts across all three ancestries.

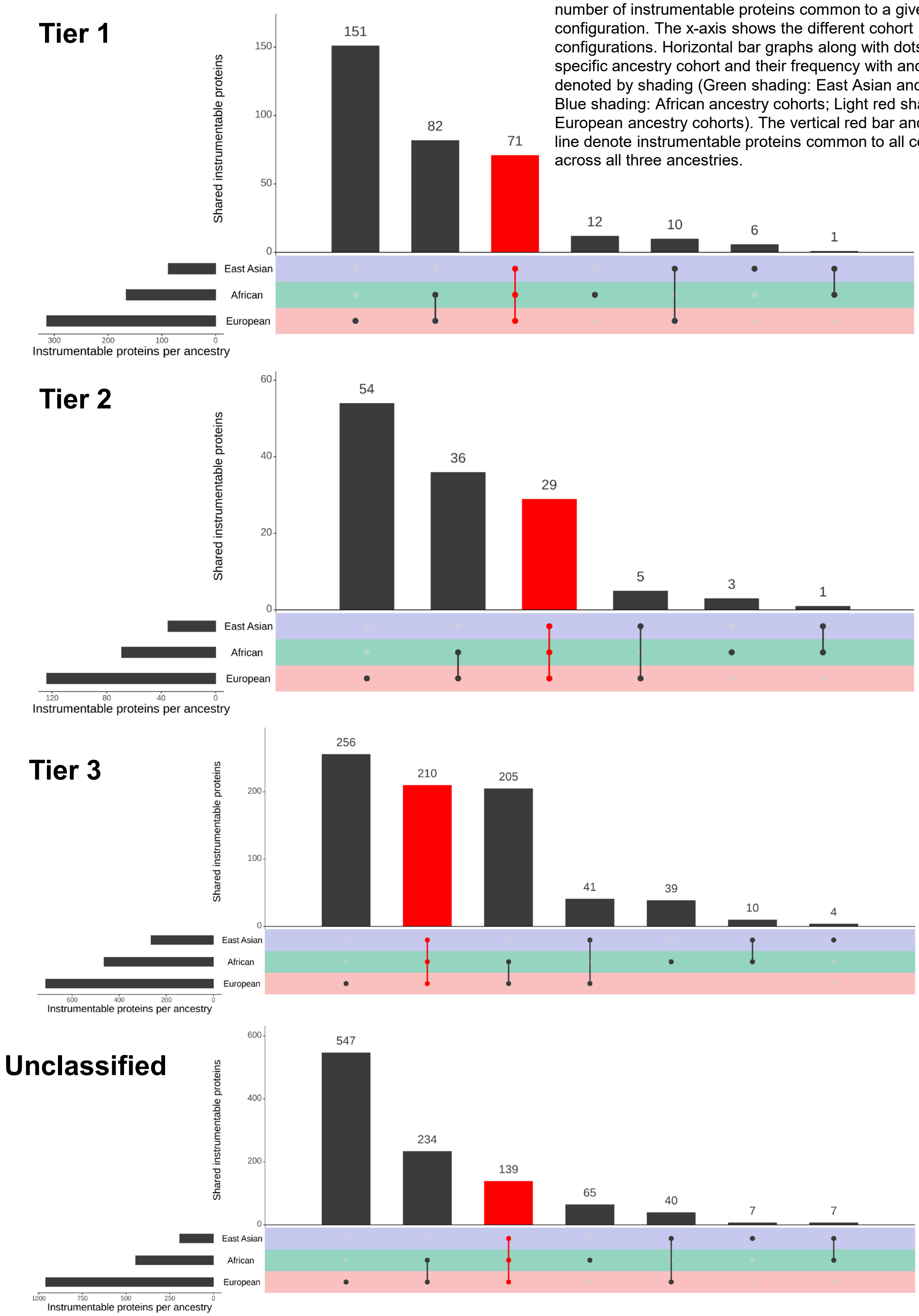

**Supplementary Figure 6. UpSet plot showing the overlap between instrumentable proteins and the druggable genome across 7 cohorts.**
(a) Tier 1, (b) Tier 2, (c) Tier 3, and (d) Unclassified groupings from Finan et al. The height of bars in the graphs display the number of instrumentable proteins common to a given cohort configuration. The x-axis shows the different cohort configurations. Horizontal bar graphs along with dots indicate a specific ancestry cohort and their frequency with ancestry denoted by shading (Green shading: East Asian ancestry cohort; Blue shading: African ancestry cohorts; Light red shading: European ancestry cohorts). The vertical red bar and red dotted line denote instrumentable proteins common to all cohorts across all seven cohorts.

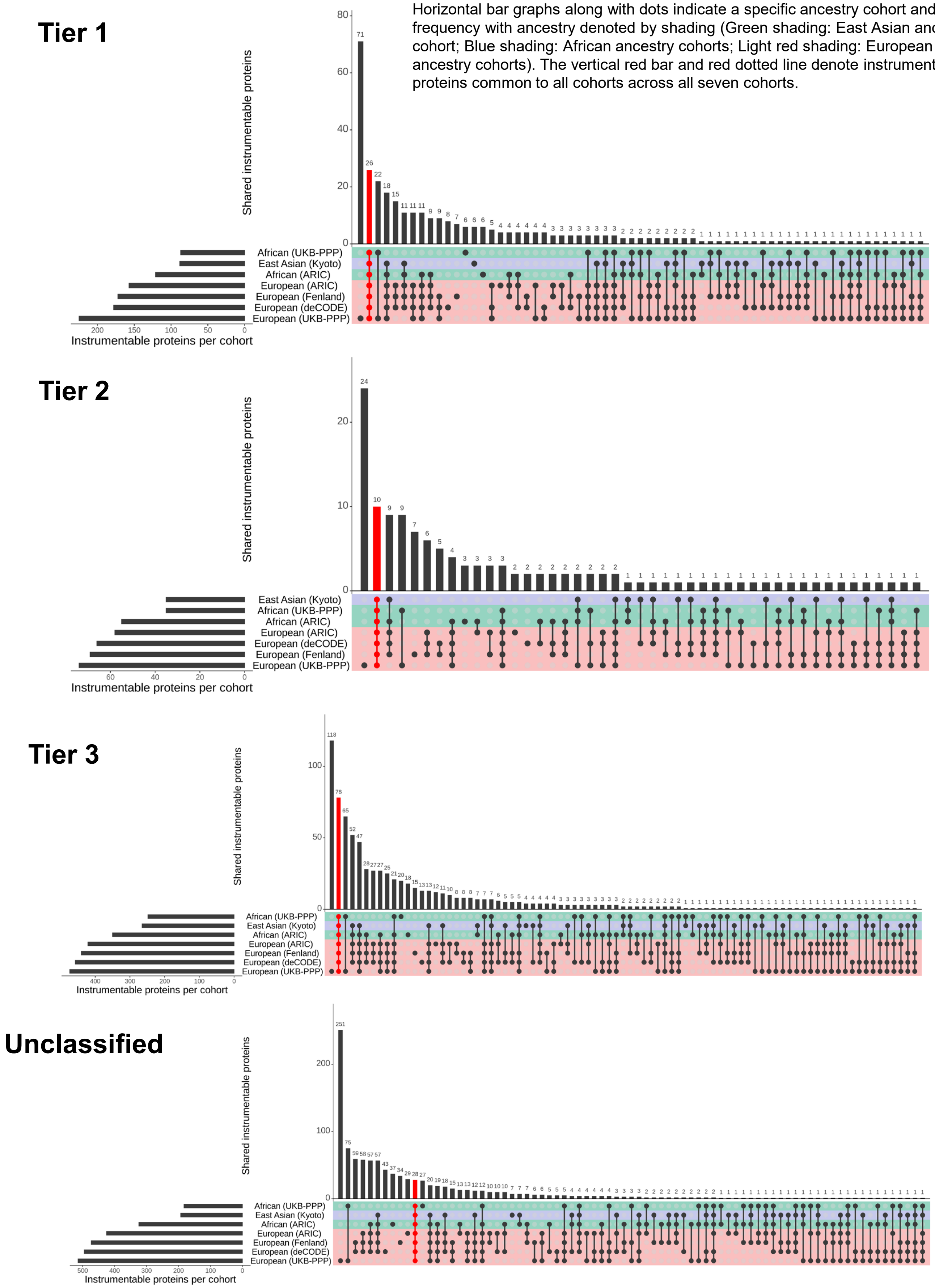

**Supplementary Figure 7. European ancestry druggability heatmaps for 12 disease categories.**

In the following supplementary figures, we show druggability heatmaps for different disease categories in European ancestries.
Cell color displays the MR effect estimate based on Z score averaged across cohorts capped at -10 to +10 with red showing a positive Z score indicating a positive MR effect of the protein on the phenotype and blue showing a negative Z score indicating a negative MR effect of the protein on the phenotype. For simplicity, in European ancestries, we only display protein-phenotypes with consistent effect across European cohorts. The y-axis shows the three drug databases. DrugBank (yellow square): DrugBank shows whether the protein has an available drug in the database. OpenTargets (pink square): Open Targets Platform shows whether the protein has available clinical trial information. Druggability: The druggable genome (as defined by Finan et al.) is shown for Tiers 1 (dark green, representing efficacy targets of approved small molecules and biotherapeutic drugs), Tier 2 (dark purple, representing proteins closely related to approved drug targets or which have associated drug-like compounds), Tier 3 (light purple, representing secreted or extracellular proteins, those distantly related to approved drug targets, and members of important druggable gene families not covered in Tier 1 or Tier 2), and Unclassified (gray, all other proteins not in Tiers 1 to 3). Proteins on the y-axis within each are sorted based on the number of supported databases.

In total, for Europeans, we assessed 15 different disease categories.
However, here, we only show heatmaps for 12 disease categories and we do not show the following 3 disease categories:

- Anthropometry, due to the sheer size
- Biomarker, due to the sheer size and
- Reproductive and urogenital system, since there was only a single association.

The remaining 12 disease categories are as follows:

1. Cardiovascular
- since Figure 7 already shows Tier 1 and Tier 2 for Cardiovascular diseases, we show Cardiovascular Tier 3 and Cardiovascular Unclassified here.

2. Autoimmune
- We show the entire Autoimmune heatmap here (including Tier 3 and Unclassified) while Figure 7 only shows Tier 1 and Tier 2 for Autoimmune diseases.

For the remaining diseases, we do not stratify by druggability tier and show all protein-phenotypes together in the heatmaps.
3. Renal
4. Respiratory
5. Metabolic/endocrine
6. Neurological
7. Psychiatric
8. Musculoskeletal
9. Gastrointestinal
10. Miscellaneous
11. Eye
12. Cancer (all cancers are grouped together for simplicity which encompasses the following) Cancer/Skin
Cancer/Metabolic disease
Cancer Cancer/Respiratory
Cancer/Neurological and psychiatric
Cancer/Gastrointestinal
Cancer/Renal

1. Cardiovascular Tier 3 (Left) and Cardiovascular Unclassified (Right)

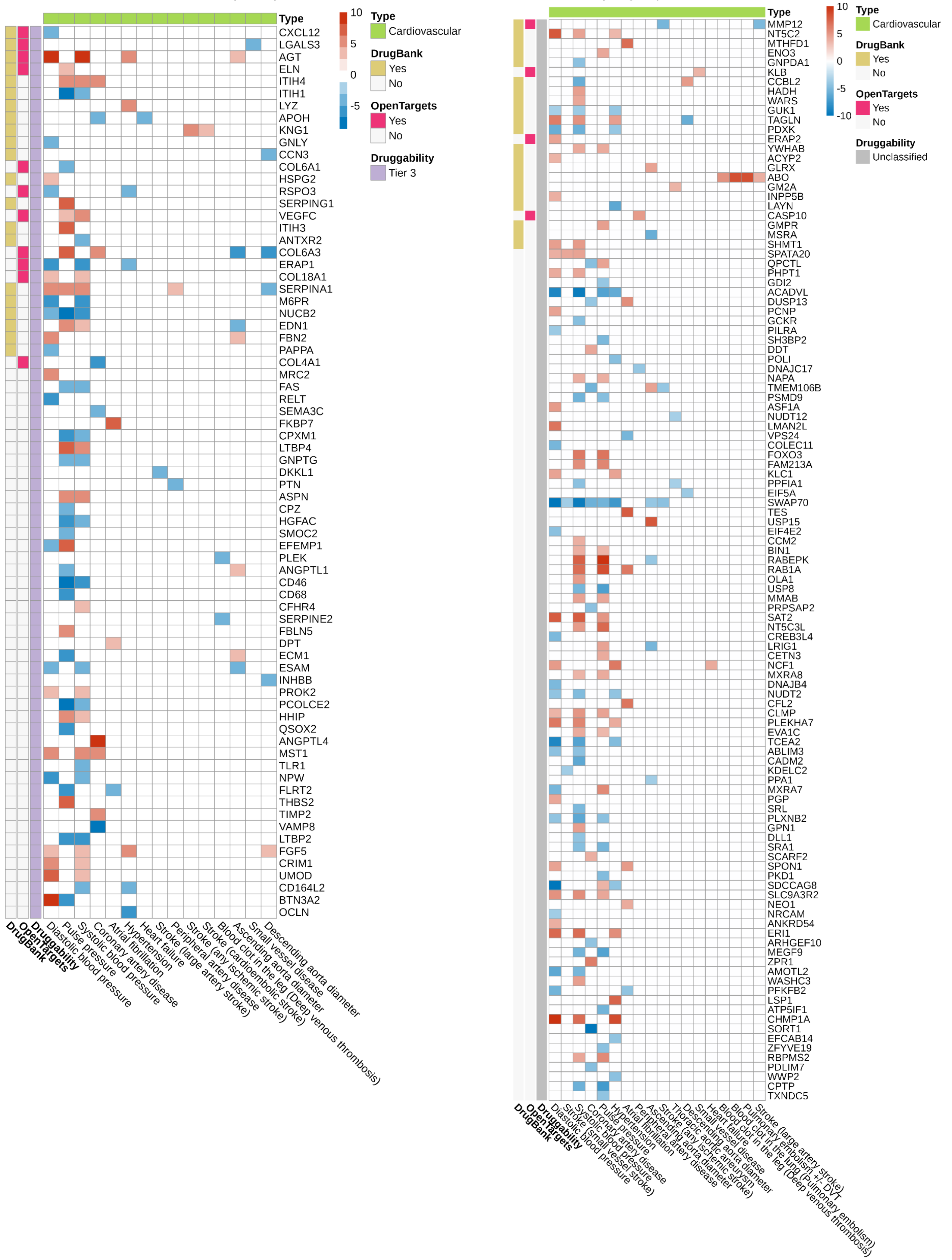

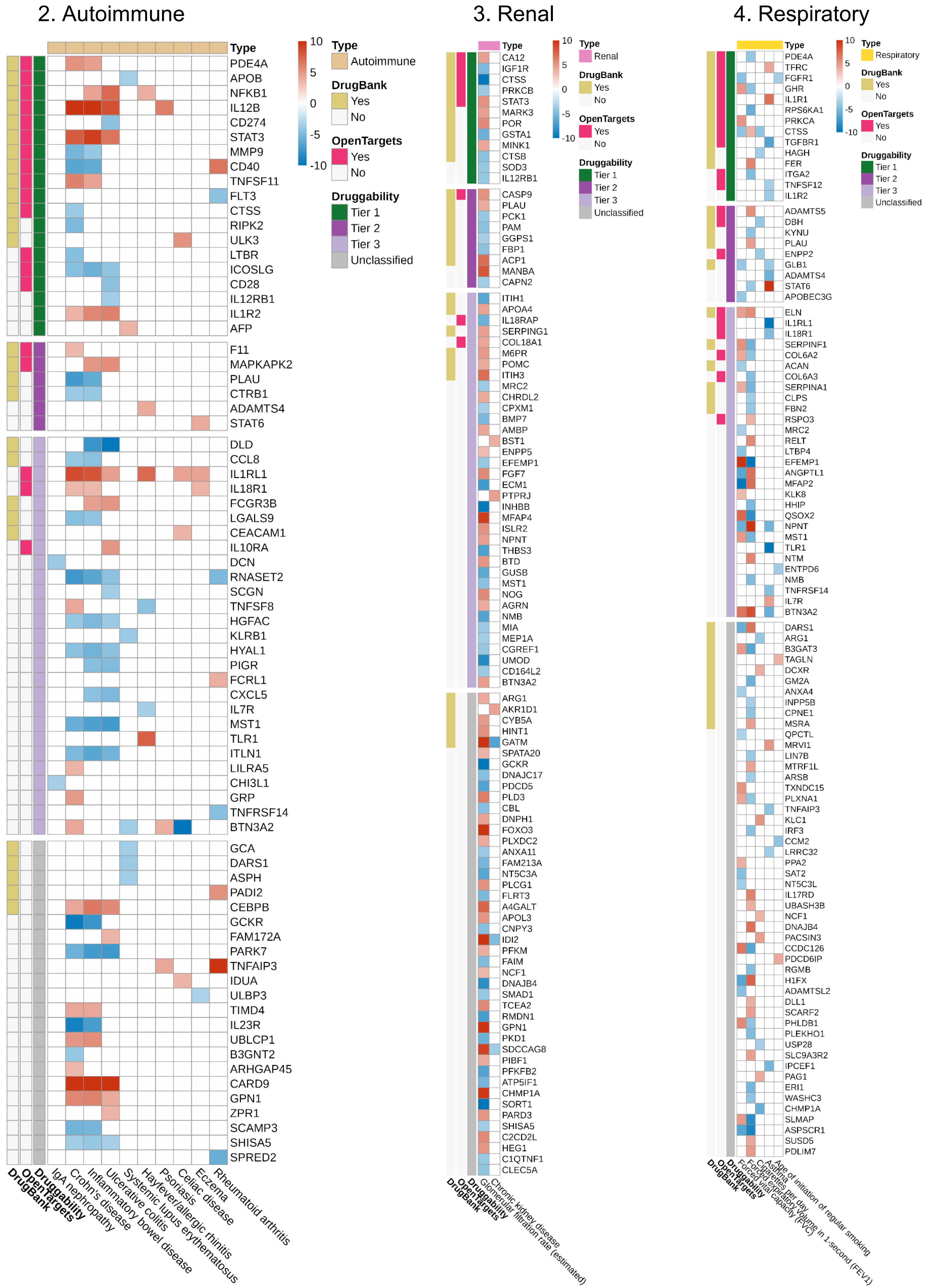
2. Autoimmune
Type
Autoimmune
DrugBank
Yes
No
OpenTargets
Yes
No
Druggability
Tier 1
Tier 2
Tier 3
Unclassified
10
5
0
-5
-10
PDE4A
APOB
NFKB1
IL12B
CD274
STAT3
MMP9
CD40
TNFSF11
FLT3
CTSS
RIPK2
ULK3
LTBR
ICOSLG
CD28
IL12RB1
IL1R2
AFP
F11
MAPKAPK2
PLAU
CTRB1
ADAMTS4
STAT6
DLD
CCL8
IL1RL1
IL18R1
FCGR3B
LGALS9
CEACAM1
IL10RA
DCN
RNASET2
SCGN
TNFSF8
HGFAC
KLRB1
HYAL1
PIGR
FCRL1
CXCL5
IL7R
MST1
TLR1
ITLN1
LILRA5
CHI3L1
GRP
TNFRSF14
BTN3A2
GCA
DARS1
ASPH
PADI2
CEBPB
GCKR
FAM172A
PARK7
TNFAIP3
IDUA
ULBP3
TIMD4
IL23R
UBLCP1
B3GNT2
ARHGAP45
CARD9
GPN1
ZPR1
SCAMP3
SHISA5
SPRED2
DrugBank
OpenTargets
Druggability
IgA nephropathy
Crohn's disease
Inflammatory bowel disease
Ulcerative colitis
Systemic lupus erythematosus
Hayfever/allergic rhinitis
Psoriasis
Celiac disease
Eczema
Rheumatoid arthritis
3. Renal
Type
Renal
DrugBank
Yes
No
OpenTargets
Yes
No
Druggability
Tier 1
Tier 2
Tier 3
Unclassified
10
5
0
-5
-10
CA12
IGF1R
CTSS
PRKCB
STAT3
MARK3
POR
GSTA1
MINK1
CTSB
SOD3
IL12RB1
CASP9
PLAU
PCK1
PAM
GGPS1
FBP1
ACP1
MANBA
CAPN2
ITIH1
APOA4
IL18RAP
SERPING1
COL18A1
M6PR
POMC
ITIH3
MRC2
CHRDL2
CPXM1
BMP7
AMBP
BST1
ENPP5
EFEMP1
FGF7
ECM1
PTPRJ
INHBB
MFAP4
ISLR2
NPNT
THBS3
BTD
GUSB
MST1
NOG
AGRN
NMB
MIA
MEP1A
CGREF1
UMOD
CD164L2
BTN3A2
ARG1
AKR1D1
CYB5A
HINT1
GATM
SPATA20
GCKR
DNAJC17
PDCD5
PLD3
CBL
DNPH1
FOXO3
PLXDC2
ANXA11
FAM213A
NT5C3A
PLCG1
FLRT3
A4GALT
APOL3
CNPY3
IDI2
PFKM
FAIM
NCF1
DNAJB4
SMAD1
TCEA2
RMDN1
GPN1
PKD1
SDCCAG8
PIBF1
PFKFB2
ATP5IF1
CHMP1A
SORT1
PARD3
SHISA5
C2CD2L
HEG1
C1QTNF1
CLEC5A
DrugBank
OpenTargets
Druggability
Chronic kidney disease
Glomerular filtration rate (estimated)
4. Respiratory
Type
Respiratory
DrugBank
Yes
No
OpenTargets
Yes
No
Druggability
Tier 1
Tier 2
Tier 3
Unclassified
10
5
0
-5
-10
PDE4A
TFRC
FGFR1
GHR
IL1R1
RPS6KA1
PRKCA
CTSS
TGFBR1
HAGH
FER
ITGA2
TNFSF12
IL1R2
ADAMTS5
DBH
KYNU
PLAU
ENPP2
GLB1
ADAMTS4
STAT6
APOBEC3G
ELN
IL1RL1
IL18R1
SERPINF1
COL6A2
ACAN
COL6A3
SERPINA1
CLPS
FBN2
RSPO3
MRC2
RELT
LTBP4
EFEMP1
ANGPTL1
MFAP2
KLK8
HHIP
QSOX2
NPNT
MST1
TLR1
NTM
ENTPD6
NMB
TNFRSF14
IL7R
BTN3A2
DARS1
ARG1
B3GAT3
TAGLN
DCXR
GM2A
ANXA4
INPP5B
CPNE1
MSRA
QPCTL
MRVI1
LIN7B
MTRF1L
ARSB
TXNDC15
PLXNA1
TNFAIP3
KLC1
IRF3
CCM2
LRRC32
PPA2
SAT2
NT5C3L
IL17RD
UBASH3B
NCF1
DNAJB4
PACSIN3
CCDC126
PDCD6IP
RGMB
H1FX
ADAMTSL2
DLL1
SCARF2
PHLDB1
PLEKHO1
USP28
SLC9A3R2
IPCEF1
PAG1
ERI1
WASHC3
CHMP1A
SLMAP
ASPSCR1
SUSD5
PDLIM7
DrugBank
OpenTargets
Druggability
Asthma
Cigarettes per day
Forced vital capacity (FVC)
Forced expiratory volume in 1-second (FEV1)
Age of initiation of regular smoking

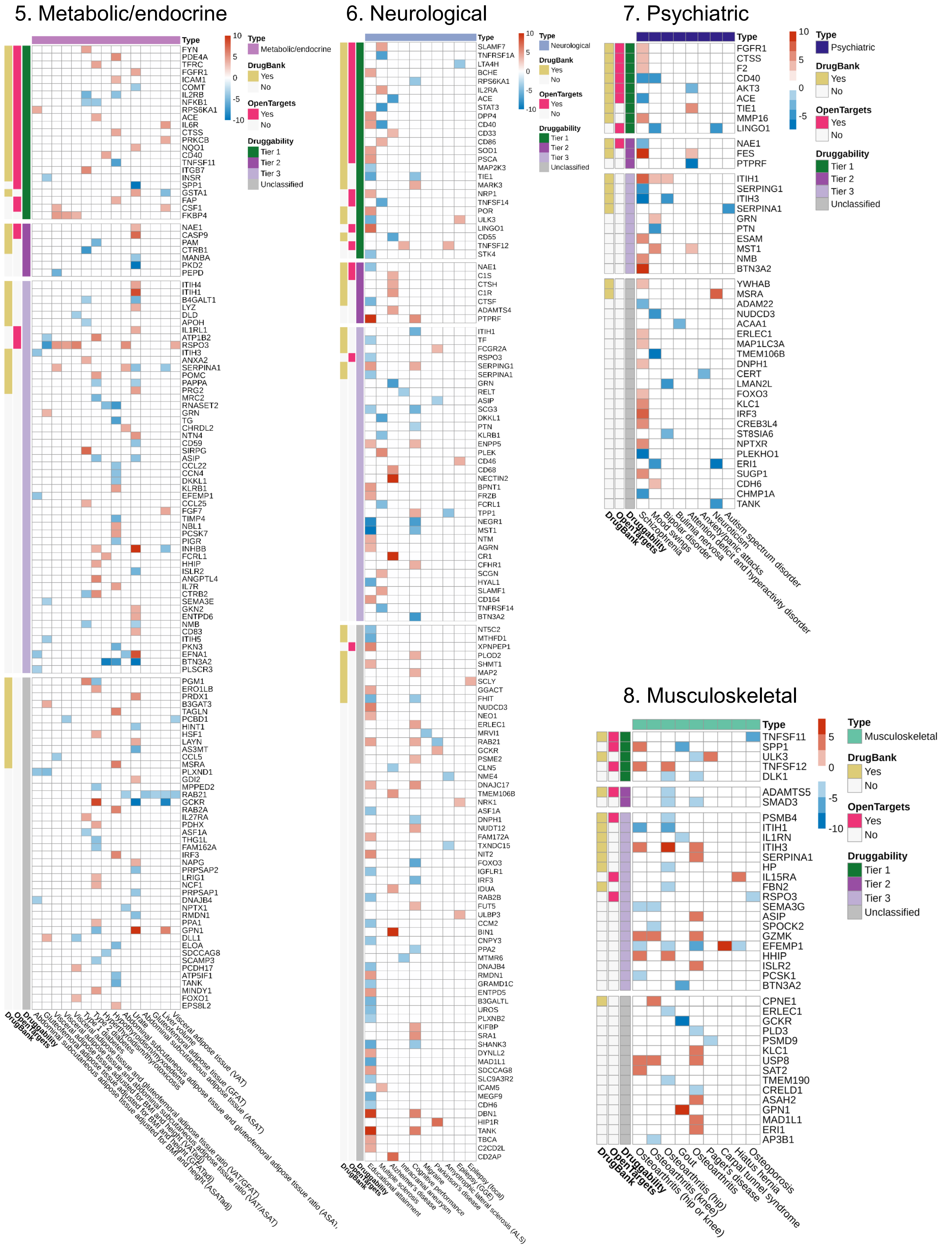

5. Metabolic/endocrine
Type
10
5
0
-5
-10
Type
Metabolic/endocrine
DrugBank
Yes
No
OpenTargets
Yes
No
Druggability
Tier 1
Tier 2
Tier 3
Unclassified
FYN
PDE4A
TFRC
FGFR1
ICAM1
COMT
IL2RB
NFKB1
RPS6KA1
ACE
IL6R
CTSS
PRKCB
NQO1
CD40
TNFSF11
ITGB7
INSR
SPP1
GSTA1
FAP
CSF1
FKBP4
NAE1
CASP9
PAM
CTRB1
MANBA
PKD2
PEPD
ITIH4
ITIH1
B4GALT1
LYZ
DLD
APOH
IL1RL1
ATP1B2
RSPO3
ITIH3
ANXA2
SERPINA1
POMC
PAPPA
PRG2
MRC2
RNASET2
GRN
TG
CHRDL2
NTN4
CD59
SIRPG
ASIP
CCL22
CCN4
DKKL1
KLRB1
EFEMP1
CCL25
FGF7
TIMP4
NBL1
PCSK7
PIGR
INHBB
FCRL1
HHIP
ISLR2
ANGPTL4
IL7R
CTRB2
SEMA3E
GKN2
ENTPD6
NMB
CD83
ITIH5
PKN3
EFNA1
BTN3A2
PLSCR3
PGM1
ERO1LB
PRDX1
B3GAT3
TAGLN
PCBD1
HINT1
HSF1
LAYN
AS3MT
CCL5
MSRA
PLXND1
GDI2
MPPED2
RAB21
GCKR
RAB2A
IL27RA
PDHX
ASF1A
THG1L
FAM162A
IRF3
NAPG
PRPSAP2
LRIG1
NCF1
PRPSAP1
DNAJB4
NPTX1
RMDN1
PPA1
GPN1
DLL1
ELOA
SDCCAG8
SCAMP3
PCDH17
ATP5IF1
TANK
MINDY1
FOXO1
EPS8L2
DrugBank
OpenTargets
Druggability
Abdominal subcutaneous adipose tissue adjusted for BMI and height (ASATadj)
Gluteofemoral adipose tissue adjusted for BMI and height (GFATadj)
Visceral adipose tissue adjusted for BMI and height (VATadj)
Visceral adipose tissue ratio (VAT/ASAT)
Visceral adipose tissue ratio (VAT/GFAT)
Type 1 diabetes
Type 2 diabetes
Hyperthyroidism/thyrotoxicosis
Hypothyroidism/myxoedema
Abdominal subcutaneous and gluteofemoral adipose tissue ratio (ASAT/
Urate
Gluteofemoral adipose tissue (GFAT)
Abdominal subcutaneous adipose tissue (ASAT)
Liver volume
Visceral adipose tissue (VAT)
6. Neurological
Type
10
5
0
-5
-10
Type
Neurological
DrugBank
Yes
No
OpenTargets
Yes
No
Druggability
Tier 1
Tier 2
Tier 3
Unclassified
SLAMF7
TNFRSF1A
LTA4H
BCHE
RPS6KA1
IL2RA
ACE
STAT3
DPP4
CD40
CD33
CD86
SOD1
PSCA
MAP2K3
TIE1
MARK3
NRP1
TNFSF14
POR
ULK3
LINGO1
CD55
TNFSF12
STK4
NAE1
C1S
CTSH
C1R
CTSF
ADAMTS4
PTPRF
ITIH1
TF
FCGR2A
RSPO3
SERPING1
SERPINA1
GRN
RELT
ASIP
SCG3
DKKL1
PTN
KLRB1
ENPP5
PLEK
CD46
CD68
NECTIN2
BPNT1
FRZB
FCRL1
TPP1
NEGR1
MST1
NTM
AGRN
CR1
CFHR1
SCGN
HYAL1
SLAMF1
CD164
TNFRSF14
BTN3A2
NT5C2
MTHFD1
XPNPEP1
PLOD2
SHMT1
MAP2
SCLY
GGACT
FHIT
NUDCD3
NEO1
ERLEC1
MRVI1
RAB21
GCKR
PSME2
CLN5
NME4
DNAJC17
TMEM106B
NRK1
ASF1A
DNPH1
NUDT12
FAM172A
TXNDC15
NIT2
FOXO3
IGFLR1
IRF3
IDUA
RAB2B
FUT5
ULBP3
CCM2
BIN1
CNPY3
PPA2
MTMR6
DNAJB4
RMDN1
GRAMD1C
ENTPD5
B3GALTL
UROS
PLXNB2
KIFBP
SRA1
SHANK3
DYNLL2
MAD1L1
SDCCAG8
SLC9A3R2
ICAM5
MEGF9
CDH6
DBN1
HIP1R
TANK
TBCA
C2CD2L
CD2AP
DrugBank
OpenTargets
Druggability
Educational attainment
Multiple sclerosis
Alzheimer's disease
Intracranial aneurysm
Cognitive performance
Migraine
Parkinson's disease
Amyotrophic lateral sclerosis (ALS)
Epilepsy (GGE)
Epilepsy (focal)
7. Psychiatric
Type
10
5
0
-5
Type
Psychiatric
DrugBank
Yes
No
OpenTargets
Yes
No
Druggability
Tier 1
Tier 2
Tier 3
Unclassified
FGFR1
CTSS
F2
CD40
AKT3
ACE
TIE1
MMP16
LINGO1
NAE1
FES
PTPRF
ITIH1
SERPING1
ITIH3
SERPINA1
GRN
PTN
ESAM
MST1
NMB
BTN3A2
YWHAB
MSRA
ADAM22
NUDCD3
ACAA1
ERLEC1
MAP1LC3A
TMEM106B
DNPH1
CERT
LMAN2L
FOXO3
KLC1
IRF3
CREB3L4
ST8SIA6
NPTXR
PLEKHO1
ERI1
SUGP1
CDH6
CHMP1A
TANK
DrugBank
OpenTargets
Druggability
Schizophrenia
Mood swings
Bipolar disorder
Bulimia nervosa
Attention deficit and hyperactivity disorder
Anxiety/panic attacks
Neuroticism
Autism spectrum disorder
8. Musculoskeletal
Type
5
0
-5
-10
Type
Musculoskeletal
DrugBank
Yes
No
OpenTargets
Yes
No
Druggability
Tier 1
Tier 2
Tier 3
Unclassified
TNFSF11
SPP1
ULK3
TNFSF12
DLK1
ADAMTS5
SMAD3
PSMB4
ITIH1
IL1RN
ITIH3
SERPINA1
HP
IL15RA
FBN2
RSPO3
SEMA3G
ASIP
SPOCK2
GZMK
EFEMP1
HHIP
ISLR2
PCSK1
BTN3A2
CPNE1
ERLEC1
GCKR
PLD3
PSMD9
KLC1
USP8
SAT2
TMEM190
CRELD1
ASAH2
GPN1
MAD1L1
ERI1
AP3B1
DrugBank
OpenTargets
Druggability
Osteoarthritis (hip or knee)
Osteoarthritis (knee)
Gout
Osteoarthritis (hip)
Paget's disease
Carpal tunnel syndrome
Hiatus hernia
Osteoporosis

#### 9. Gastrointestinal

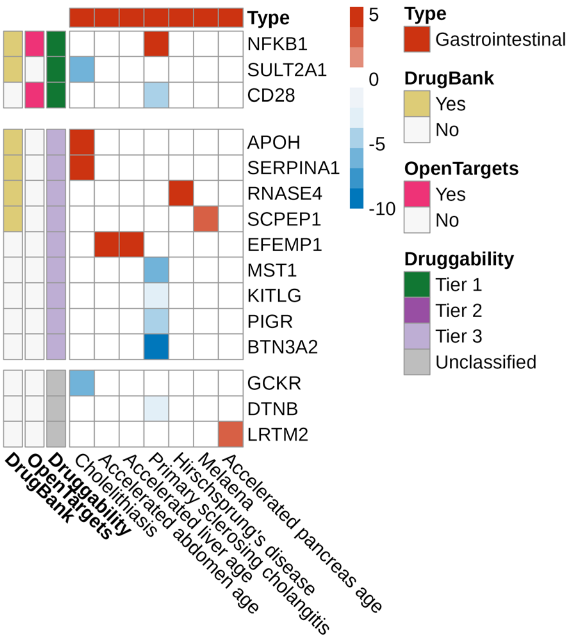


#### 10. Miscellaneous

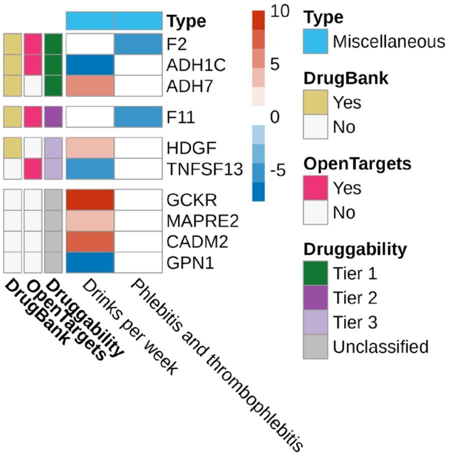


#### 11. Eye

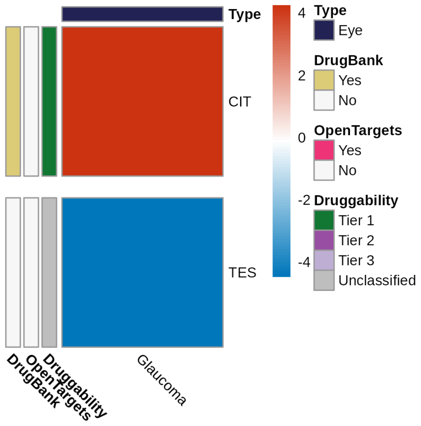


#### 12. Cancer

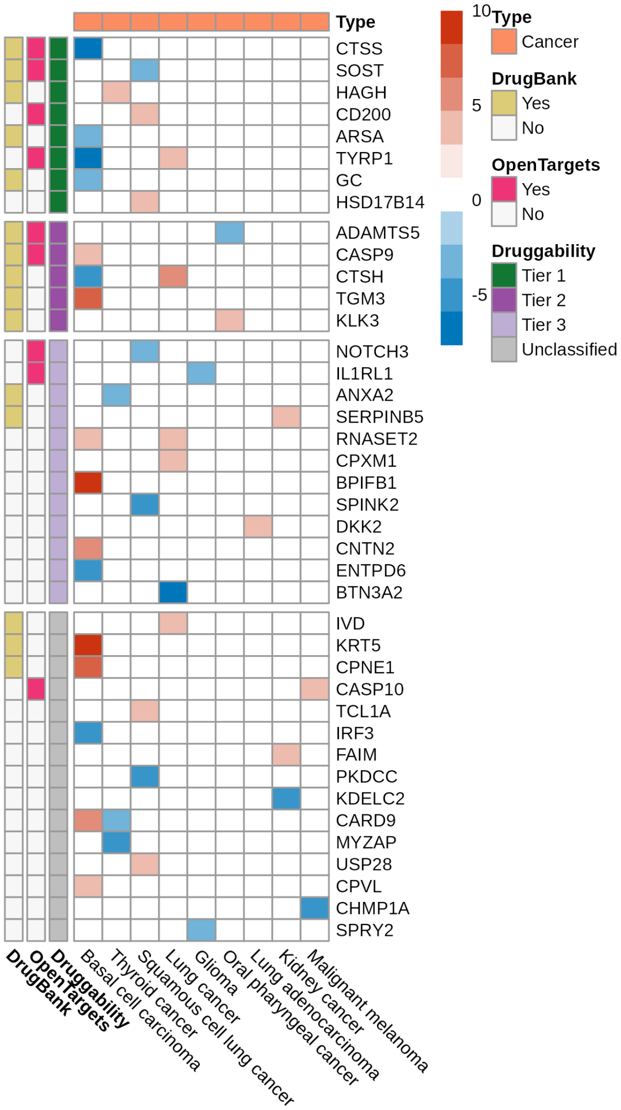

**Supplementary Figure 8. East Asian ancestry druggability heatmaps for 11 disease categories.**

In the following supplementary figures, we show druggability heatmaps for East Asian disease categories. Cell color displays the MR effect estimate based on Z score averaged across cohorts capped at -10 to +10 with red showing a positive Z score indicating a positive MR effect of the protein on the phenotype and blue showing a negative Z score indicating a negative MR effect of the protein on the phenotype. For simplicity, in European ancestries, we only display protein-phenotypes with consistent effect across European cohorts. The y-axis shows the three drug databases. DrugBank (yellow square): DrugBank shows whether the protein has an available drug in the database. OpenTargets (pink square): Open Targets Platform shows whether the protein has available clinical trial information. Druggability: The druggable genome (as defined by Finan et al.) is shown for Tiers 1 (dark green, representing efficacy targets of approved small molecules and biotherapeutic drugs), Tier 2 (dark purple, representing proteins closely related to approved drug targets or which have associated drug-like compounds), Tier 3 (light purple, representing secreted or extracellular proteins, those distantly related to approved drug targets, and members of important druggable gene families not covered in Tier 1 or Tier 2), and Unclassified (gray, all other proteins not in Tiers 1 to 3). Proteins on the y-axis within each are sorted based on the number of supported databases.

In total, for East Asians, we assessed 14 different disease categories.
However, here, we only show heatmaps for 11 disease categories and we do not show the following 3 disease categories:

- Renal, since there was only a single association.
- Eye, since there was only a single association.
- Autoimmune, since the heatmap for all proteins is already shown in Figure 8.

For the remaining diseases, we do not stratify by druggability tier and show all protein-phenotypes together in the heatmaps. All diseases are shown separately except for Neurological and Psychiatric which we group together in one heatmap for simplicity. The remaining 11 disease categories are as follows:

1. Cardiovascular

- We show the entire Cardiovascular heatmap here (including Tier 3 and Unclassified) while Figure 8 only shows Tier 1 and Tier 2 for Cardiovascular diseases.
- 2. Respiratory
- 3. Metabolic/endocrine
- 4. Neurological+Psychiatric (shown together for simplicity)
- 5. Gastrointestinal
- 6. Miscellaneous
- 7. Cancer
- 8. Haematology
- 9. Anthropometry
- 10. Biomarker

#### 1. Cardiovascular

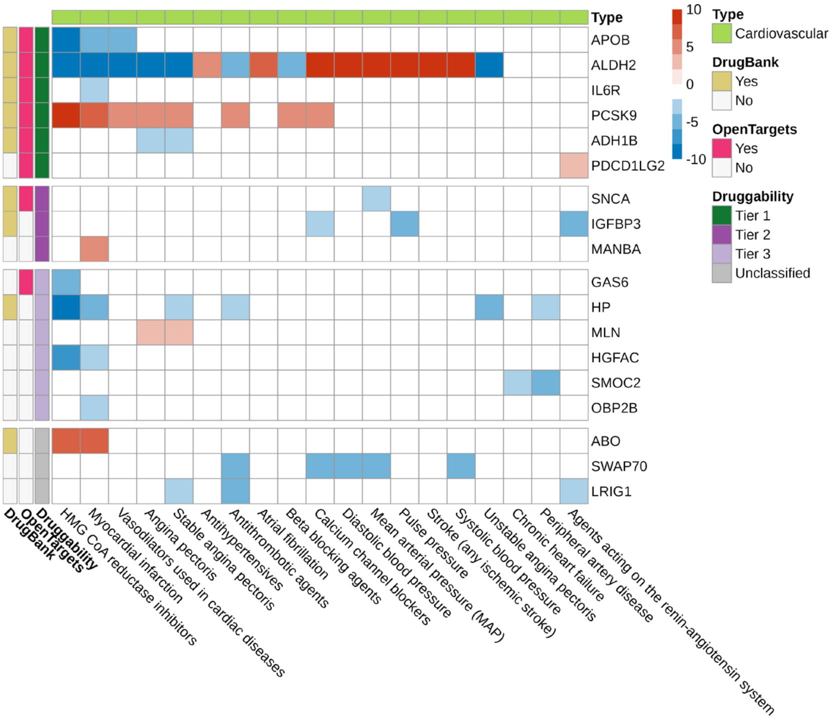


#### 2. Respiratory

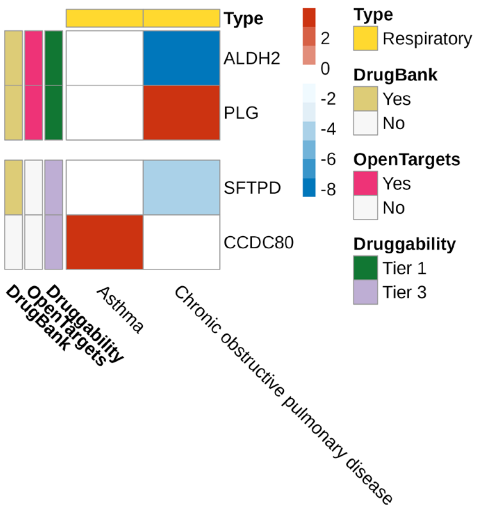


#### 3. Metabolic/endocrine

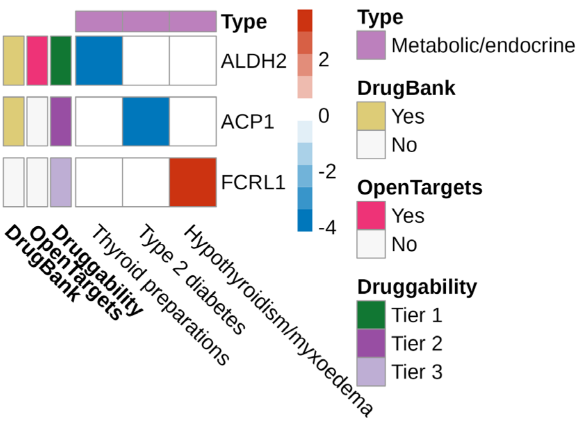


#### 4. Neurological and Psychiatric (shown together for simplicity)

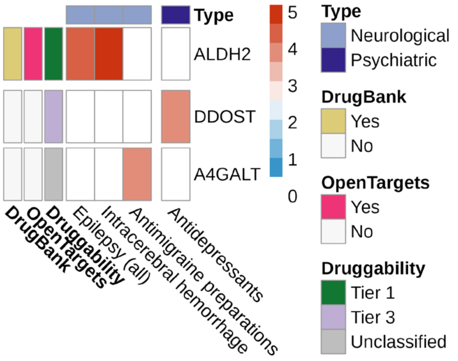


#### 5. Gastrointestinal

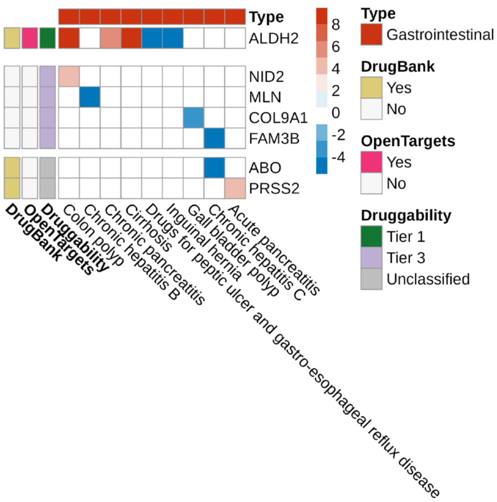


#### 6. Miscellaneous

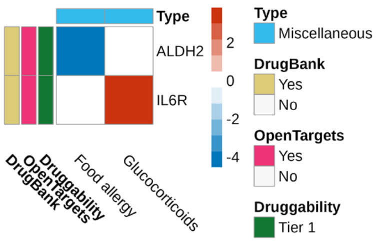

#### 7. Cancer

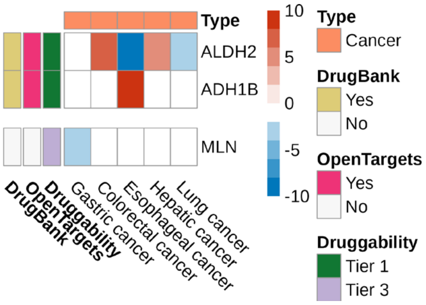


#### 9. Anthropometry

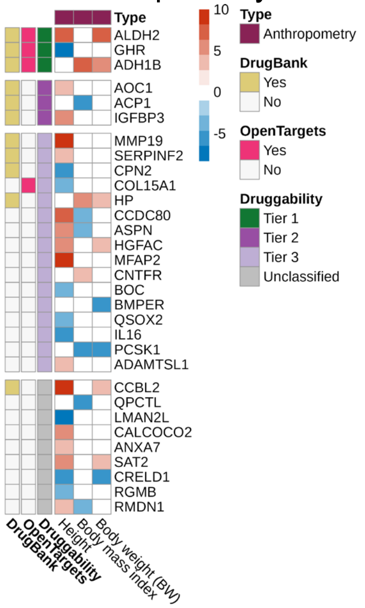


#### 8. Haematology

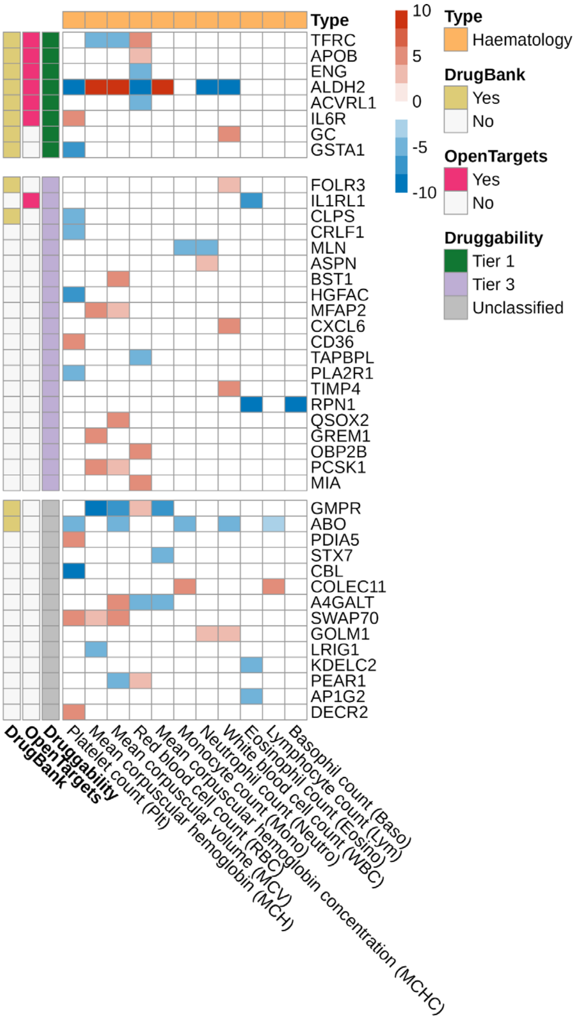


#### 10. Biomarker

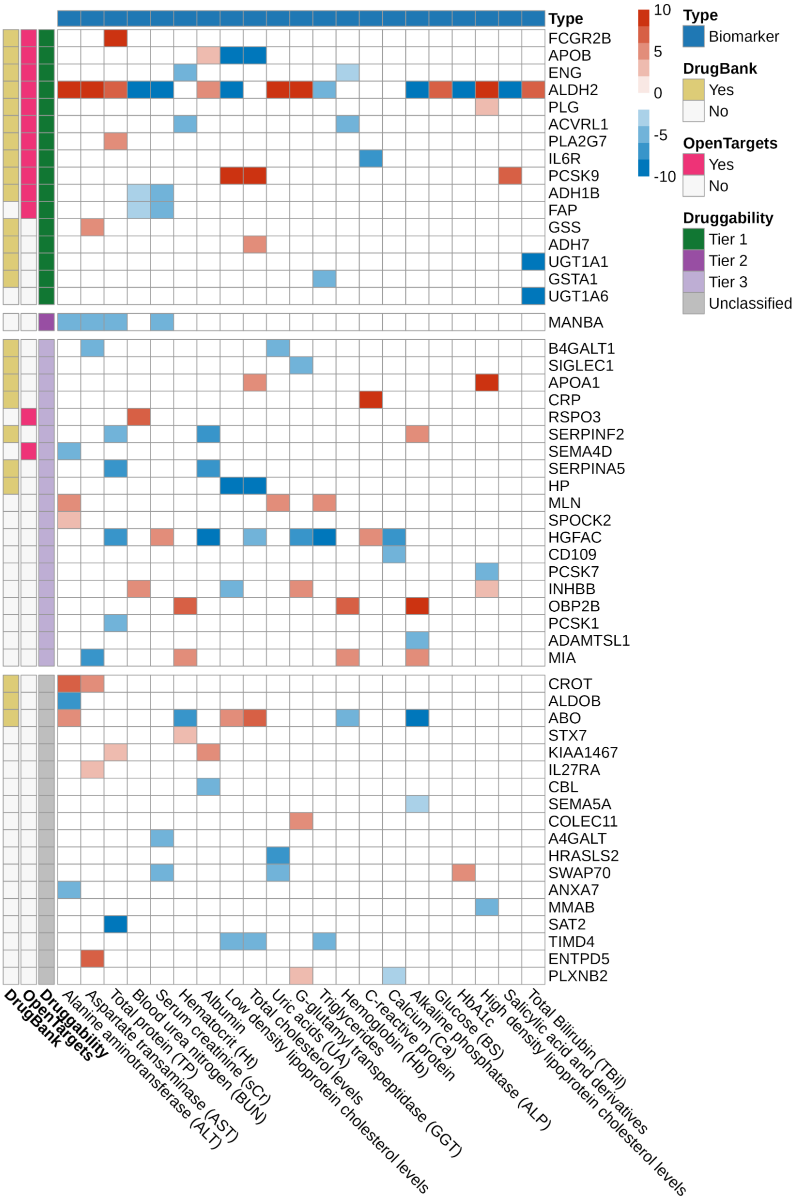

**Supplementary Figure 9. Prioritizing proteins for coronary artery disease and type 2 diabetes.**

Flow diagram showing filtering steps for MR estimates and Cox regression observational estimates to identify potential targets for (a) coronary artery disease and (b) type 2 diabetes. CAD = coronary artery disease; OR = odds ratio; T2D: Type 2 diabetes.

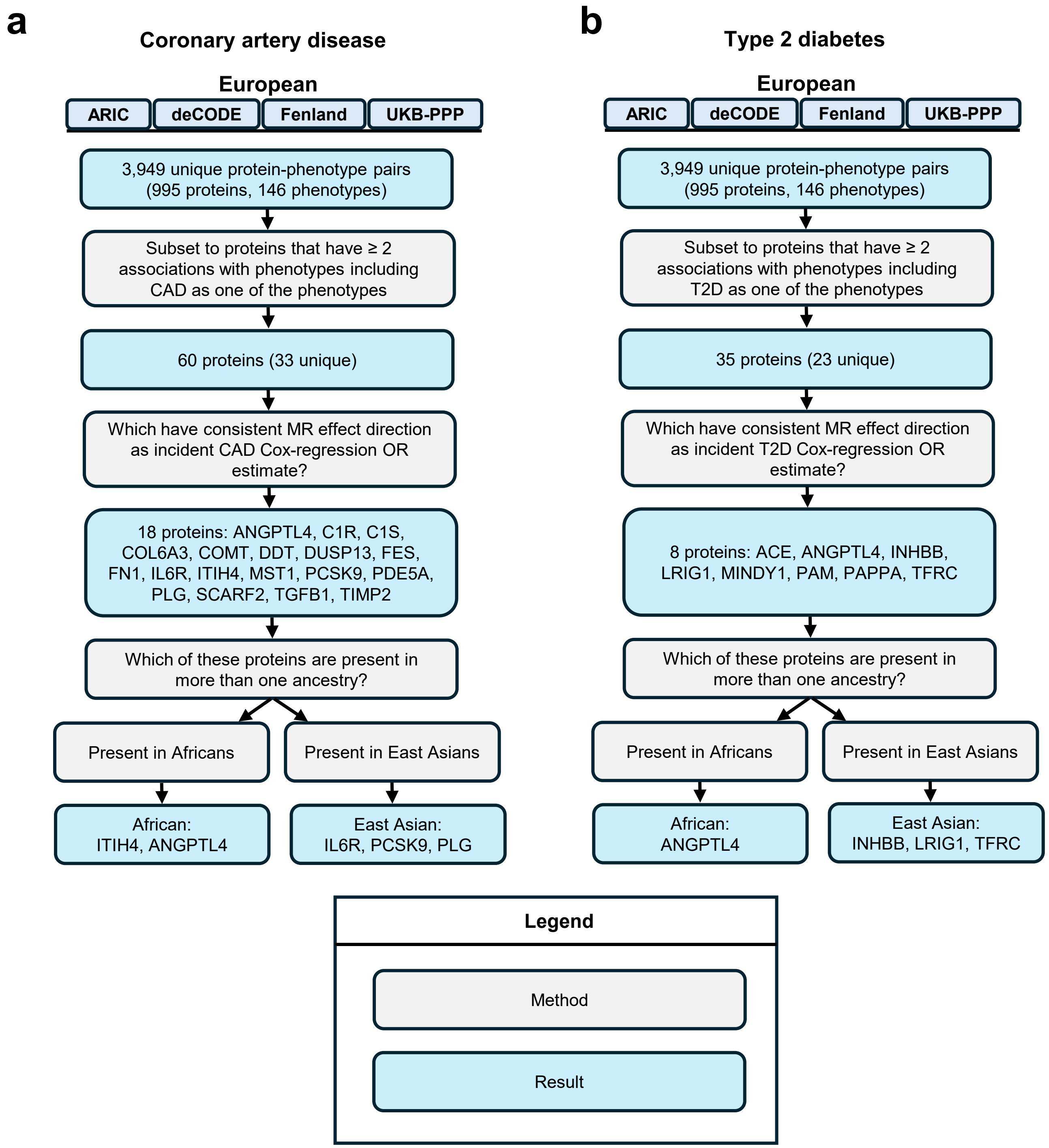

#### Supplementary Note-only References


1. Burgess, S., Davies, N. M. & Thompson, S. G. Bias due to participant overlap in two-sample Mendelian randomization. *Genetic Epidemiology* **40**, 597–608 (2016).
2. Yuan, S. *et al.* Plasma proteins and onset of type 2 diabetes and diabetic complications: Proteome-wide Mendelian randomization and colocalization analyses. *Cell Reports Medicine* **4**, 101174 (2023).
3. High-Throughput Characterization of Blood Serum Proteomics of IBD Patients with Respect to Aging and Genetic Factors | PLOS Genetics. https://journals.plos.org/plosgenetics/article?id=10.1371/journal.pgen.1006565.
4. Identification of Common and Rare Genetic Variation Associated With Plasma Protein Levels Using Whole-Exome Sequencing and Mass Spectrometry | Circulation: Genomic and Precision Medicine. https://www.ahajournals.org/doi/10.1161/CIRCGEN.118.002170.
5. Yoshiji, S. *et al.* COL6A3-derived endotrophin mediates the effect of obesity on coronary artery disease: an integrative proteogenomics analysis. 2023.04.19.23288706 Preprint at https://doi.org/10.1101/2023.04.19.23288706 (2023).
6. Sun, B. B. *et al.* Genomic atlas of the human plasma proteome. *Nature* **558**, 73–79 (2018).
7. Zheng, J. *et al.* Phenome-wide Mendelian randomization mapping the influence of the plasma proteome on complex diseases. *Nat Genet* **52**, 1122–1131 (2020).
8. Ravindran, A. *et al.* Translatome profiling reveals Itih4 as a novel smooth muscle cell–specific gene in atherosclerosis. *Cardiovascular Research* cvae028 (2024) doi:10.1093/cvr/cvae028.
9. Koprulu, M. *et al.* Proteogenomic links to human metabolic diseases. *Nat Metab* **5**, 516–528 (2023).
10. Yoshiji, S. *et al.* Proteome-wide Mendelian randomization implicates nephronectin as an actionable mediator of the effect of obesity on COVID-19 severity. *Nat Metab* **5**, 248–264 (2023).

16. Common CD36 SNPs reduce protein expression and may contribute to a protective atherogenic profile | Human Molecular Genetics | Oxford Academic. https://academic.oup.com/hmg/article/20/1/193/2386020.

24. Rosenson Robert S. *et al.* Zodasiran, an RNAi Therapeutic Targeting ANGPTL3, for Mixed Hyperlipidemia. *New England Journal of Medicine* **0**,.

25. Yang, J. *et al.* Conditional and joint multiple-SNP analysis of GWAS summary statistics identifies additional variants influencing complex traits. *Nat Genet* **44**, 369–375 (2012).

26. Giambartolomei, C. *et al.* Bayesian Test for Colocalisation between Pairs of Genetic Association Studies Using Summary Statistics. *PLOS Genetics* **10**, e1004383 (2014).

27. Robinson, J. W. *et al.* An efficient and robust tool for colocalisation: Pair-wise Conditional and Colocalisation (PWCoCo). 2022.08.08.503158 Preprint at https://doi.org/10.1101/2022.08.08.503158 (2022).

28. Zhang, W. *et al.* SharePro: an accurate and efficient genetic colocalization method accounting for multiple causal signals. *Bioinformatics* **40**, btae295 (2024).
